# Algorithmic Fairness of QPrediction Cardiometabolic Risk Prediction Models

**DOI:** 10.1101/2025.08.20.25333669

**Authors:** Inchuen Huynh, Denise Utochkin, Alexandros Katsiferis, Tri-Long Nguyen, Tibor V Varga

## Abstract

**Background:** There is a lack of fairness evaluation of clinical prediction models used in routine practice. This study aimed to evaluate the fairness of three established risk prediction models belonging to the QPrediction family: QRISK3, QDiabetes, and QStroke, which estimate 10-year risks for cardiovascular disease, type 2 diabetes, and stroke, respectively.

**Methods and findings:** We used data from the UK Biobank, a large-scale prospective cohort study comprising 502,131 participants. To assess calibration, the predicted risks were compared to the observed cumulative incidence rates, and survival Brier scores were calculated as a summary metric of model calibration. To assess discrimination, we calculated overall and group-specific true positive, true negative, false positive, and false negative rates (TPR, TNR, FPR, FNR). We compared the rates between subgroups of demographics and considered prediction parity when values were close to each other to a prespecified degree. Overall, all models showed systematic overprediction of risks for the UK Biobank validation cohorts, with the degree of overprediction varying between the models. Prediction parity was not reached in FNR and TNR in terms of ethnicity, education level, average household income before taxation, and Townsend deprivation score. For immigration status, there were no major differences in the discrimination metrics, and prediction parity was achieved. For QDiabetes, the trends were similar, apart from ethnicity, where the opposite was observed as for QRISK3.

**Conclusions:** Inclusion of ethnicity and deprivation in the models may have contributed to more consistent calibration across subgroups. Variables were measured only at baseline, while many social and economic conditions can shift over time. Although including social determinants of health can improve predictive performance, the models are typically deployed in a clinical setting to guide individual-level decision making, which can inadvertently shift responsibility for health outcomes onto patients.

## 1 Introduction

Prognostic predictive models enable the identification of individuals at risk of a disease, providing the opportunity for early preventive interventions. Although hundreds of cardiometabolic clinical prediction models have been published to date [1][2], only a handful of these models have been incorporated into national guidelines [3] or clinical practice [4][5].

In the process of validating prognostic prediction models, the performance of models is most often assessed based on their ability to provide overall accurate predictions, while the fairness of the provided predictions has received less attention. In recent years, a number of alarming reports emerged showcasing algorithms and healthcare processes that benefit privileged population subgroups, while underperforming among those vulnerable [6][7][8][9]. The reasons for such results are manifold but often include developing models using imbalanced datasets, using inappropriate feature or outcome proxies, omitting key socioeconomic determinants of health (SDOH) in model development, and failing to co-create models with vulnerable communities to avoid knowledge gaps in model creation and to assess the needs of the patients [10][11][12].

Algorithmic fairness has become a rapidly developing field of science with the overall goal to conceptualize, quantify, and mitigate biases arising from prediction models. Fairness aspects of predictive models have been gaining increasing attention in recent years, with TRIPOD+AI [13], the prevailing consensus guideline established to improve the reporting of prediction models, calling for all papers reporting on diagnostic or prognostic prediction models to address aspects of fairness. *Fairness* is a subjective term, making the assessment of prediction models from a fairness perspective a multifaceted and complex, if not impossible, task. In this project, we define fairness as a property of predictive models to provide an equal average prediction for prespecified groups in populations based on various attributes that might make a group more vulnerable.

In this article, we evaluate the fairness of three established risk prediction models belonging to the QPrediction family: QRISK3 [14], QDiabetes [15], and QStroke [16], predicting future risk of cardiovascular disease (CVD), type 2 diabetes, and stroke, respectively. All three tools are publicly accessible, with QRISK3, QDiabetes, and QStroke being integrated into primary clinical practice in the United Kingdom (UK), used by over 55% of general practices in England [17] and 46% of general practices in Wales [18], and being the most widely used primary care clinical software in Scotland [19]. QRISK3 is recommended for clinical use in the UK by the National Institute for Health and Care Excellence (NICE) [3]. For risk identification of type 2 diabetes, NICE recommends using computer-based risk assessment tools [20] and up until 2019, QDiabetes was included in the list of tools recommended in the NHS Health Check best practice guideline [21].

Although the performance of QPrediction models has been extensively validated in previous articles [22][23][24][25], fairness aspects have not been featured in these studies. In this article, we use data from the UK Biobank for validation, a longitudinal prospective cohort study with over half a million participants from the UK with extensive information about their health status, sociodemographic information, lifestyle, and other factors.

## 2 Methods

### 2.1 Design and Study Population

The UK Biobank is a longitudinal prospective cohort study encompassing over half a million participants from the UK. Extensive individual-level data is available on participants’ lifestyles, SDOH parameters, biomarkers, and morbidities. Participants were recruited during 2006-2010 and were between ages 40-69 at the time of recruitment. Four repeat assessments were carried out during 2012-2019. At baseline, questionnaire and interview data, physical measurements, and samples of blood, urine, and saliva were collected as described previously [26]. Follow-up on the participants is conducted through linkages to routinely available national datasets, including death registrations, cancer registration, hospital inpatient episodes, hospital outpatient episodes, and primary care data. The hospital inpatient data used to identify disease incidence for this article covers individual patient journeys up until 2022. This study involves human participants and was approved by the National Health Service’s National Research Ethics Service North West (21/NW/0157). Participants gave informed consent to participate in the study before taking part.

We applied different exclusion criteria to assess each of the QPrediction models, as described in Figure 1. For all of the models, participants with missing Townsend deprivation scores were excluded as well as those with preexisting diagnoses of the disease outcome of interest. Additionally, participants using statins at study entry were excluded from the study population for QRISK3 and those using anticoagulants were excluded from the study population for QStroke. The diagnostic codes used to define the exclusion criteria related to pre-existing diseases correspond to those defined in the original articles for the development of the models.

**Figure 1:**
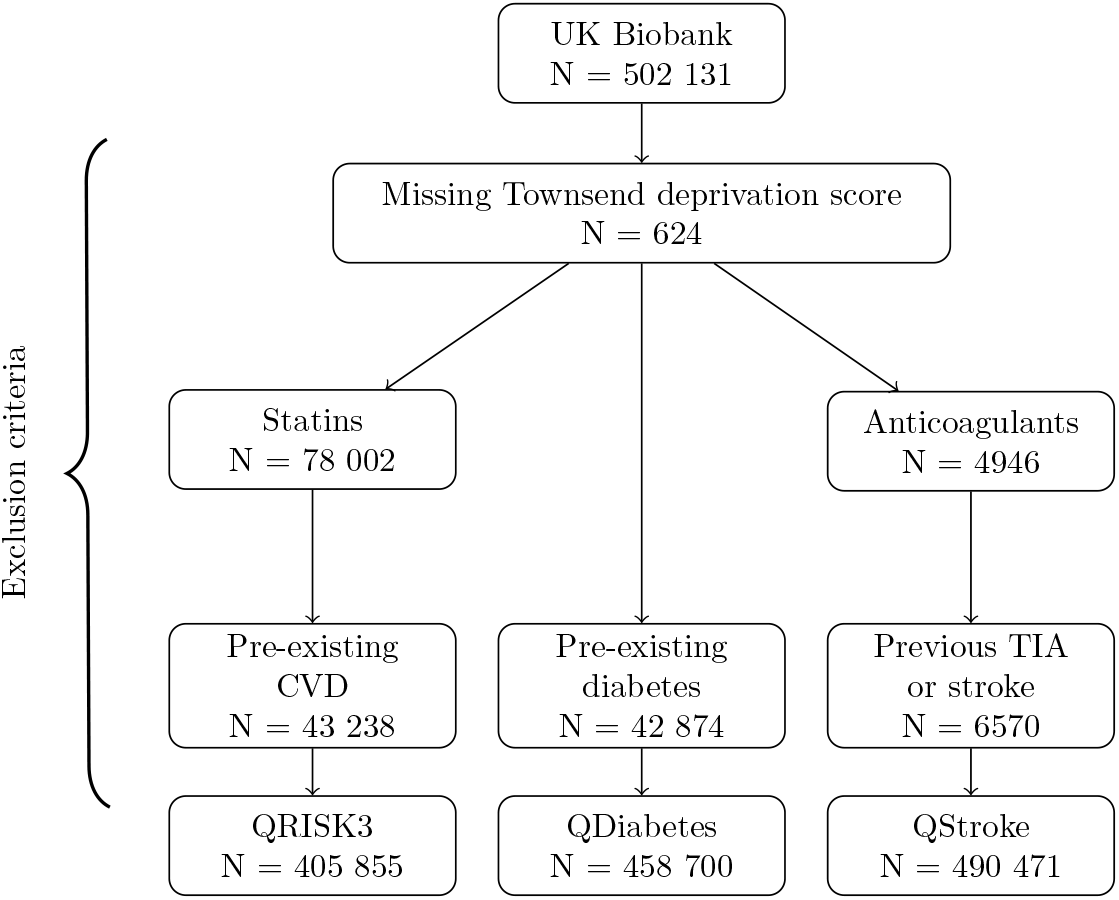
Flowchart of exclusion criteria for each QPrediction model. Note that there are overlaps between groups with various exclusion criteria, therefore the total number of excluded individuals is less than the sum of the individuals under each exclusion criterion.

### 2.2 Variables

We considered variables measured at the baseline assessment to allow for the longest possible follow-up times. The variables were matched to the risk factors included in the QPrediction models (Table S2.1.1-S2.1.3) using data from the baseline assessment as well as from data linkages. For the QRISK3-model, the variables matched were extracted from a previous validation study using UK Biobank data [27], whereas, for the QDiabetes and QStroke models, variables were matched as described in Table S2.2.1-S2.3.3. When there were no direct matches in available UK Biobank variables for a variable in the QPrediction model, the closest possible match was chosen. The QDiabetes model had three variations and the model chosen to calculate the risk score was based on whether the values for HbA1c or fasting glucose were available, or neither. Since the developers found the model including HbA1c without fasting blood glucose to be the best performing [15], we chose to validate this model.

**Table 1:**
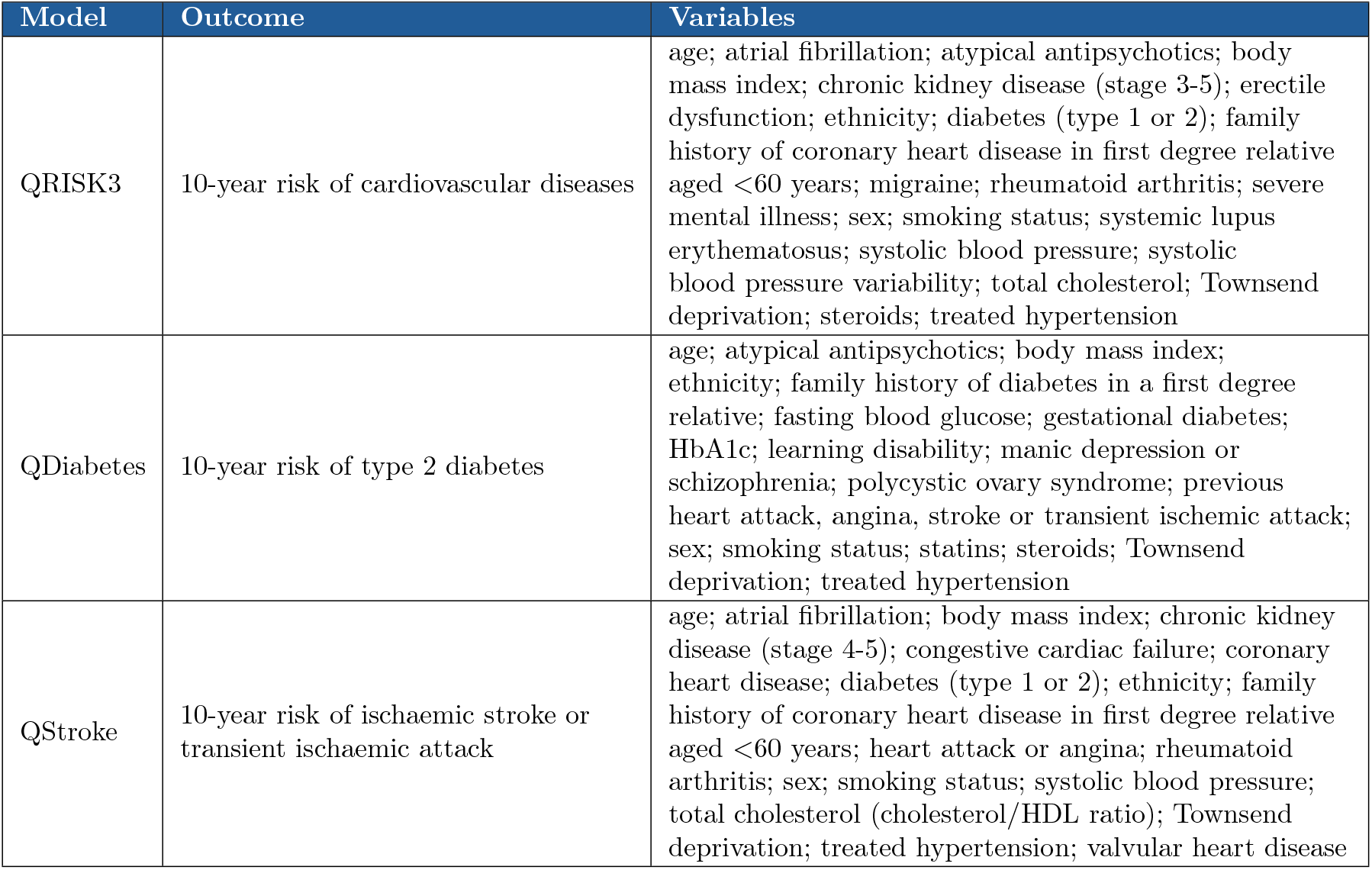
Outcomes and variables included in QRISK3, QDiabetes, and QStroke models.

| Model | Outcome | Variables |
| --- | --- | --- |
| QRISK3 | 10-year risk of cardiovascular diseases | age; atrial fibrillation; atypical antipsychotics; body mass index; chronic kidney disease (stage 3-5); erectile dysfunction; ethnicity; diabetes (type 1 or 2); family history of coronary heart disease in first degree relative aged <60 years; migraine; rheumatoid arthritis; severe mental illness; sex; smoking status; systemic lupus erythematosus; systolic blood pressure; systolic blood pressure variability; total cholesterol; Townsend deprivation; steroids; treated hypertension |
| QDiabetes | 10-year risk of type 2 diabetes | age; atypical antipsychotics; body mass index; ethnicity; family history of diabetes in a first degree relative; fasting blood glucose; gestational diabetes; HbA1c; learning disability; manic depression or schizophrenia; polycystic ovary syndrome; previous heart attack, angina, stroke or transient ischemic attack; sex; smoking status; statins; steroids; Townsend deprivation; treated hypertension |
| QStroke | 10-year risk of ischaemic stroke or transient ischaemic attack | age; atrial fibrillation; body mass index; chronic kidney disease (stage 4-5); congestive cardiac failure; coronary heart disease; diabetes (type 1 or 2); ethnicity; family history of coronary heart disease in first degree relative aged <60 years; heart attack or angina; rheumatoid arthritis; sex; smoking status; systolic blood pressure; total cholesterol (cholesterol/HDL ratio); Townsend deprivation; treated hypertension; valvular heart disease |

The variables used to define the subgroups for fairness assessments were sex (men/women), age group (35-45/46-55/56-65/66-75), self-reported ethnicity (White/Asian or Asian British/Black or Black British/Chinese/Mixed/Other ethnic group/Prefer not to answer/Do not know), immigration status (no/yes), Townsend deprivation score (binned to quantiles: [−7,−4]/(−4,−2]/(−2,1]/(1,11]), index of multiple deprivation ([0,20]/(20,40]/(40,60]/(60,80]/(80,100]), average household income (Greater than 100,000/52,000 to 100,000/31,000 to 51,999/18,000 to 30,999/Less than 18,000/Prefer not to answer/Do not know), highest education level (College or University degree/National Vocational Qualification (NVQ) or Higher National Diploma (HND) or Higher National Certificate (HNC) or equivalent/Other prof. qual. eg: nursing, teaching/A levels or AS levels or equivalent/O levels or General Certificate of Secondary Education (GCSEs) or equivalent/Certificate of Secondary Education (CSEs) or equivalent/None of the above/Prefer not to answer), with the assumed privileged groups presented first in the parentheses above.

### 2.3 Outcomes

Outcomes of interest for the QPrediction models were defined using the International Classification of Diseases (ICD). For the validation data, we matched the codings to the ICD10 and ICD9 (ICD version 9 and 10) equivalents as well as the Office of Population Censuses and Surveys Classification of Interventions and Procedures version 4 (OPCS-4) codes from hospital episode statistics and death registration data. The matching process is described in detail in Table S3. Follow-up time was measured in years defined as time from the date of baseline assessment until the date of an event of cardiovascular disease, type 2 diabetes, or stroke, death from other causes than the event of interest, loss to follow-up, or the administrative censoring date of the hospital inpatient data in the UK Biobank, which differed by country (England: 31 October 2022; Scotland: 31 August 2022; Wales: 31 May 2022) [28].

### 2.4 Missing Data Strategy

Missing data was imputed using multiple imputation by chained equations (mice) using the *mice* R package [29]. We assumed a missing-at-random missingness pattern. A total of five imputed copies were generated based on the percentage of missingness in the variables (2.8%) [30]. Convergence was checked visually for all the datasets, and the baseline characteristics were checked before and after the imputations. Results were pooled using Rubin’s rules [31].

### 2.5 Algorithmic Fairness in Calibration

Numerous *algorithmic group fairness* metrics have been developed to assess equal predictive performance; therefore, in general, it is paramount for researchers to accurately outline the purpose of the utilized metrics in relation to the goals of research projects. In our work, we measured algorithmic group fairness through two distinct routes: by quantifying differences in the calibration of models across groups and in prediction error to gain a holistic picture of performance parity.

The fairness of the QPrediction models was evaluated using metrics of calibration and discrimination. The purpose of calibration is to quantify the agreement between observed risks and predictions obtained from the models. Here, we compared the predicted probabilities of the QPrediction models to the observed cumulative incidences derived from weighted Kaplan-Meier estimates. The Kaplan-Meier estimates were weighted using inverse probability censoring weighting (IPCW) to account for competing risk, which occurred when a participant experienced a competing event that prevented them from experiencing the event of interest. Competing risk can be thought of as a special case of informative censoring, where the censoring mechanism is associated with the event of interest, for example, by sharing similar risk factors. Not accounting for competing risk and informative censoring could lead to an overestimation of the risk since the population is not representative of the true population at risk of experiencing the event of interest [32]. With IPCW, the uncensored population is up-weighted for each participant lost to follow-up due to informative censoring [33][34]. The derived weighted Kaplan-Meier risk estimates were validated against the correct Aalen-Johansen risk estimates of the event of interest. In case the weighted Kaplan-Meier estimates matched the ones derived using the Aalen-Johanssen estimator, we deemed the IPCW method as properly accounting for competing risk. The weights obtained using IPCW were used to calculate survival Brier scores, as the time-dependent mean squared difference of the predicted and observed values, as a summary metric of model calibration. The Brier scores have a theoretical range of 0-1, 0 representing perfect calibration, and 1 representing the poorest calibration. For a visual check of calibration, the predicted risks (obtained from the QPrediction models) and observed risks (obtained from weighted Kaplan-Meier estimates) were binned into deciles based on the predicted risk scores and plotted against each other. In addition, differences between the predicted and observed risks were plotted to compare the model performances for different categories of each subgroup. The Brier scores and calibration curves were compared across population subgroups. The survival Brier scores were not calculated for the QStroke model, as the IPCW weighted estimates did not match the Aalen-Johanssen estimates.

### 2.6 Algorithmic Fairness in Discrimination

For discrimination comparisons, we calculated key prediction indicators using the four main metrics of the confusion matrix for binary predictions: true positives (TP), true negatives (TN), false positives (FP), and false negatives (FN). Specifically, we quantified overall and group-specific true positive rates (TPR or sensitivity, TP/TP+FN), true negative rates (TNR, specificity, TN/FP+TN), false positive rates (FPR, 1-specificity), and false negative rates (FNR, 1-sensitivity). We considered prediction parity with regard to each metric when values were close to each other to a prespecified degree (80% of the values in the assumed privileged group). The goal of equalizing errors is to further prevent minoritized groups from disproportionately bearing the negative impacts caused by errors and thus perpetuating cycles of inequality [35]. We chose these metrics of discrimination as they are commonly used and widely understood in clinical settings for binary prediction problems. All values for the confusion matrices are weighted using the weights derived using IPCW.

While for calibration we considered time-to-event statistics, for discrimination, we considered binary outcomes. The thresholds used to dichotomize individual risks were chosen from QPrediction guidelines for clinical use. These thresholds hold clinical significance, as individuals surpassing these are likely to receive additional healthcare resources or be prioritized for preventive action. For QRISK3, NICE recommends offering statins for people with a score of 10% or more for the prevention of cardiovascular disease [3]. For QDiabetes, the NHS Health Check Best Practice Guideline 2019 considers people with a QDiabetes score of 5.6% or more to be at high risk of diabetes [21]. As QStroke has not been recommended by any national guidelines, we omitted QStroke from the discrimination analysis as the dichotomization of the risk scores could not be clinically reasoned, and the impact of the results would not be interpretable in a clinical setting.

For assessing the cost of misclassification, we calculated net benefits for QRISK3 and QDiabetes for each demographic subgroup. Net benefit (NB) is the expected number of true-positive treatments with a subtracted weighted penalty for unnecessary treatment per patient [36]. The weights for the unnecessary treatments are calculated using the thresholds from QPrediction guidelines for clinical use mentioned earlier. The calculated net benefits are compared to scenarios where every individual is treated (TrA) and where no one is treated (TrN) to determine the performance of the model. In addition, we calculated the net interventions avoided (NI avoided) to assess whether using the QPrediction models helps reduce unnecessary treatments.

This study was undertaken following the TRIPOD+AI guidelines [13] (see checklist in Table S10).

### 2.7 Software and Code

R was used for data cleaning and analysis, and version control was done using Git. The codes are available at GitHub: https://github.com/ihuynhi/qpred_uk_biobank. No study protocol was prepared beforehand.

## 3 Results

In total, 502,131 UK Biobank participants were considered for inclusion in this study. After applying model-specific exclusion criteria, we analyzed data from 405,855 participants for QRISK3, 458,700 participants for QDiabetes, and 490,471 participants for QStroke. The median follow-up times were 13.60 years for QRISK3, 13.62 years for QDiabetes, and 13.63 years for QStroke. Baseline characteristics of the model-specific cohorts can be found in the supplemental tables (Table S4.1-S4.3). The baseline cohort (QRISK3) had a mean age of 55.5 (SD = 8.1), and was 57.9% female and 94.2% self-reported their ethnicity as White.

**Table 2:**
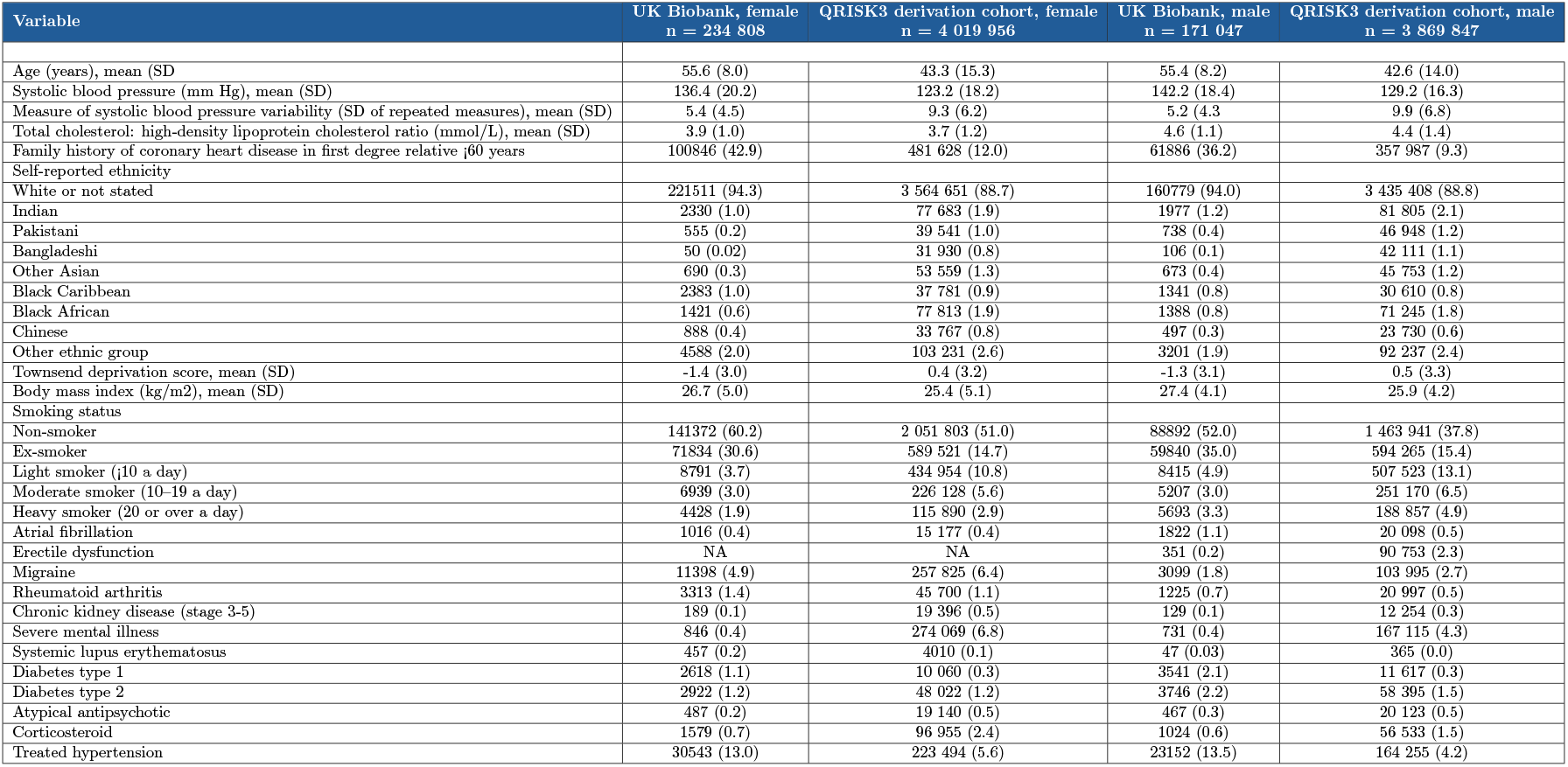
Baseline charasteristics of UK Biobank cohort used for validating QRISK3 as well as the QResearch cohort used to derive the models. The tables of baseline charasteristics for QDiabetes and QStroke can be found from the supplements.

### 3.1 Calibration

All models showed systematic overprediction of risks for the UK Biobank validation cohorts based on the calibration plots (Figure S8.1) and survival Brier scores (Table S9.1, S9.2), with only the degree of overprediction varying between the models. As the survival Brier scoe

The survival Brier scores ranged between 0.59, the best calibration across our analysis demonstrated for the oldest age group (65-75 year-olds), to 0.93, the poorest calibration across our analysis demonstrated for the youngest age group (35-45 year-olds). When comparing the differences between observed and predicted risks, only minor differences could be seen across subgroups. For example, Figure 2 shows the risk differences based on the highest education level, and the overprediction was the highest in QRISK3 for participants with a college or university degree by a small margin. For QDiabetes and QStroke, the differences appeared negligible. Similar patterns could be observed for other subgroups.

**Figure 2:**
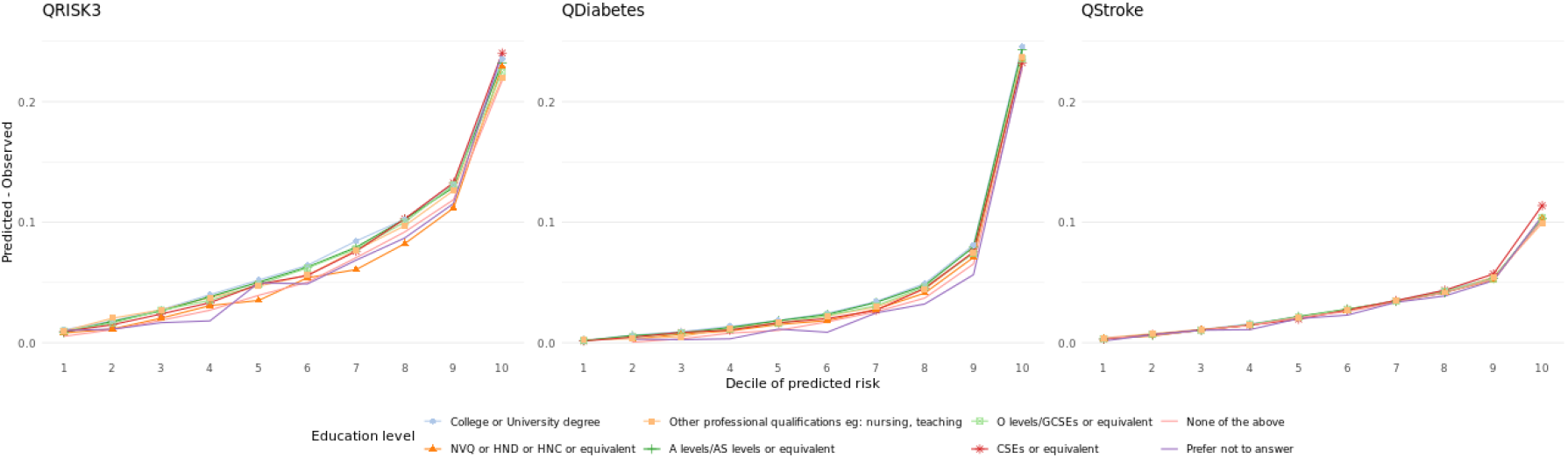
Difference of predicted and observed values based on education level for QRISK3, QDiabetes, and QStroke.

### 3.2 Discrimination

For ethnicity, the TNR and FNR values were lower for participants who self-reported as White in QRISK3 compared with any other ethnicity, apart from Asian and Asian British, although the sample size of the latter group was small. That is, the participants who self-reported as White were less likely to remain undiagnosed, both in case of them truly having no CVD (TNR) and in case they truly having CVD (FNR). In other words, White participants were in overall more likely to be diagnosed with CVD: they were subjects to both overestimation (FPR) and accurate diagnosis of CVD (TPR). Assuming White to be the privileged group and considering sample size, prediction parity was not reached in any of the subgroups in terms of TPR and FPR. In terms of education level, average household income before taxation, Townsend deprivation score, and index of multiple deprivation, the TNR and FNR seemed generally lower in the lower levels of education, income, and deprivation score. Here, prediction parity was not achieved in FNR and TNR, but was achieved in FPR and TPR for all of the variables mentioned apart from income. For immigration status, there were no major differences in the discrimination metrics, and prediction parity was achieved. For QDiabetes, the trends were similar, apart from ethnicity, where we could observe the opposite pattern as shown for QRISK3.

Apart from the age group 35-45, all the net interventions avoided were negative (Table S9.3) which translates to over-treatment and increase of unnecessary interventions, corollary to the overprediction observed in calibration and FPRs. Comparing NB and TrN for different ethnicities in QRISK3, the NBs were slightly higher than the TrN values for participants who self-reported as Black or Black British, Asian or Asian British, Do not know, and Prefer not to answer. For all the ethnicities, the TrA values were higher than the NB values. That is, deciding to treat everyone seems to yield less unnecessary interventions compared to assigning interventions based on the QRISK3.

## 4 Discussion

In our analysis, all models demonstrated systematic overprediction overall and across all evaluated subgroups defined by protected attributes, such as sex, age, ethnicity, immigrant status, income, education, and area-level deprivation. When comparing predictive performance across subgroups, we observed substantial variation in discrimination metrics across certain groups. However, calibration was relatively consistent, with no large differences observed across subgroups except for a few comparisons.

Our analysis allowed us to quantify average predictive metrics across population subgroups in the UK; by comparing these metrics, we were able to evaluate aspects of algorithmic fairness. While a wide range of algorithmic group fairness metrics have been proposed to assess parity in predictive performance, their interpretation depends heavily on the context and objectives of the study. Here, we assessed group fairness along two distinct routes: differences in prediction error and differences in model calibration across subgroups. Together, these measures provide a more holistic picture of performance disparities.

**Table 3:**
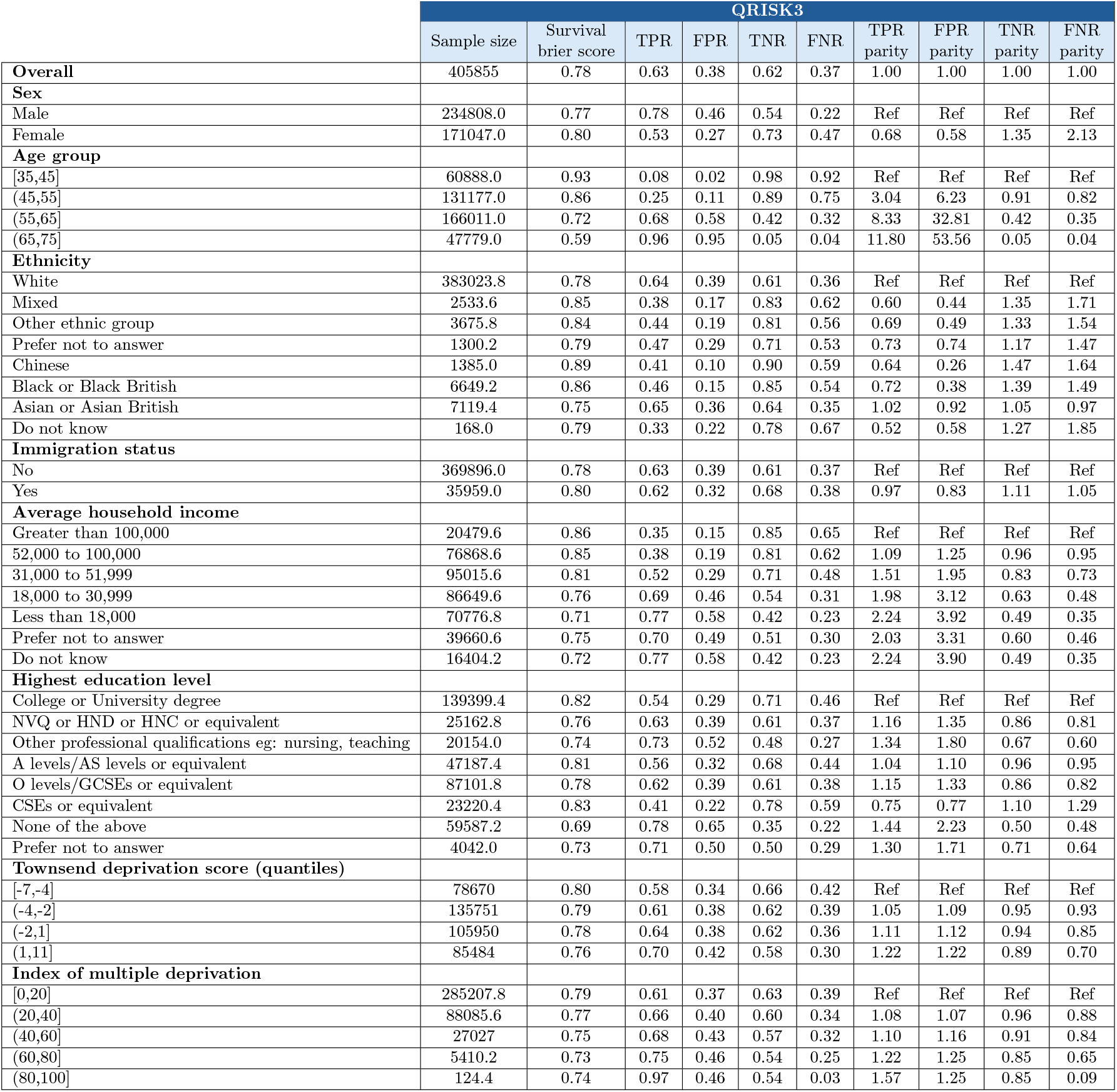
Table of sample sizes, survival Brier scores, confusion matrices, and rate parities for demographic subgroups for QRISK3.

**Table 4:**
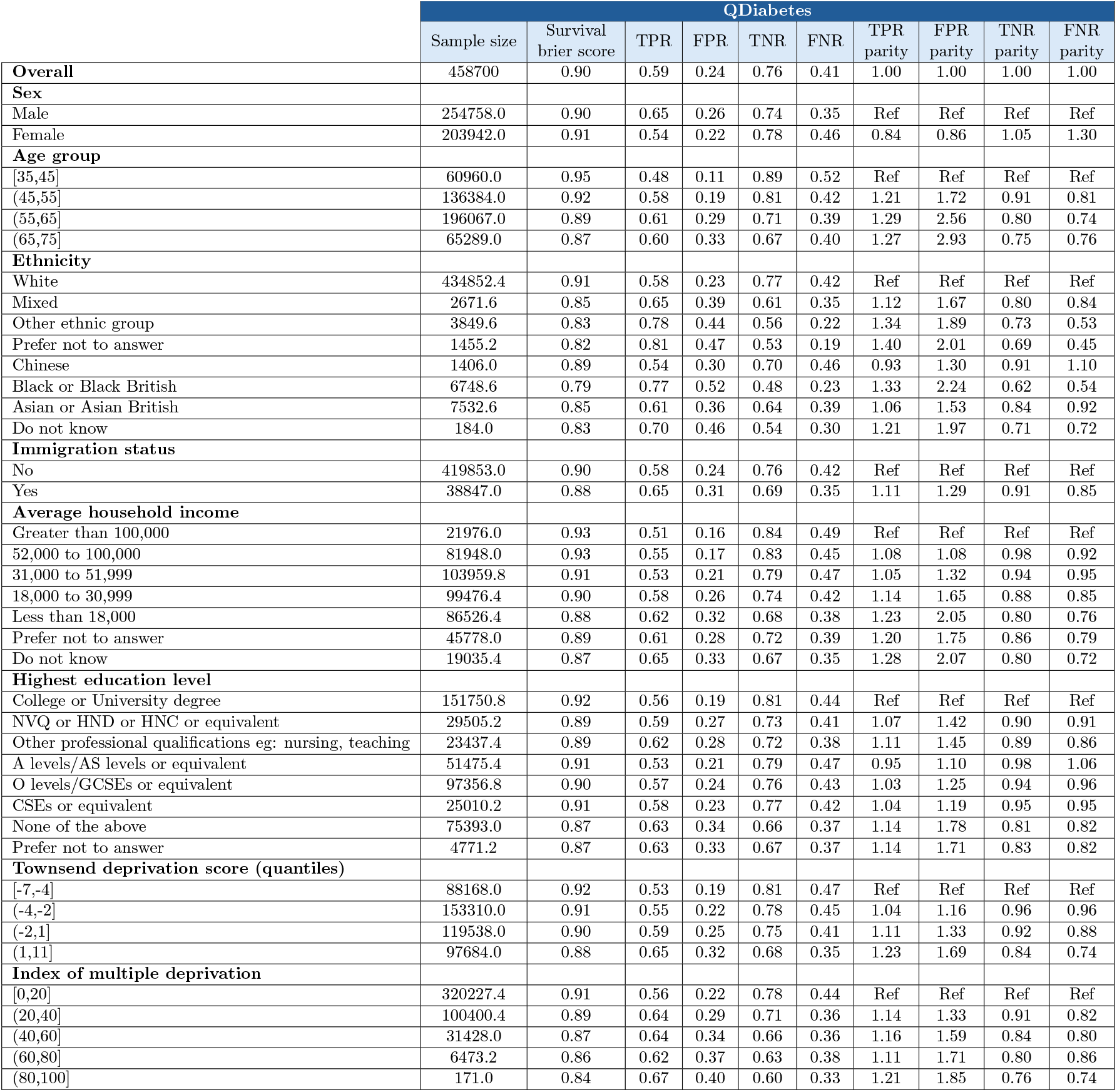
Table of sample sizes, survival Brier scores, confusion matrices, and rate parities for demographic subgroups for QDiabetes.

Models trained on biased datasets or without explicitly addressing data biases are likely to perpetuate and amplify existing inequities in clinical decision-making [6][37][8]. Prior research suggests that incorporating social determinants of health (SDOH) into clinical prognostic prediction models can enhance performance for specific population subgroups [38]. QPrediction models integrate certain SDOH, such as area-level deprivation, measured using the Townsend deprivation score, and self-reported ethnicity. However, the representation of these variables is often overly simplistic and unbalanced. These simplistic categorizations obscure the complexity of social and cultural identities, raising concerns about whether such representations meaningfully enhance fairness, even when explicitly included in risk models. Despite these limitations, we hypothesize that the inclusion of ethnicity and deprivation in the QPrediction models may have contributed to more consistent calibration across subgroups defined by protected attributes, which are themselves often correlated, such as ethnicity and socioeconomic status. This could partially explain why we observed relatively small differences in calibration curves across subgroups. Furthermore, the original QPrediction derivation cohorts included a higher proportion of participants from ethnic minority backgrounds compared to our validation cohorts drawn from the UK Biobank. This greater diversity in the development datasets may have supported more equitable model performance, particularly in terms of calibration, across population subgroups.

While calibration was relatively similar across subgroups, it should be emphasized that we observed overprediction for all subgroups in our analysis. Given that QPrediction models are applied in the clinic, one explanation for the observed overprediction is that some participants may have previously received preventive care as a result of QPrediction risk scores, systematically lowering the actual event rates. Similarly, the prevented adverse outcomes have most likely caused the increase in FPRs as well as inflated the net interventions avoided.

Despite similar calibration across population subgroups, we observed substantial disparities in FPR and FNR. Among women, for example, the FPR for QRISK3 was approximately 40% lower than that for men. False negatives, on the other hand, were twice as high in women compared to men. Similar, though less pronounced, patterns were observed in the QDiabetes model. These results suggest that women were less likely to be incorrectly flagged as high-risk, but more likely to be incorrectly assessed as low-risk. This phenomenon may result in the undertreatment of women, as true cases may go unrecognized.

For QRISK3, we found that older individuals, those with lower income levels, and those with lower educational attainment had higher FPRs and lower FNRs. These trends were largely mirrored in QDiabetes. For income, disparities in FPR larger than 50% were common in the observed data. Patterns for ethnicity were different between QRISK3 and QDiabetes. In QRISK3, individuals who self-identified as non-White experienced higher FNRs and lower FPRs, meaning the model was more likely to miss true cases among minority groups. Conversely, QDiabetes showed the opposite pattern: higher FPRs and lower FNRs among non-White groups, suggesting a tendency to overpredict risk in these populations, similar to another study predicting undiagnosed hypertension [39]. When analyzing immigration status, QRISK3 showed parity, with no major differences in error rates between immigrants and non-immigrants. However, in QDiabetes, immigrants had approximately 30% higher FPR, suggesting a greater likelihood of being incorrectly flagged as high-risk.

For QRISK3, both FPR and FNR were relatively stable across quintiles of the Townsend deprivation score, suggesting that including this variable in the model may have helped mitigate socioeconomic disparities in classification errors. In contrast, QDiabetes showed similar FNRs across deprivation quintiles, but FPRs were higher among individuals with greater deprivation, indicating potential over-prediction in more socioeconomically disadvantaged groups.

The observation that calibration was consistent across subgroups, while FPR and FNR differed substantially, is not surprising and is well-explained by the fairness impossibility theorem [40][41]. This theorem demonstrates that in the presence of unequal base rates (i.e., different outcome prevalences across groups), it is mathematically impossible to simultaneously satisfy calibration and particular discrimination metrics. In other words, ensuring that predicted probabilities are accurate for each group will inherently lead to disparities in error rates. A well-known example of this trade-off can be observed in ProPublica’s investigation of the COMPAS risk assessment tool, where the model was found to be similarly calibrated for Black and White defendants but exhibited stark imbalances in FPR and FNR across racial groups [42].

We highlight that while expanding datasets to be more representative of population subgroups and incorporating SDOH into model development can improve predictive performance and potentially advance equity [10], such technical solutions are ultimately limited. Algorithmic bias and unfairness [43] often reflect underlying structural inequities that are embedded and also extend beyond the healthcare system. Even when models like the QPrediction algorithms integrate variables such as area-level deprivation or self-reported ethnicity, the models are typically deployed in a clinical setting to guide individual-level decision making, such as initiating preventive medications. This can inadvertently shift responsibility (and perhaps blame) for health outcomes onto patients, reinforcing individualistic explanations for disease while obscuring the structural and societal forces, such as racism, poverty, and unequal access to care, that shape health risks in the first place [44].

The validity of our results relies on the assumption that the variables and outcomes in the UK Biobank accurately reflect real-world events and characteristics. However, several forms of data bias must be considered; rather than enlisting these as analytical limitations, as is usually done in scientific literature, we present these points as key discussion points here. Outcome ascertainment in the Biobank relies on electronic health records and linked data, which may under-capture certain conditions, particularly in groups with lower healthcare utilization [45]. Moreover, variables were measured only at baseline, while many social and economic conditions, such as employment status, neighborhood deprivation, and even self-identified ethnicity, can shift over time [46]. The long follow-up periods for events mean that important social changes occurring between baseline and event onset are not captured, potentially affecting our analysis. Furthermore, self-reported data is prone to systematic reporting errors, which might further decrease the validity of the findings [47].

Ethnicity, in particular, poses complex challenges [48][49], and while the incorporation of these constructs has been shown to reduce algorithmic bias [50], their use is still controversial for multiple reasons. This study, like many others, relied on self-reported ethnic background. While self-identification is a valid and commonly used method, it is ambiguous and subject to both individual interpretation and broader social dynamics. Research has shown that self-assigned and socially-assigned race or ethnicity can diverge significantly [51], with meaningful implications for health outcomes and discrimination. The categories available in the UK Biobank and the QPrediction models are coarse and rooted in geographic or national labels. For instance, broad racial labels like “White” are categorized alongside specific nationalities such as “Indian” and “Pakistani”, and national identities such as “British” and “Irish” are grouped at the same level as “African”, which encompasses an entire continent that is the most genetically diverse in the world [52]. While the latter phenomenon is likely due to practical considerations related to sample size, it is undeniable that grouping diverse populations under umbrella labels conceals meaningful heterogeneity in culture, biology, and lived experience. It also remains unclear whether these categories are intended to reflect shared genetics, cultural heritage, or shared experiences of being racialized, and how histories of colonization, migration, or social stratification influence these identities. Due to the crudeness of the ethnic categories, further research is needed to assess whether the inclusion of ethnicity in the models is actually useful for minoritized groups.

In the EU alone, health inequalities account for 20% of total healthcare costs and related welfare losses amount to nearly 1 trillion EUR per year [53]. More importantly, the amount of avoidable health inequalities caused by structural inequities [54] cannot be adequately quantified. Despite the potential for reducing health(care) inequalities through the use of bias mitigation techniques, we argue that it is unlikely that algorithmic bias mitigation strategies, alone, would be adequate in addressing the structural drivers of health disparities rooted in racial capitalism, colonialism, and patriarchy [55][56][57]. Closing the inequity gap is a self-evident moral imperative: as a WHO report from 2008 puts it, currently observed health inequalities linked to social disadvantage “should simply never happen” [54].

## Data Availability

Data can be applied to via https://www.ukbiobank.ac.uk. This study was conducted based on the UK Biobank cohort study under application number 103919.

https://github.com/ihuynhi/qpred_uk_biobank

## 5 Acknowledgements

No generative AI tools have been used in the creation of this article.

## 8 CRediT Author Statement

The contributors are listed alphabetically.

Conceptualization: IH, DU, TVV

Methodology: IH, AK, TLN, DU, TVV

Software: IH

Validation: IH

Formal analysis: IH

Investigation: IH, TVV

Resources: TVV

Data Curation: IH

Writing - Original Draft: IH, DU, TVV

Writing - Review & Editing: IH, AK, TLN, DU, TVV

Visualization: IH

Supervision: TLN, DU, TVV

Project administration: TVV

Funding acquisition: TVV

