## supplementary_material for "Algorithmic Fairness of QPrediction Cardiometabolic Risk Prediction Models"

### 2 Variable matching

#### 2.1 QRISK3

##### 2.1.1 Table of variable names in QRISK3 and their equivalents in UK Biobank

| Variable name in QRISK3 | Variable name in UK Biobank | Notes |
|---|---|---|
| age | 34: Year of birth<br>52: Month of birth<br>53: Date of attending assessment centre | Age calculated between year and month of the birth (the date set to the first) and the time of the baseline assesment |
| atrial fibrillation | 41270: Diagnoses - ICD10 (I48)<br>41280: Date of first in-patient diagnosis - ICD10<br><br>41271: Diagnoses - ICD9 (4273)<br>41281: Date of first in-patient diagnosis - ICD9<br><br>41272: Operative procedures - OPCS4 (K62.2, K62.3)<br>41282: Date of first operative procedure - OPCS4<br><br>20002: Non-Cancer Illness Code (1471, 1483) | |
| body mass index | 21001: Body mass index (BMI) | |
| chronic kidney disease (stage 3-5) | 41270: Diagnoses - ICD10 (N18.3, N18.4, N18.5)<br>41280: Date of first in-patient diagnosis - ICD10<br><br>41271: Diagnoses - ICD9 (585, 5959, 5853, 5855, 5810, 5820, 5900, V420, V451)<br>41281: Date of first in-patient diagnosis - ICD9<br><br>20002: Non-Cancer Illness Code (1192, 1519, 1609) | Added 585, 5959 |
| diagnosis or treatment for erectile dysfunction | 41270: Diagnoses - ICD10 (N48.4)<br>41280: Date of first in-patient diagnosis - ICD10<br><br>41271: Diagnoses - ICD9 (60784)<br>41281: Date of first in-patient diagnosis - ICD9<br><br>20002: Non-Cancer Illness Code (1518)<br><br>20003: Treatment/medication code (1141168936, 1141168948, 1141168944, 1141168946, 1140869100, 1140883010) | |
| ethnicity (self-reported) | 21000: Ethnic background (All instances) | See QRISK3_categories_coding |
| family history of coronary heart disease in first degree relative aged <60 years | 20107: Illnesses of father (= heart disease)<br>20110: Illnesses of mother (= heart disease)<br>20111: Illnesses of siblings (= heart disease) | Not available for coronary heart disease and no age restrictions. Assumed that CHD included in "heart disease" and that relative < 60 at the time of diagnosis |
| migraine | 41270: Diagnoses - ICD10 (G43, G44.0, N94.3)<br>41280: Date of first in-patient diagnosis - ICD10<br><br>41271: Diagnoses - ICD9 (346)<br>41281: Date of first in-patient diagnosis - ICD9<br><br>20002: Non-Cancer Illness Code (1265)<br><br>120016: Ever had migraine (= yes) | |
| rheumatoid arthritis | 41270: Diagnoses - ICD10 (M05, M06)<br>41280: Date of first in-patient diagnosis - ICD10<br><br>41271: Diagnoses - ICD9 (714)<br>41281: Date of first in-patient diagnosis - ICD9<br><br>20002: Non-Cancer Illness Code (1464) | |
| severe mental illness<br>(schizophrenia, bipolar disorder and moderate/severe depression) | 41270: Diagnoses - ICD10 (F03, F06.8, F09, F20, F22, F23*, F25.9, F28, F29, F31, F39, F53, F33*)<br>41280: Date of first in-patient diagnosis - ICD10<br><br>41271: Diagnoses - ICD9 (295, 298, 296)<br>41281: Date of first in-patient diagnosis - ICD9<br><br>20002: Non-Cancer Illness Code (1289, 1291) | |
| sex | 31: Sex | |
| smoking status | 20116: Smoking status (all instances)<br>22506: Tobacco smoking<br>22508: Amount of tobacco currently smoked<br>1239: Current tobacco smoking (all instances)<br>3456: Number of cigarettes currently smoked daily (current cigarette smokers) (all instances) | |
| systemic lupus erythematosus | 41270: Diagnoses - ICD10 (M32)<br>41280: Date of first in-patient diagnosis - ICD10<br><br>41271: Diagnoses - ICD9 (7100)<br>41281: Date of first in-patient diagnosis - ICD9<br><br>20002: Non-Cancer Illness Code (1381) | |
| systolic blood pressure | 4080: Systolic blood pressure, automated reading<br>93: Systolic blood pressure, manual reading | A manual sphygmometer was used if the standard automated device could not be employed. |
| systolic blood pressure variability (standard deviation of at least two most recent systolic blood pressure readings (mmHg)) | 93: Systolic blood pressure, manual reading<br>4080: Systolic blood pressure, automated reading | A manual sphygmometer was used if the standard automated device could not be employed. |
| total cholesterol (cholesterol/HDL ratio) | 30690: Cholesterol (All instances, available for the first two)<br>30760: HDL cholesterol (All instances, available for the first two) | |
| Townsend deprivation score | 22189: Townsend deprivation index at recruitment | |
| type 1 diabetes | 41270: Diagnoses - ICD10 (E10, O24.0)<br>41280: Date of first in-patient diagnosis - ICD10<br><br>41271: Diagnoses - ICD9 (25001, 25011, 25021, 25031, 25041, 25051, 25061, 25071, 25081, 25091, 25003, 25013, 25023, 25033, 25043, 25053, 25063, 25073, 25083, 25093)<br>41281: Date of first in-patient diagnosis - ICD9<br><br>20002: Non-Cancer Illness Code (1222) | |
| type 2 diabetes | 41270: Diagnoses - ICD10 (E11, O24.1)<br>41280: Date of first in-patient diagnosis - ICD10<br><br>41271: Diagnoses - ICD9 (25000, 25010, 25020, 25030, 25040, 25050, 25060, 25070, 25080, 25090, 25002, 25012, 25022, 25032, 25042, 25052, 25062, 25072, 25082, 25092)<br>41281: Date of first in-patient diagnosis - ICD9<br><br>20002: Non-Cancer Illness Code (1223, 1220)<br><br>20003: Treatment/medication code (1140868902, 1140874646, 1140874674, 1140874718, 1140874744, 1140883066, 1140884600, 1141152590, 1141157284, 1141168660, 1141171646, 1141173882, 1141189090)<br><br>2443: Diabetes diagnosed by doctor (All instances)<br><br>30750: Glycated haemoglobin (HbA1c) (>= 48) | |

#### 2.1.2 Table of medication variables names in QRISK3 and their equivalents in medication codes in UK Biobank

| QRISK3 | UK Biobank | | |
|---|---|---|---|
| Variable name | Variable name | Codes | Notes |
| atypical antipsychotics (Including amisulpride, aripiprazole, clozapine, lurasidone, olanzapine, paliperidone, quetiapine, risperidone, sertindole, or zotepine) | 20003: Treatment/medication code | 1140867420, 1140867444, 1140927956, 1140928916, 1141152848, 1141153490, 1141169714, 1141195974 | |
| regular use of steroid tablets (including oral or parenteral prednisolone, betamethasone, cortisone, depo-medrone, dexamethasone, deflazacort, efcortesol, hydrocortisone, methylprednisolone, or triamcinolone) | 20003: Treatment/medication code | 1140874790, 1140874816, 1140874896.00, 1140874930, 1140874976, 1141145782, 1141173346 | British National Formulary (BNF) chapter 6.3.2 |
| treated hypertension | 6177: Medication for cholesterol, blood pressure or diabetes (All instances) | Blood pressure medication | |
| treated hypertension | 6153: Medication for cholesterol, blood pressure, diabetes, or take exogenous hormones | Blood pressure medication | |
| treated hypertension | 20003: Treatment/medication code | 1140860192, 1140860292, 1140860696, 1140860728, 1140860750, 1140860806, 1140860882, 1140860904, 1140861088, 1140861190, 1140861276, 1140866072, 1140866078, 1140866090, 1140866102, 1140866108, 1140866122, 1140866138, 1140866156, 1140866162, 1140866724, 1140866738, 1140868618, 1140872568, 1140874706, 1140874744, 1140875808, 1140879758, 1140879760, 1140879762, 1140879802, 1140879806, 1140879810, 1140879818, 1140879822, 1140879826, 1140879830, 1140879834, 1140879842, 1140879866, 1140884298, 1140888552, 1140888556, 1140888560, 1140888646, 1140909706, 1140910442, 1140910614, 1140916356, 1140923272, 1140923336, 1140923404, 1140923712, 1140926778, 1140928226, 1141145660, 1141146126, 1141152998, 1141153026, 1141164276, 1141165470, 1141166006, 1141169516, 1141171336, 1141180592, 1141180772, 1141180778, 1141184722, 1141193282, 1141194794, 1141194810 | |

#### 2.1.3 Table of recategorization of UK Biobank variables to match the categories in QRISK3 variables

| QRISK3 | | UK Biobank | | |
|---|---|---|---|---|
| Variable name | Categories | Variable name | Categories | Notes |
| ethnicity (self-reported) | White or not stated, Indian, Pakistani, Bangdaleshi, Other Asian, Black Caribbean, Black African, Chinese, Other ethnic group | 21000: Ethnic background | **White or not stated**<br>White (British, Irish, Any other white background), Do not know, Prefer not to answer<br>**Indian**<br>Indian<br>**Pakistani**<br>Pakistani<br>**Bangdaleshi**<br>Bangladeshi<br>**Other Asian**<br>Any other Asian background<br>**Black Caribbean**<br>Caribbean<br>**Black African**<br>African<br>**Chinese**<br>Chinese<br>**Other ethnic group**<br>Mixed (White and Black Caribbean, White and Black African, White and Asian, Any other mixed background), Any other Black background, Other ethnic group | White (British, Irish, Any other white background), Mixed (White and Black Caribbean, White and Black African, White and Asian, Any other mixed background), Asian or Asian British (Indian, Pakistani, Bangladeshi, Any other Asian background), Black or Black British (Caribbean, African, Any other Black background), Chinese, Other ethnic group, Do not know, Prefer not to answer |
| smoking status | non-smoker, ex-smoker, light smoker (less than 10), moderate smoker (10 to 19), heavy smoker (over 20) | 20116: Smoking status<br>22506: Tobacco smoking<br>22508: Amount of tobacco currently smoked<br>1239: Current tobacco smoking<br>3456: Number of cigarettes currently smoked daily (current cigarette smokers) | **non-smoker:**<br>20116: Smoking status: Never<br>1239: Current tobacco smoking: No<br>**ex-smoker:**<br>20116: Smoking status: Previous<br>**light smoker (less than 10), moderate smoker (10 to 19), heavy smoker (over 20):**<br>1239: Only occasionally<br>3456: Number of cigarettes currently smoked daily (current cigarette smokers) | If current smoker based on 1239, the number of cigarettes smoked taken from 3456. If 3456 is unavailable, then separate category "smoker". |

#### 2.2 QDiabetes

##### 2.2.1 Table of variable names in QDiabetes and their equivalents in UK Biobank

| Variable name in QDiabetes | Variable name in UK Biobank | Notes |
|---|---|---|
| age | 34: Year of birth<br>52: Month of birth<br>53: Date of attending assessment centre | Age calculated between year and month of the birth (the date set to the first) and the time of the baseline assesment |
| body mass index | 21001: Body mass index (BMI) | |
| family history of diabetes in a first degree relative | 20107: Illnesses of father (= Diabetes)<br>20110: Illnesses of mother (= Diabetes)<br>20111: Illnesses of siblings (=Diabetes) | |
| fasting blood glucose | 23470: Glucose<br>Fasting time from<br>74: Fasting time<br><br>30740: Glucose<br>Fasting time from<br>74: Fasting time | 23470_ Glucose from Nightingale Health data. Biomarker group: Glycolysis related metabolites<br>30740: Measured by hexokinase analysis on a Beckman Coulter AU5800 |
| gestational diabetes | 41270: Diagnoses - ICD10 (O24)<br>41280: Date of first in-patient diagnosis - ICD10<br><br>41271: Diagnoses - ICD9 (648)<br>41281: Date of first in-patient diagnosis - ICD9<br><br>4041: Gestational diabetes only (= Yes) | |
| HbA1c | 30750: Glycated haemoglobin (HbA1c) | |
| learning disability | 41270: Diagnoses - ICD10 (F81.0)<br>41280: Date of first in-patient diagnosis - ICD10<br><br>41271: Diagnoses - ICD9 (31500, 31502, 31509, 3152, V40.0)<br>41281: Date of first in-patient diagnosis - ICD9 | |
| disorder or schizophrenia | 41270: Diagnoses - ICD10 (F20, F22, F23*, F25.9, F28, F29, F31)<br>41280: Date of first in-patient diagnosis - ICD10<br><br>41271: Diagnoses - ICD9 (295, 298, 296)<br>41281: Date of first in-patient diagnosis - ICD9<br><br>20002: Non-Cancer Illness Code (1291) | |
| peripheral vascular disease | 41270: Diagnoses - ICD10 (I70-I79)<br>41280: Date of first in-patient diagnosis - ICD10<br><br>41271: Diagnoses - ICD9 (2506, 4400, 4402, 4431, 4438, 4439)<br>41281: Date of first in-patient diagnosis - ICD9<br><br>41272: Operative procedures - OPCS4 (L21.6, L51.3, L51.6, L51.8, L52.1, L52.2, L54.1, L54.4, L54.8, L59.1, L59.2, L59.3, L59.4, L59.5, L59.6, L59.7, L59.8, L60.1, L60.2, L63.1, L63.5, L63.9, L66.7)<br>41282: Date of first operative procedure - OPCS4<br><br>20002: Non-Cancer Illness Code (1067, 1088)<br><br>20004: Operation code, self-reported (1102, 1108, 1440, X09.3, X09.4, X09.5)<br><br>20003: Treatment/medication code (1141181150, 1141181154) | Disease codes from<br>https://stanfordhealthcare.org/medical-conditions/blood-heart-circulation/peripheral-vascular-disease/types.html |
| polycystic ovary syndrome | 41270: Diagnoses - ICD10 (E28.2)<br>41280: Date of first in-patient diagnosis - ICD10<br><br>41271: Diagnoses - ICD9 (2564)<br>41281: Date of first in-patient diagnosis - ICD9 | |
| previous heart attack, angina, stroke or transient ischemic attack | 6150: Vascular/heart problems diagnosed by doctor (= Heart attack, Angina, Stroke)<br><br>41270: Diagnoses - ICD10 (I21, I20, I24, I60, I61, I63, I64, G45)<br>41280: Date of first in-patient diagnosis - ICD10<br><br>41271: Diagnoses - ICD9 (410, 411, 412, 413, 414, 429.79, 434, 435, 436)<br>41281: Date of first in-patient diagnosis - ICD9<br><br>41272: Operative procedures - OPCS4 (K40, K41, K42, K43, K44, K45, K46, K47.1, K49, K50, K75)<br>41282: Date of first operative procedure - OPCS4<br><br>20004: Operation code, self-reported (1074, 1075, 1081, 1082, 1086, 1491, 1583) | |
| sex | 31: Sex | |
| Townsend deprivation score | 22189: Townsend deprivation index at recruitment | |

#### 2.2.2 Table of medication variables names in QDiabetes and their equivalents in medication codes in UK Biobank

| Variable name | Variable name | Codes | Notes |
|---|---|---|---|
| atypical antipsychotics (Including amisulpride, aripiprazole, clozapine, lurasidone, olanzapine, paliperidone, quetiapine, risperidone, sertindole, or zotepine) | 20003: Treatment/medication code | 1140867420, 1140867444, 1140927956, 1140928916, 1141152848, 1141153490, 1141169714, 1141195974 | |
| treated hypertension | 6177: Medication for cholesterol, blood pressure or diabetes (All instances) | Blood pressure medication | |
| treated hypertension | 6153: Medication for cholesterol, blood pressure, diabetes, or take exogenous hormones | Blood pressure medication | |
| treated hypertension | 20003: Treatment/medication code | 1140860192, 1140860292, 1140860696, 1140860728, 1140860750, 1140860806, 1140860882, 1140860904, 1140861088, 1140861190, 1140861276, 1140866072, 1140866078, 1140866090, 1140866102, 1140866108, 1140866122, 1140866138, 1140866156, 1140866162, 1140866724, 1140866738, 1140868618, 1140872568, 1140874706, 1140874744, 1140875808, 1140879758, 1140879760, 1140879762, 1140879802, 1140879806, 1140879810, 1140879818, 1140879822, 1140879826, 1140879830, 1140879834, 1140879842, 1140879866, 1140884298, 1140888552, 1140888556, 1140888560, 1140888646, 1140909706, 1140910442, 1140910614, 1140916356, 1140923272, 1140923336, 1140923404, 1140923712, 1140926778, 1140928226, 1141145660, 1141146126, 1141152998, 1141153026, 1141164276, 1141165470, 1141166006, 1141169516, 1141171336, 1141180592, 1141180772, 1141180778, 1141184722, 1141193282, 1141194794, 1141194810 | |
| regular use of steroid tablets (including oral or parenteral prednisolone, betamethasone, cortisone, depo-medrone, dexamethasone, deflazacort, efcortesol, hydrocortisone, methylprednisolone, or triamcinolone) | 20003: Treatment/medication code | 1140874790, 1140874816, 1140874896.00, 1140874930, 1140874976, 1141145782, 1141173346 | British National Formulary (BNF) chapter 6.3.2 |
| use of statins | 20003: Treatment/medication code | 1140861958, 1141146234, 1140888648, 1141192410, 1140888594 | |

#### 2.2.3 Table of recategorization of UK Biobank variables to match the categories in QDiabetes variables

| QDiabetes | | UK Biobank | | |
|---|---|---|---|---|
| Variable name | Categories | Variable name | Categories | Notes |
| ethnicity (self-reported) | White or not stated, Indian, Pakistani, Bangdaleshi, Other Asian, Black Caribbean, Black African, Chinese, Other ethnic group | 21000: Ethnic background | **White or not stated**<br>White (British, Irish, Any other white background), Do not know, Prefer not to answer<br>**Indian**<br>Indian<br>**Pakistani**<br>Pakistani<br>**Bangdaleshi**<br>Bangladeshi<br>**Other Asian**<br>Any other Asian background<br>**Black Caribbean**<br>Caribbean<br>**Black African**<br>African<br>**Chinese**<br>Chinese<br>**Other ethnic group**<br>Mixed (White and Black Caribbean, White and Black African, White and Asian, Any other mixed background), Any other Black background, Other ethnic group | White (British, Irish, Any other white background), Mixed (White and Black Caribbean, White and Black African, White and Asian, Any other mixed background), Asian or Asian British (Indian, Pakistani, Bangladeshi, Any other Asian background), Black or Black British (Caribbean, African, Any other Black background), Chinese, Other ethnic group, Do not know, Prefer not to answer |
| smoking status | non-smoker, ex-smoker, light smoker (less than 10), moderate smoker (10 to 19), heavy smoker (over 20) | 20116: Smoking status<br>1239: Current tobacco smoking<br>3456: Number of cigarettes currently smoked daily (current cigarette smokers) | **non-smoker:**<br>20116: Smoking status: Never<br>1239: Current tobacco smoking: No<br>**ex-smoker:**<br>20116: Smoking status: Previous<br>**light smoker (less than 10), moderate smoker (10 to 19), heavy smoker (over 20):**<br>1239: Only occasionally<br>3456: Number of cigarettes currently smoked daily (current cigarette smokers) | If current smoker based on 1239, the number of cigarettes smoked taken from 3456. If 3456 is unavailable, then separate category "smoker". |

### 2.3 QStroke

#### 2.3.1 Table of variable names in QStroke and their equivalents in UK Biobank

| Variable name in QStroke | Variable name in UK Biobank | Notes |
|---|---|---|
| age | 34: Year of birth<br>52: Month of birth<br>53: Date of attending assessment centre | Age calculated between year and month of the birth (the date set to the first) and the time of the baseline assesment |
| atrial fibrillation | 41270: Diagnoses - ICD10 (I48)<br>41280: Date of first in-patient diagnosis - ICD10<br><br>41271: Diagnoses - ICD9 (4273)<br>41281: Date of first in-patient diagnosis - ICD9<br><br>41272: Operative procedures - OPCS4 (K62.2, K62.3)<br>41282: Date of first operative procedure - OPCS4<br><br>20002: Non-Cancer Illness Code (1471, 1483) | |
| body mass index | 21001: Body mass index (BMI) | |
| chronic kidney disease (stage 4-5) | 41270: Diagnoses - ICD10 (N18.4, N18.5)<br>41280: Date of first in-patient diagnosis - ICD10<br><br>41271: Diagnoses - ICD9 (585, 5959, 5855, 5810, 5820, 5900, V420, V451)<br>41281: Date of first in-patient diagnosis - ICD9<br><br>20002: Non-Cancer Illness Code (1192, 1519, 1609) | |
| congestive cardiac failure | 41270: Diagnoses - ICD10 (I50, I11.0, I13.0, I13.2)<br>41280: Date of first in-patient diagnosis - ICD10<br><br>41271: Diagnoses - ICD9 (428)<br>41281: Date of first in-patient diagnosis - ICD9<br><br>20002: Non-Cancer Illness Code (1076) | https://bjsm.bmj.com/content/bjsports/suppl/2024/02/28/bjsports-2023-106862.DC1/bjsports-2023-106862supp001_data_ |
| coronary heart disease | 6150: Vascular/heart problems diagnosed by doctor (= Heart attack)<br><br>41270: Diagnoses - ICD10 (I21, I22, I23, I24.1, I25.2)<br>41280: Date of first in-patient diagnosis - ICD10<br><br>41271: Diagnoses - ICD9 (410, 411, 412, 429.79, 413, 414)<br>41281: Date of first in-patient diagnosis - ICD9<br><br>41272: Operative procedures - OPCS4 (K40, K40.1, K40.2, K40.3, K40.4, K41.1, K41.2, K41.3, K41.4, K45.1, K45.2, K45.3, K45.4, K45.5, K49.1, K49.2, K49.8, K49.9, K50.2, K75.1, K75.2, K75.3, K75.4, K75.8, K75.9)<br>41282: Date of first operative procedure - OPCS4<br><br>20004: Operation code (1070, 1095)<br><br>20002: Non-Cancer Illness Code (1074, 1075) | |
| family history of coronary heart disease in first degree relative aged <60 years | 20107: Illnesses of father (= Heart disease)<br>20110: Illnesses of mother (= Heart disease)<br>20111: Illnesses of siblings (= Heart disease) | |
| heart attack or angina | 6150: Vascular/heart problems diagnosed by doctor (= Heart attack, Angina, Stroke)<br><br>41270: Diagnoses - ICD10 (I21, I20, I24, I60, I61, I63)<br>41280: Date of first in-patient diagnosis - ICD10<br><br>41271: Diagnoses - ICD9 (410, 411, 412, 413, 414, 429.79, 434, 436)<br>41281: Date of first in-patient diagnosis - ICD9<br><br>41272: Operative procedures - OPCS4 (K40, K41, K42, K43, K44, K45, K46, K47.1, K49, K50, K75)<br>41282: Date of first operative procedure - OPCS4<br><br>20004: Operation code, self-reported (1074, 1075, 1081, 1082, 1086, 1491, 1583) | |
| rheumatoid arthritis | 41270: Diagnoses - ICD10 (M05, M06)<br>41280: Date of first in-patient diagnosis - ICD10<br><br>41271: Diagnoses - ICD9 (714)<br>41281: Date of first in-patient diagnosis - ICD9<br><br>20002: Non-Cancer Illness Code (1464) | |
| sex | 31: Sex | |
| systolic blood pressure | 4080: Systolic blood pressure, automated reading<br>93: Systolic blood pressure, manual reading | |
| total cholesterol (cholesterol/HDL ratio) | 30690: Cholesterol (All instances, available for the first two)<br>30760: HDL cholesterol (All instances, available for the first two) | |
| Townsend deprivation score | 22189: Townsend deprivation index at recruitment | |
| type 1 diabetes | 41270: Diagnoses - ICD10 (E10, O24.0)<br>41280: Date of first in-patient diagnosis - ICD10<br><br>41271: Diagnoses - ICD9 (25001, 25011, 25021, 25031, 25041, 25051, 25061, 25071, 25081, 25091, 25003, 25013, 25023, 25033, 25043, 25053, 25063, 25073, 25083, 25093)<br>41281: Date of first in-patient diagnosis - ICD9<br><br>20002: Non-Cancer Illness Code (1222) | |
| type 2 diabetes | 41270: Diagnoses - ICD10 (E11, O24.1)<br>41280: Date of first in-patient diagnosis - ICD10<br><br>41271: Diagnoses - ICD9 (25000, 25010, 25020, 25030, 25040, 25050, 25060, 25070, 25080, 25090, 25002, 25012, 25022, 25032, 25042, 25052, 25062, 25072, 25082, 25092)<br>41281: Date of first in-patient diagnosis - ICD9<br><br>20002: Non-Cancer Illness Code (1223, 1220)<br><br>20003: Treatment/medication code (1140868902, 1140874646, 1140874674, 1140874718, 1140874744, 1140883066, 1140884600, 1141152590, 1141157284, 1141168660, 1141171646, 1141173882, 1141189090)<br><br>2443: Diabetes diagnosed by doctor<br><br>30750: Glycated haemoglobin (HbA1c) (>= 48) | |
| valvular heart disease | 41270: Diagnoses - ICD10 (I34, I35, I36, I37, I39.0, I39.1, I39.2, I39.3, I39.4, I05, I06, I07, I08, I09.1, I09.8)<br>41280: Date of first in-patient diagnosis - ICD10<br><br>41271: Diagnoses - ICD9 (424, 394, 395, 396, 397, 3979, 39891, 39899)<br>41281: Date of first in-patient diagnosis - ICD9<br><br>20002: Non-Cancer Illness Code (1078, 1488, 1584, 1585, 1489, 1586, 1587, 1490)<br><br>41272: Operative procedures - OPCS4 (K26, K25, K27, K30.1, K30.8, K30.9, K34.1, K34.2, K35.1, K35.5, K35.8, K35.9, Y79, Y53, Y07.2) | https://onlinecjc.ca/article/S0828-282X(23)01948-7/fulltext#appsec1<br>https://openheart.bmj.com/content/openhrt/suppl/2022/09/14/openhrt-2022-002039.DC1/openhrt-2022-002039supp001_data_supplement.pdf |

##### 2.3.2 Table of medication variables names in QStroke and their equivalents in medication codes in UK Biobank

| QStroke | UK Biobank | | |
|---|---|---|---|
| Variable name | Variable name | Codes | Notes |
| treated hypertension | 6177: Medication for cholesterol, blood pressure or diabetes | Blood pressure medication | |
| treated hypertension | 6153: Medication for cholesterol, blood pressure, diabetes, or take exogenous hormones | Blood pressure medication | |
| treated hypertension | 20003: Treatment/medication code | 1140860192, 1140860292, 1140860696, 1140860728, 1140860750, 1140860806, 1140860882, 1140860904, 1140861088, 1140861190, 1140861276, 1140866072, 1140866078, 1140866090, 1140866102, 1140866108, 1140866122, 1140866138, 1140866156, 1140866162, 1140866724, 1140866738, 1140868618, 1140872568, 1140874706, 1140874744, 1140875808, 1140879758, 1140879760, 1140879762, 1140879802, 1140879806, 1140879810, 1140879818, 1140879822, 1140879826, 1140879830, 1140879834, 1140879842, 1140879866, 1140884298, 1140888552, 1140888556, 1140888560, 1140888646, 1140909706, 1140910442, 1140910614, 1140916356, 1140923272, 1140923336, 1140923404, 1140923712, 1140926778, 1140928226, 1141145660, 1141146126, 1141152998, 1141153026, 1141164276, 1141165470, 1141166006, 1141169516, 1141171336, 1141180592, 1141180772, 1141180778, 1141184722, 1141193282, 1141194794, 1141194810 | |

##### 2.3.3 Table of recategorization of UK Biobank variables to match the categories in QStroke variables

| QStroke | | UK Biobank | | |
|---|---|---|---|---|
| Variable name | Categories | Variable name | Categories | Notes |
| ethnicity (self-reported) | White or not stated, Indian, Pakistani, Bangdaleshi, Other Asian, Black Caribbean, Black African, Chinese, Other ethnic group | 21000: Ethnic background | **White or not stated**<br>White (British, Irish, Any other white background), Do not know, Prefer not to answer<br>**Indian**<br>Indian<br>**Pakistani**<br>Pakistani<br>**Bangdaleshi**<br>Bangladeshi<br>**Other Asian**<br>Any other Asian background<br>**Black Caribbean**<br>Caribbean<br>**Black African**<br>African<br>**Chinese**<br>Chinese<br>**Other ethnic group**<br>Mixed (White and Black Caribbean, White and Black African, White and Asian, Any other mixed background), Any other Black background, Other ethnic group | White (British, Irish, Any other white background), Mixed (White and Black Caribbean, White and Black African, White and Asian, Any other mixed background), Asian or Asian British (Indian, Pakistani, Bangladeshi, Any other Asian background), Black or Black British (Caribbean, African, Any other Black background), Chinese, Other ethnic group, Do not know, Prefer not to answer |
| smoking status | non-smoker, ex-smoker, light smoker (less than 10), moderate smoker (10 to 19), heavy smoker (over 20) | 20116: Smoking status<br>1239: Current tobacco smoking<br>3456: Number of cigarettes currently smoked daily (current cigarette smokers) | **non-smoker:**<br>20116: Smoking status: Never<br>1239: Current tobacco smoking: No<br>**ex-smoker:**<br>20116: Smoking status: Previous<br>**light smoker (less than 10), moderate smoker (10 to 19), heavy smoker (over 20):**<br>1239: Only occasionally<br>3456: Number of cigarettes currently smoked daily (current cigarette smokers) | If current smoker based on 1239, the number of cigarettes smoked taken from 3456. If 3456 is unavailable, then separate category "smoker". |

### 3 Outcomes and exclusion criteria

| Model | Outcome (ICD10 codes in original paper) | Exclusion criteria | Outcome variable name in UK Biobank | Exclusion criteria variable name in UK Biobank | Notes |
|---|---|---|---|---|---|
| QDiabetes | 10-year risk of type 2 diabetes (E11) | Type 1 or type 2 diabetes at study entry, missing Townsend score, fasting blood glucose concentration of 7 mmol/L or more or HBA1c value of 48 mmol/mol | 41270: Diagnoses - ICD10 (E11, E10)<br>41280: Date of first in-patient diagnosis - ICD10<br><br>41271: Diagnoses - ICD9 (25000, 25010, 25020, 25030, 25040, 25050, 25060, 25070, 25080, 25090, 25002, 25012, 25022, 25032, 25042, 25052, 25062, 25072, 25082, 25092)<br>41281: Date of first in-patient diagnosis - ICD9 | In addition to variables to define the outcome:<br>41270: Diagnoses - ICD10 (E10)<br><br>41271: Diagnoses - ICD9 (25001, 25011, 25021, 25031, 25041, 25051, 25061, 25071, 25081, 25091, 25003, 25013, 25023, 25033, 25043, 25053, 25063, 25073, 25083, 25093)<br><br>20002: Non-Cancer Illness Code (1222, 1223,1220)<br><br>2443: Diabetes diagnosed by doctor (instance 0)<br><br>120007: Ever had diabetes (Type I or Type II)<br><br>30740: Glucose<br>Fasting time from 74: Fasting time<br><br>20003: Treatment/medication code (1140868902, 1140874646, 1140874674, 1140874718, 1140874744, 1140883066, 1140884600, 1141152590, 1141157284, 1141168660, 1141171646, 1141173882, 1141189090)<br><br>30750: Glycated haemoglobin (HbA1c) | Date only available for O24 (diabetes mellitus in pregnancy) but added filter so participants have O24.1<br><br>Medications taken from (for those that are available):<br>https://www.diabetes.co.uk/Diabetes-drugs.html |
| QRISK3 | 10-year risk of cardiovascular diseases (G45, I20, I21, I22, I23, I24, I25, I63, I64) | Cardiovascular disease, use of statins at study entry, missing Townsend score | 41270: Diagnoses - ICD10 (G45, I20, I21, I22, I23, I24, I25, I63, I64)<br>41280: Date of first in-patient diagnosis - ICD10<br><br>41271: Diagnoses - ICD9 (410, 411, 412, 413, 414, 434, 436)<br>41281: Date of first in-patient diagnosis - ICD9<br><br>41272: Operative procedures - OPCS4 (K40, K41, K42, K43, K44, K45, K46, K47.1, K49, K50, K75)<br>41282: Date of first operative procedure - OPCS4 | In addition to variables to define the outcome:<br><br>20003: Treatment/medication code (atorvastatin, simvastatin, fluvastatin, rosuvastatin, pravastatin)<br><br>20002: Non-Cancer Illness Code (1074, 1075, 1082, 1583)<br><br>20004: Operation code (1070, 1071, 1095, 1105, 1109, 1514)<br><br>6150: Vascular/heart problems diagnosed by a doctor (1, 2, 3)<br><br>22189: Townsend deprivation index at recruitment | Statins listed in a paper QDiabetes refered to: https://www.sciencedirect.com/science/article/pii/S0140673609619656?via%3Dihub<br>Numbers don't match the external validation |
| QStroke | 10-year risk of ischaemic stroke or transient ischaemic attack (I63, I64) | Prior recorded diagnosis of stroke or transient ischaemic attack, use of anticoagulants at study entry, missing Townsend score | 41270: Diagnoses - ICD10 (I63, I64)<br>41280: Date of first in-patient diagnosis - ICD10<br><br>41271: Diagnoses - ICD9 (433, 434)<br>41281: Date of first in-patient diagnosis - ICD9 | 20003: Treatment/medication code (warfarin, acenocoumarol, phenindione, dabigatran, rivaroxaban, apixaban)<br><br>20002: Non-Cancer Illness Code (1583, 1082)<br><br>6150: Vascular/heart problems diagnosed by doctor (stroke)<br><br>22189: Townsend deprivation index at recruitment | Anticoagulants from 2.8.2 of the British National Formulary |

### 4 Baseline characteristics

#### 4.1 Table of baseline characteristics for variables included in QRISK3

Table comparing baseline characteristics of the QRISK3 derivation cohort and the UK Biobank validation cohort for QRISK3.

| | UK Biobank, female n = 234 808 | QRISK3 derivation cohort, female n = 4 019 956 | UK Biobank, male n = 171 047 | QRISK3 derivation cohort, male n = 3 869 847 |
|---|---|---|---|---|
| Age (years), mean (SD) | 55.6 (8.0) | 43.3 (15.3) | 55.4 (8.2) | 42.6 (14.0) |
| Systolic blood pressure (mm Hg), mean (SD) | 136.4 (20.2) | 123.2 (18.2) | 142.2 (18.4) | 129.2 (16.3) |
| Measure of systolic blood pressure variability (SD of repeated measures), mean (SD) | 5.4 (4.5) | 9.3 (6.2) | 5.2 (4.3 | 9.9 (6.8) |
| Total cholesterol: high-density lipoprotein cholesterol ratio (mmol/L), mean (SD) | 3.9 (1.0) | 3.7 (1.2) | 4.6 (1.1) | 4.4 (1.4) |
| Family history of coronary heart disease in first degree relative <60 years | 100846 (42.9) | 481 628 (12.0) | 61886 (36.2) | 357 987 (9.3) |
| Self-reported ethnicity | | | | |
| White or not stated | 221511 (94.3) | 3 564 651 (88.7) | 160779 (94.0) | 3 435 408 (88.8) |
| Indian | 2330 (1.0) | 77 683 (1.9) | 1977 (1.2) | 81 805 (2.1) |
| Pakistani | 555 (0.2) | 39 541 (1.0) | 738 (0.4) | 46 948 (1.2) |
| Bangladeshi | 50 (0.02) | 31 930 (0.8) | 106 (0.1) | 42 111 (1.1) |
| Other Asian | 690 (0.3) | 53 559 (1.3) | 673 (0.4) | 45 753 (1.2) |
| Black Caribbean | 2383 (1.0) | 37 781 (0.9) | 1341 (0.8) | 30 610 (0.8) |
| Black African | 1421 (0.6) | 77 813 (1.9) | 1388 (0.8) | 71 245 (1.8) |
| Chinese | 888 (0.4) | 33 767 (0.8) | 497 (0.3) | 23 730 (0.6) |
| Other ethnic group | 4588 (2.0) | 103 231 (2.6) | 3201 (1.9) | 92 237 (2.4) |
| Townsend deprivation score, mean (SD) | -1.4 (3.0) | 0.4 (3.2) | -1.3 (3.1) | 0.5 (3.3) |
| Body mass index (kg/m2), mean (SD) | 26.7 (5.0) | 25.4 (5.1) | 27.4 (4.1) | 25.9 (4.2) |
| Smoking status | | | | |
| Non-smoker | 141372 (60.2) | 2 051 803 (51.0) | 88892 (52.0) | 1 463 941 (37.8) |
| Ex-smoker | 71834 (30.6) | 589 521 (14.7) | 59840 (35.0) | 594 265 (15.4) |
| Light smoker (<10 a day) | 8791 (3.7) | 434 954 (10.8) | 8415 (4.9) | 507 523 (13.1) |
| Moderate smoker (10–19 a day) | 6939 (3.0) | 226 128 (5.6) | 5207 (3.0) | 251 170 (6.5) |
| Heavy smoker (20 or over a day) | 4428 (1.9) | 115 890 (2.9) | 5693 (3.3) | 188 857 (4.9) |
| Atrial fibrillation | 1016 (0.4) | 15 177 (0.4) | 1822 (1.1) | 20 098 (0.5) |
| Erectile dysfunction | NA | NA | 351 (0.2) | 90 753 (2.3) |
| Migraine | 11398 (4.9) | 257 825 (6.4) | 3099 (1.8) | 103 995 (2.7) |
| Rheumatoid arthritis | 3313 (1.4) | 45 700 (1.1) | 1225 (0.7) | 20 997 (0.5) |
| Chronic kidney disease (stage 3-5) | 189 (0.1) | 19 396 (0.5) | 129 (0.1) | 12 254 (0.3) |
| Severe mental illness | 846 (0.4) | 274 069 (6.8) | 731 (0.4) | 167 115 (4.3) |
| Systemic lupus erythematosus | 457 (0.2) | 4010 (0.1) | 47 (0.03) | 365 (0.0) |
| Diabetes type 1 | 2618 (1.1) | 10 060 (0.3) | 3541 (2.1) | 11 617 (0.3) |
| Diabetes type 2 | 2922 (1.2) | 48 022 (1.2) | 3746 (2.2) | 58 395 (1.5) |
| Atypical antipsychotic | 487 (0.2) | 19 140 (0.5) | 467 (0.3) | 20 123 (0.5) |
| Corticosteroid | 1579 (0.7) | 96 955 (2.4) | 1024 (0.6) | 56 533 (1.5) |
| Treated hypertension | 30543 (13.0) | 223 494 (5.6) | 23152 (13.5) | 164 255 (4.2) |

#### 4.2 Table of baseline characteristics for variables included in QDiabetes

Table comparing baseline characteristics of the QDiabetes derivation cohort and the UK Biobank validation cohort for QDiabetes. The number of participants diagnosed with gestational diabetes, learning disability, and PCOS in the validation cohort is significantly lower compared to the participants in the derivation cohort, as the UK Biobank are older in age and the medical history is not available that far back.

| | UK Biobank<br>n = 458 700 | QDiabetes derivation cohort<br>n = 8 186 705 |
|---|---|---|
| Age (years), mean (SD) | 56.3 (8.1) | 44.9 (15.3) |
| Townsend deprivation score, mean (SD) | -1.4 (3.1) | 0.5 (3.3) |
| Body mass index (kg/m2), mean (SD) | 27.1 (4.6) | 26.0 (5.0) |
| Fasting blood glucose, mean (SD) | 4.9 (0.6) | 5.0 (0.6) |
| HbA1c, mean (SD) | 34.9 (3.7) | 37.2 (4.5) |
| Self-reported ethnicity | | |
| White or not stated | 434082 (94.6) | 7 136 377 (87.2) |
| Indian | 4617 (1.0) | 188 049 (2.3) |
| Pakistani | 1304 (0.3) | 101 231 (1.2) |
| Bangladeshi | 152 (0.03) | 81 834 (1.0) |
| Other Asian | 1459 (0.3) | 122 981 (1.5) |
| Black Caribbean | 3759 (0.8) | 80 657 (1.0) |
| Black African | 2855 (0.6) | 179 423 (2.2) |
| Chinese | 1406 (0.3) | 65 999 (0.8) |
| Other ethnic group | 8290 (1.8) | 230 154 (2.8) |
| Smoking status | | |
| Non-smoker | 252878 (55.1) | 4 441 795 (54.3) |
| Ex-smoker | 155195 (33.8) | 1 518 799 (18.6) |
| Light smoker (<10 a day) | 19332 (4.2) | 1 098 645 (13.4) |
| Moderate smoker (10–19 a day) | 14045 (3.1) | 485 756 (5.9) |
| Heavy smoker (20 or over a day) | 12026 (2.6) | 289 649 (3.5) |
| Family history of diabetes in a first degree relative | 91225 (19.9) | 1 21 8682 (14.9) |
| Treated hypertension | 89229 (19.5) | 737 303 (9.0) |
| Gestational diabetes | 19 (0.004) | 17 214 (0.4) |
| Learning disability | 11 (0.002) | 56 092 (0.7) |
| Bipolar disorder or schizophrenia | 1386 (0.3) | 62 014 (0.8) |
| Polycystic ovary syndrome | 38 (0.008) | 81 164 (2.0) |
| Previous heart attack, angina, stroke or transient ischemic attack | 9792 (2.1) | 290 345 (3.5) |
| Atypical antipsychotic | 1096 (0.2) | 58 655 (0.7) |
| Corticosteroid | 3183 (0.7) | 238 683 (2.9) |
| Statins | 55722 (12.1) | 526 969 (6.4) |

#### 4.3 Table of baseline characteristics for variables included in QStroke

Table comparing baseline characteristics of the QStroke derivation cohort and the UK Biobank validation cohort for QStroke.

| | UK Biobank n = 490 471 | QStroke derivation cohort n = 3 549 478 |
|---|---|---|
| Age (years), mean (SD) | 56.5 (8.1) | 45.0 (15.4) |
| Townsend deprivation score, mean (SD) | -1.3 (3.1) | −0.1 (3.5) |
| Self-reported ethnicity | | |
| White or not stated | 461189 (94.0) | 3 333 695 (93.9) |
| Indian | 5857 (1.2) | 43 175 (1.2) |
| Pakistani | 1805 (0.4) | 21 101 (0.6) |
| Bangladeshi | 232 (0.05) | 13 878 (0.4) |
| Other Asian | 1828 (0.4) | 26 435 (0.7) |
| Black Caribbean | 4424 (0.9) | 17 488 (0.5) |
| Black African | 3331 (0.7) | 36 952 (1.0) |
| Chinese | 1551 (0.3) | 13 862 (0.4) |
| Other ethnic group | 9372 (1.9) | 42 892 (1.2) |
| Family history of coronary heart disease in first degree relative <60 years | 209322 (42.7) | 424 745 (12.0) |
| Atrial fibrillation | 3561 (0.7) | 15 371 (0.4) |
| Rheumatoid arthritis | 5990 (1.2) | 20 580 (0.6) |
| Chronic kidney disease (stage 4-5) | 587 (0.1) | 6 536 (0.2) |
| Diabetes type 1 | 23035 (4.7) | 11 636 (0.3) |
| Diabetes type 2 | 24156 (4.9) | 79 567 (2.2) |
| Treated hypertension | 108683 (22.2) | 236 900 (6.7) |
| Coronary heart disease | 20627 (4.2) | 99 561 (2.8) |
| Congestive cardiac failure | 480 (0.1) | 16 294 (0.5) |
| Valvular heart disease | 4213 (0.9) | 13 510 (0.4) |
| Body mass index (kg/m2), mean (SD) | 27.4 (4.8) | 26.0 (4.6) |
| Systolic blood pressure (mm Hg), mean (SD) | 139.8 (19.7) | 129.9 (19.9) |
| Total cholesterol: high-density lipoprotein cholesterol ratio (mmol/L), mean (SD) | 4.1 (1.1) | 4.1 (1.3) |
| Smoking status | | |
| Non-smoker | 268 023 (54.6) | 1 789 104 (50.4) |
| Ex-smoker | 168 027 (34.3) | 595 287 (16.8) |
| Light smoker (<10 a day) | 20 473 (4.2) | 246 174 (6.9) |
| Moderate smoker (10–19 a day) | 15 016 (3.1) | 293 150 (8.3) |
| Heavy smoker (20 or over a day) | 13 101 (2.7) | 203 303 (5.7) |

### 5 Follow up time and number of outcome

#### 5.1 Table of summary follow up time for QRISK3, QDiabetes, and QStroke.

The follow up times are calculated in years for each model after applying exclusion criteria.

| Model | Min. | 1st Qu. | Median | Mean | 3rd Qu. | Max. |
|---|---|---|---|---|---|---|
| QRISK3 | 0.003 | 12.734 | 13.596 | 12.946 | 14.283 | 16.663 |
| QDiabetes | 0.003 | 12.802 | 13.624 | 13.158 | 14.294 | 16.663 |
| QStroke | 0.008 | 12.871 | 13.632 | 13.207 | 14.297 | 16.663 |

#### 5.2 Table of number of outcomes for QRISK3, QDiabetes, and Stroke

The number of outcomes are calculated for each model in 10 years. The outcomes are defined as the first occurrence of the disease of interest derived from hospital inpatient and death records.

| | QRISK3 | QDiabetes | QStroke |
|---|---|---|---|
| Number of outcomes | 20256 | 8641 | 5065 |

### 6 Missingness

The missingness was assessed for all variables included in the models and the variables defining the subgroups excluding diagnostic and medication codes as these were obtained from registry linkages and assumed to represent the true values. The percentage of missingness is calculated based on the number of participants included in each model.

| Variable | QRISK3 (% of total) | QDiabetes (% of total) | QStroke (% of total) |
|---|---|---|---|
| average total household income before tax | 4604 (1.13) | 5321 (1.16) | 5816 (1.19) |
| bmi | 2335 (0.57) | 2622 (0.57) | 2931 (0.60) |
| ethnicity | 739 (0.18) | 776 (0.17) | 882 (0.18) |
| index of multiple deprivation | 10220 (2.52) | 11551 (2.52) | 12349 (2.52) |
| qualifications | 3823 (0.94) | 4242 (0.92) | 4534 (0.92) |
| sex | 0 (0) | 0 (0) | 0 (0) |
| smoking status | 4444 (1.09) | 5224 (1.14) | 5831 (1.19) |
| systolic blood pressure | 1116 (0.27) | 1152 (0.25) | 1298 (0.26) |
| systolic blood pressure variability | 1471 (0.36) | 1537 (0.34) | Not in the model |
| total cholesterol | 58960 (14.52) | Not in the model | 70882 (14.45) |
| year immigrated to UK * | 762 (0.19) | 883 (0.17) | 981 (0.16) |
| HbA1c | Not in the model | 32811 (7.15) | Not in the model |
| Fasting blood glucose | Not in the model | 67078 (14.62) | Not in the model |

*Calculated for participants for who the country of birth (UK/elsewhere) takes values "elsewhere" or "Republic of Ireland".

### 7 Differences of predicted and observed values

The differences of the predicted and observed values compares the calibration of the models for different demographic subgroups. The differences are calculated based on deciles of predicted risk for each model and the results are pooled from five MICE imputed datasets using Rubin's rule.

E.g. Figure 2.1 shows the difference of predicted and observed values based on sex. This is calculated by dividing the validation cohort of QRISK3 into deciles based on the predicted QRISK3 score. The observed values for each of the deciles are obtained using IPCW weighted Kaplan-Meier estimates. The deciles are then examined based on sex and the observed values are subtracted from the predicted values. The differences are calculated for each MICE imputed datasets and pooled using Rubin's rule.

#### 7.1 Difference of predicted and observed values based on sex

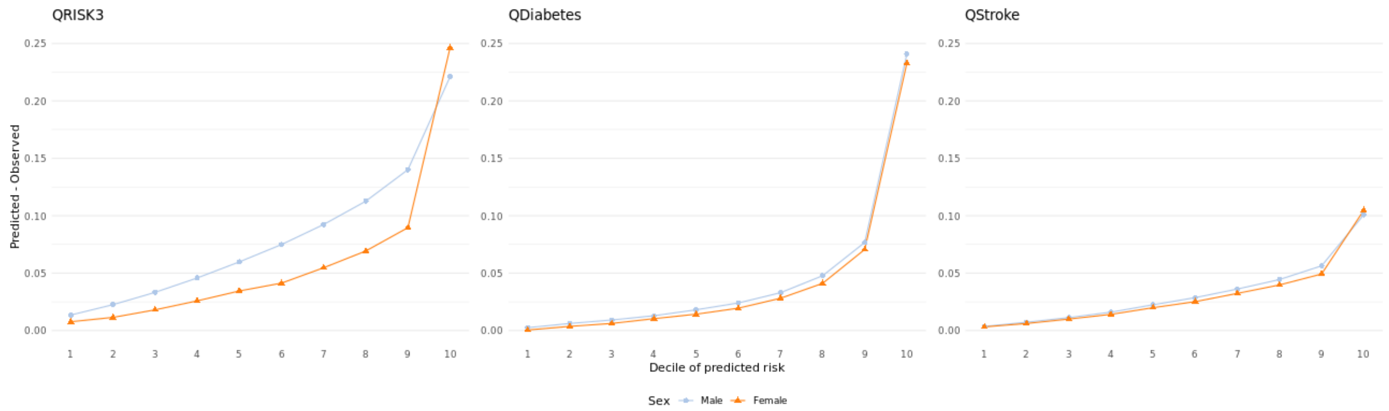

#### 7.2 Difference of predicted and observed values based on ethnicity

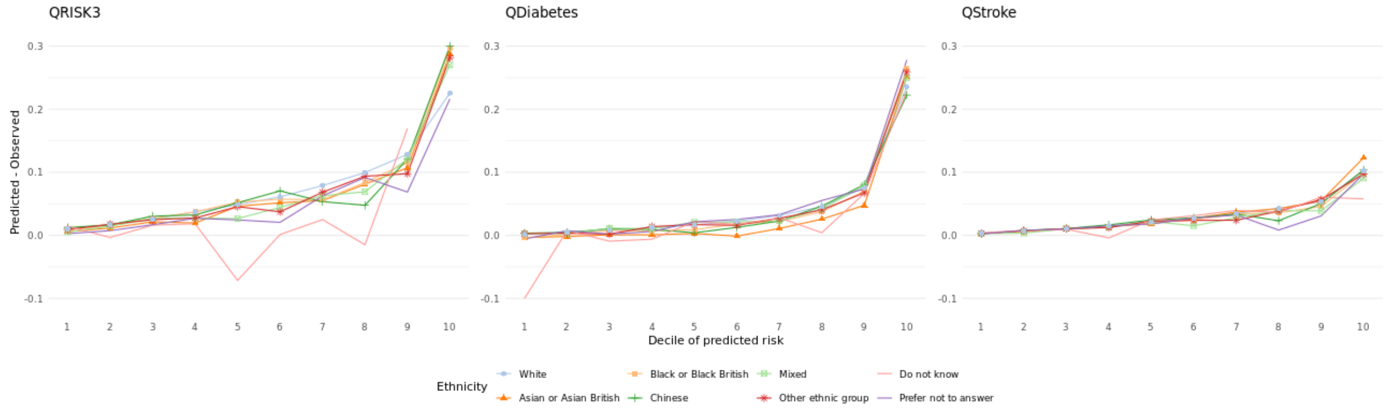

#### 7.3 Difference of predicted and observed values based on age at baseline assessment

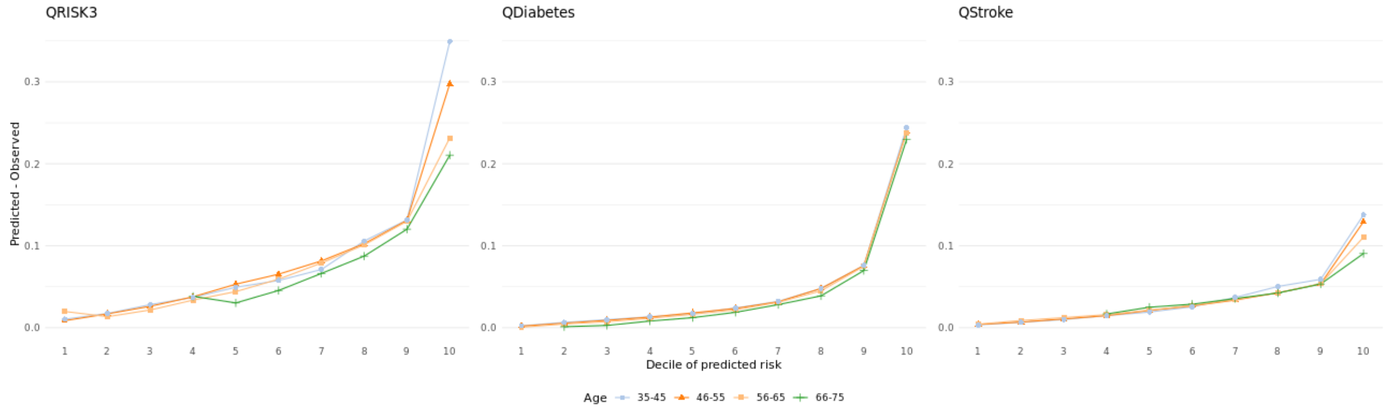

#### 7.4 Difference of predicted and observed values based on immigration status

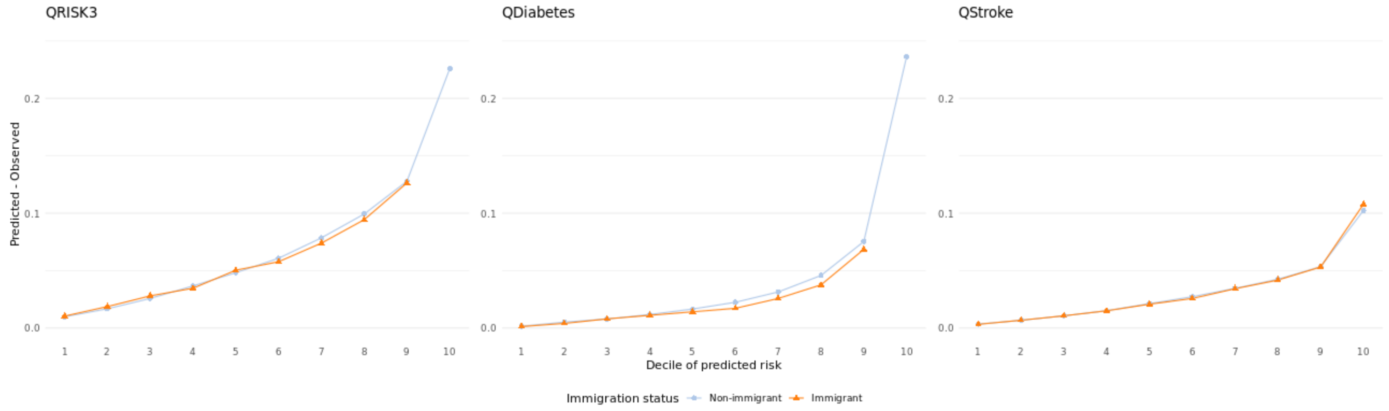

#### 7.5 Difference of predicted and observed values based on average household income before taxation

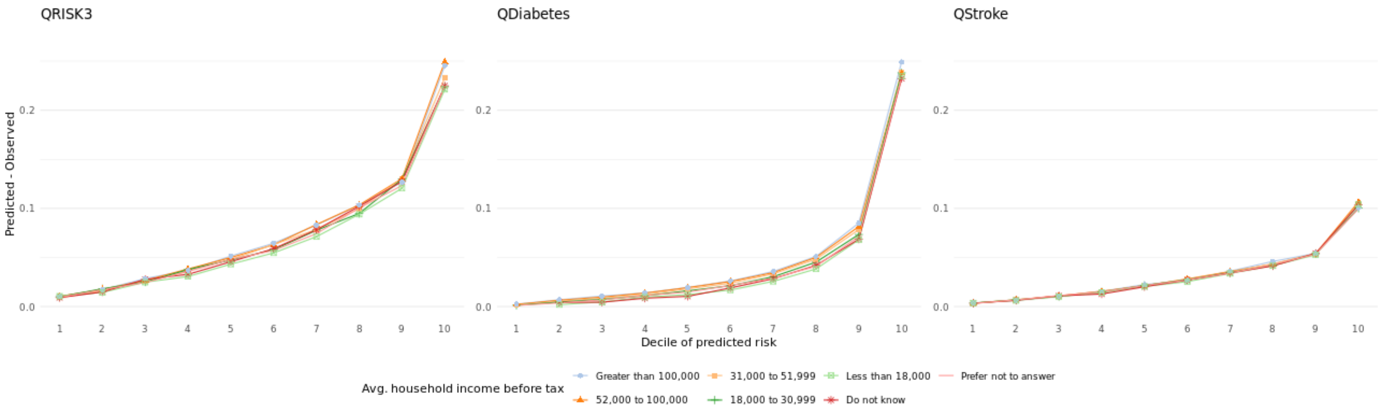

#### 7.6 Difference of predicted and observed values based on education level

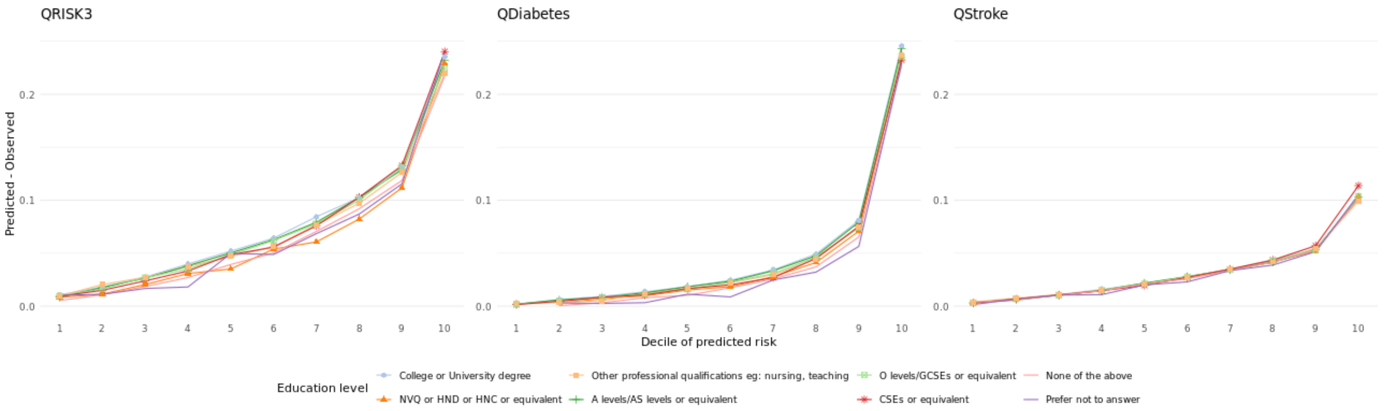

#### 7.7 Difference of predicted and observed values based on Townsend deprivation score

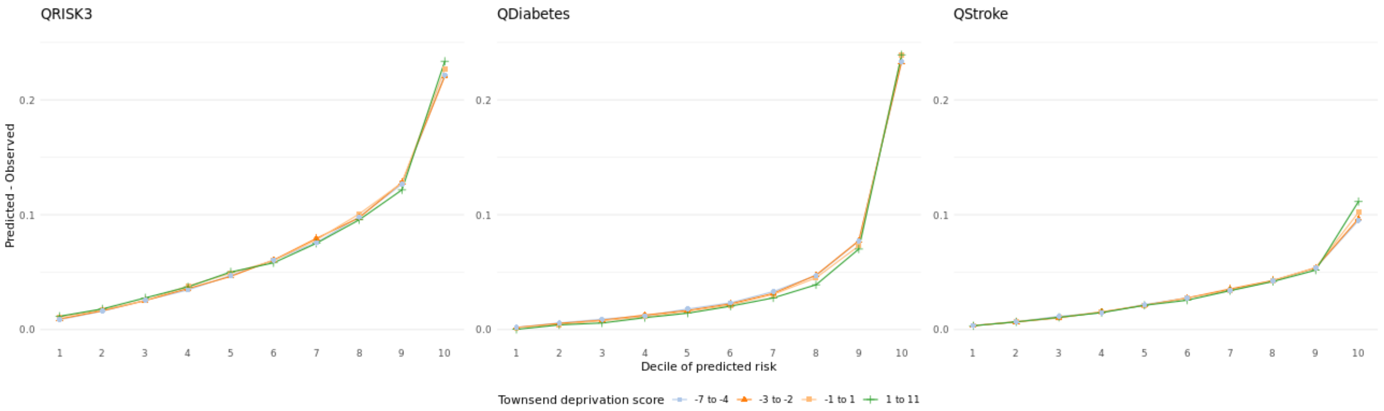

#### 7.8 Difference of predicted and observed values based on index of multiple deprivation

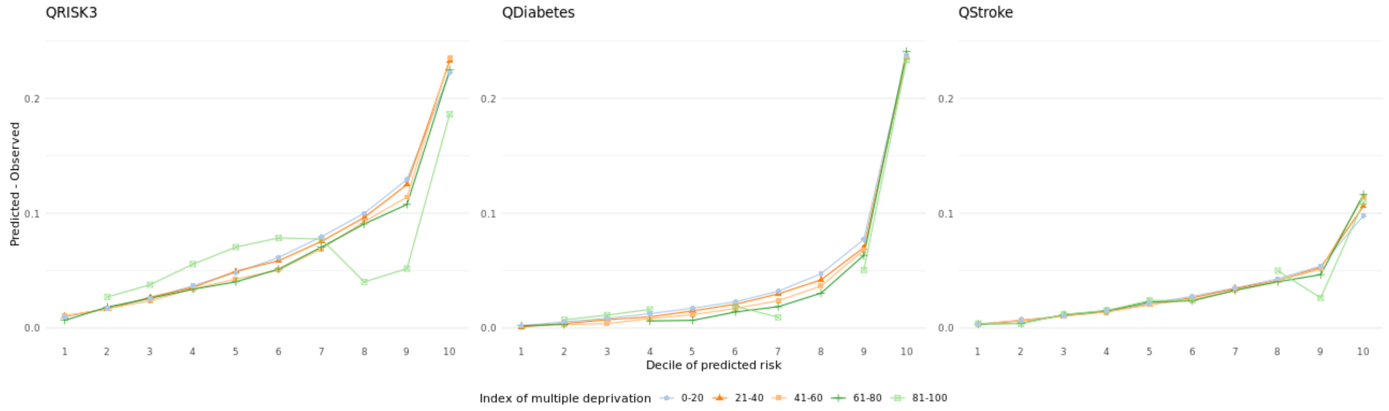

### 8 Calibration plots

Calibration plots show the calibration by comparing the predicted values to the observed values for the models overall as well as for demographic subgroups. The values are calculated based on deciles of predicted risk for each model and all categories within each subgroup and the results are pooled from five imputed datasets using Rubin's rule.

E.g. The first plot of figure 3.2 shows the calibration of QRISK3 based on sex. This is calculated by dividing the validation cohort of QRISK3 into deciles based on the predicted QRISK3 score for each sex separately. The observed values for each of the deciles and sexes are obtained using IPCW weighted Kaplan-Meier estimates. The values are calculated for each MICE imputed datasets and pooled using Rubin's rule.

#### 8.1 Calibration of QRISK3, QDiabetes, and Stroke overall

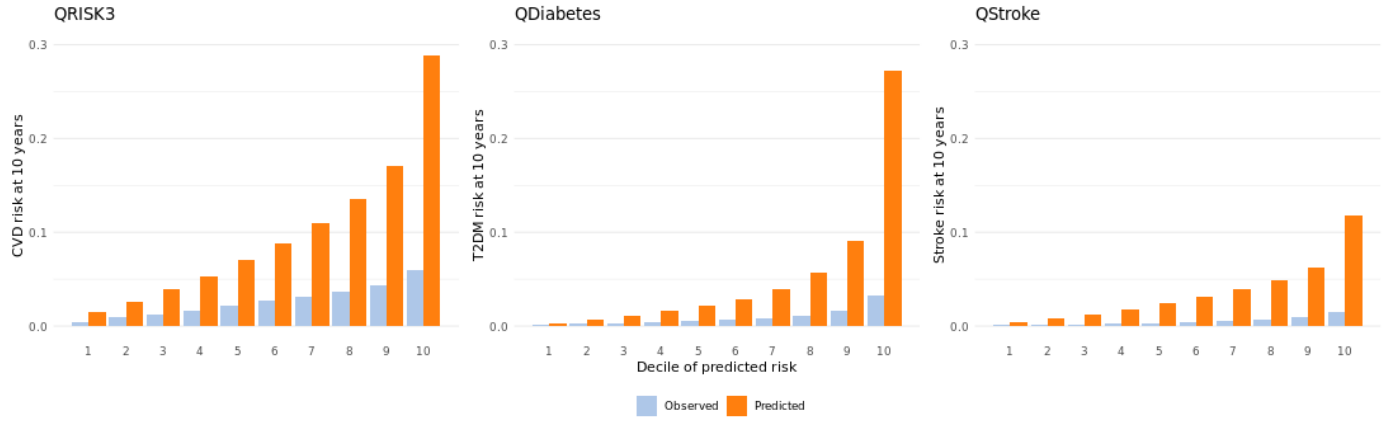

#### 8.2 Calibration plot based on sex

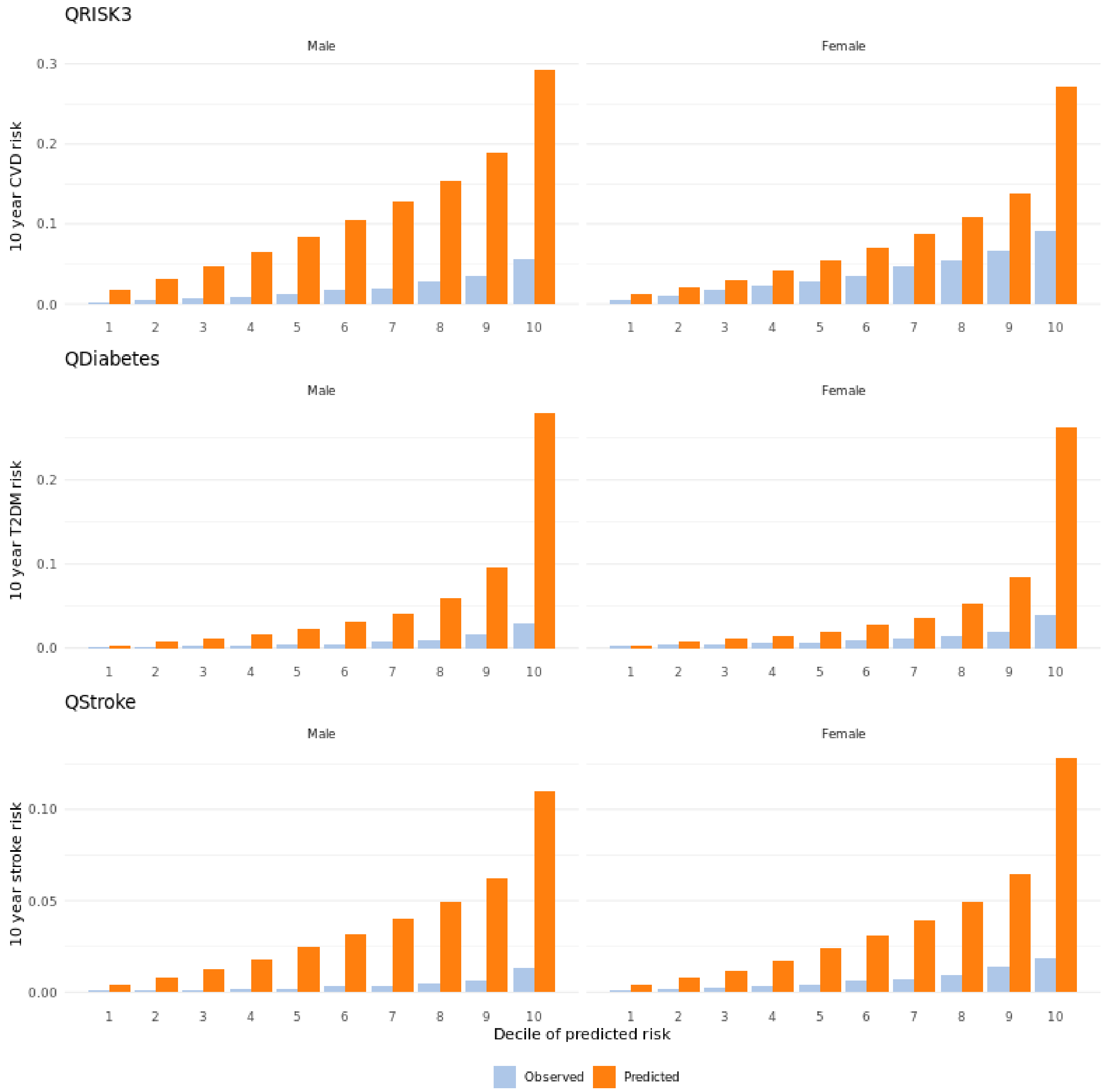

#### 8.3 Calibration plot based on ethnicity

QRISK3

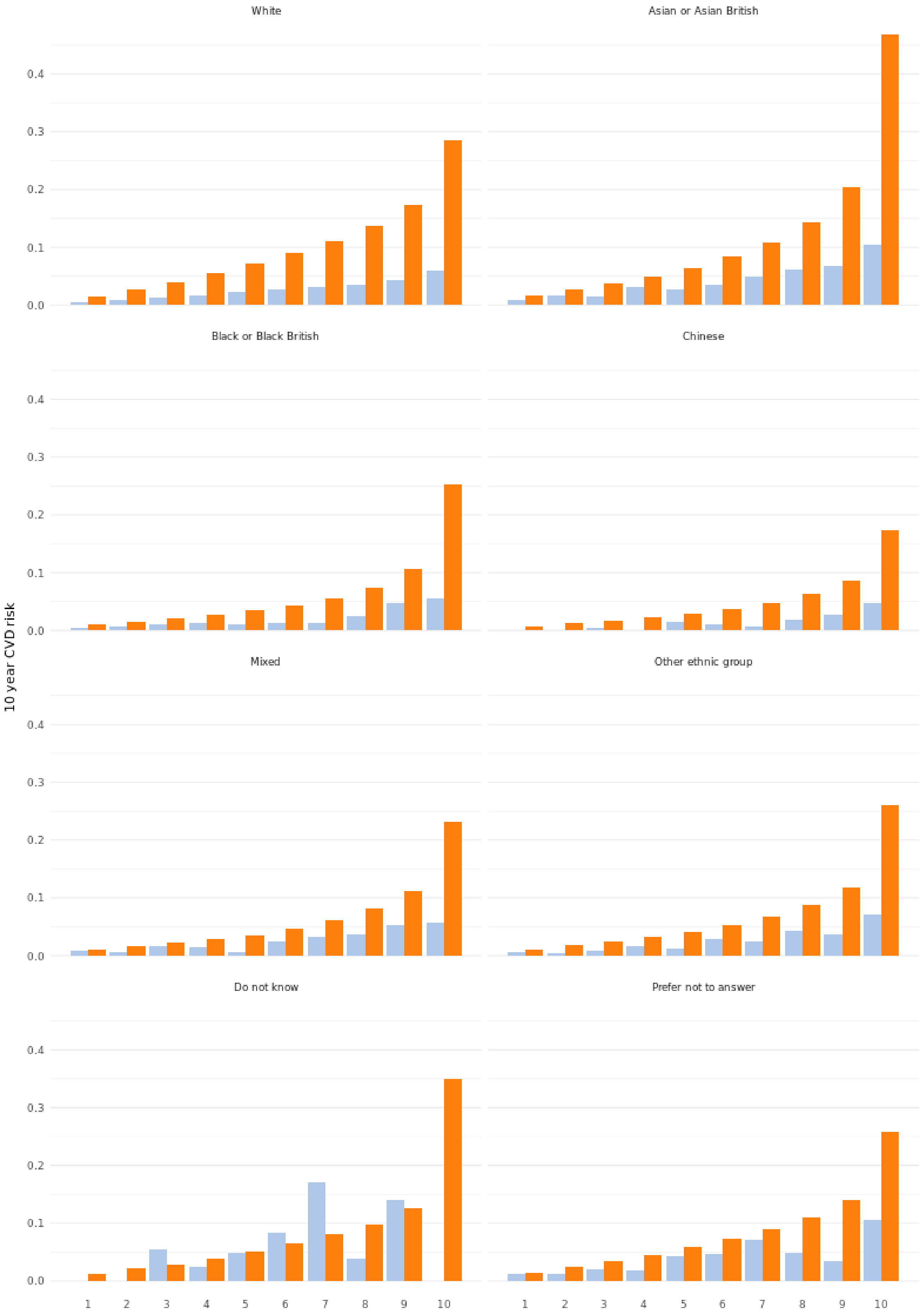

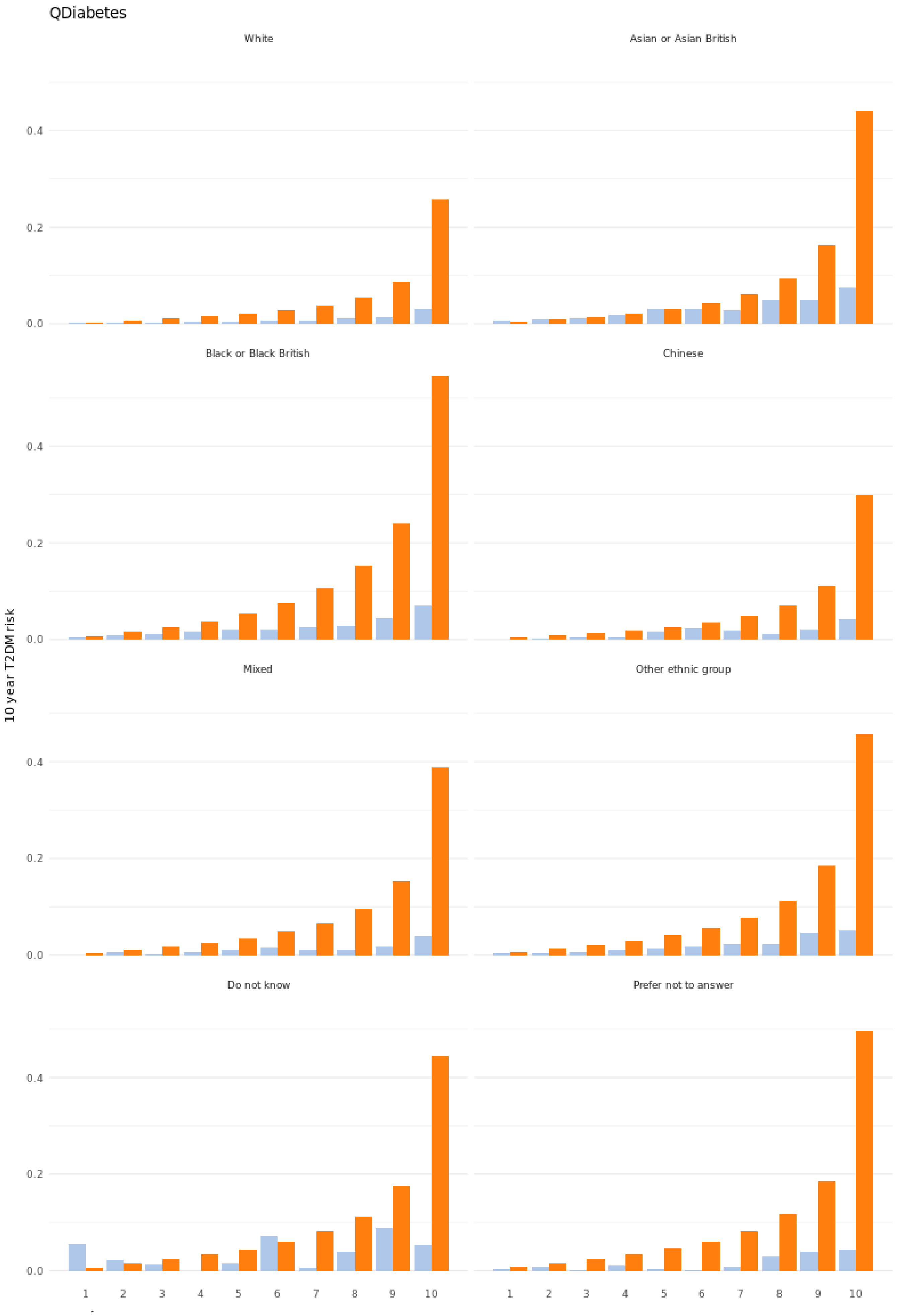
QDiabetes
White
Asian or Asian British
Black or Black British
Chinese
Mixed
Other ethnic group
Do not know
Prefer not to answer
10 year T2DM risk
0.0
0.2
0.4
1
2
3
4
5
6
7
8
9
10

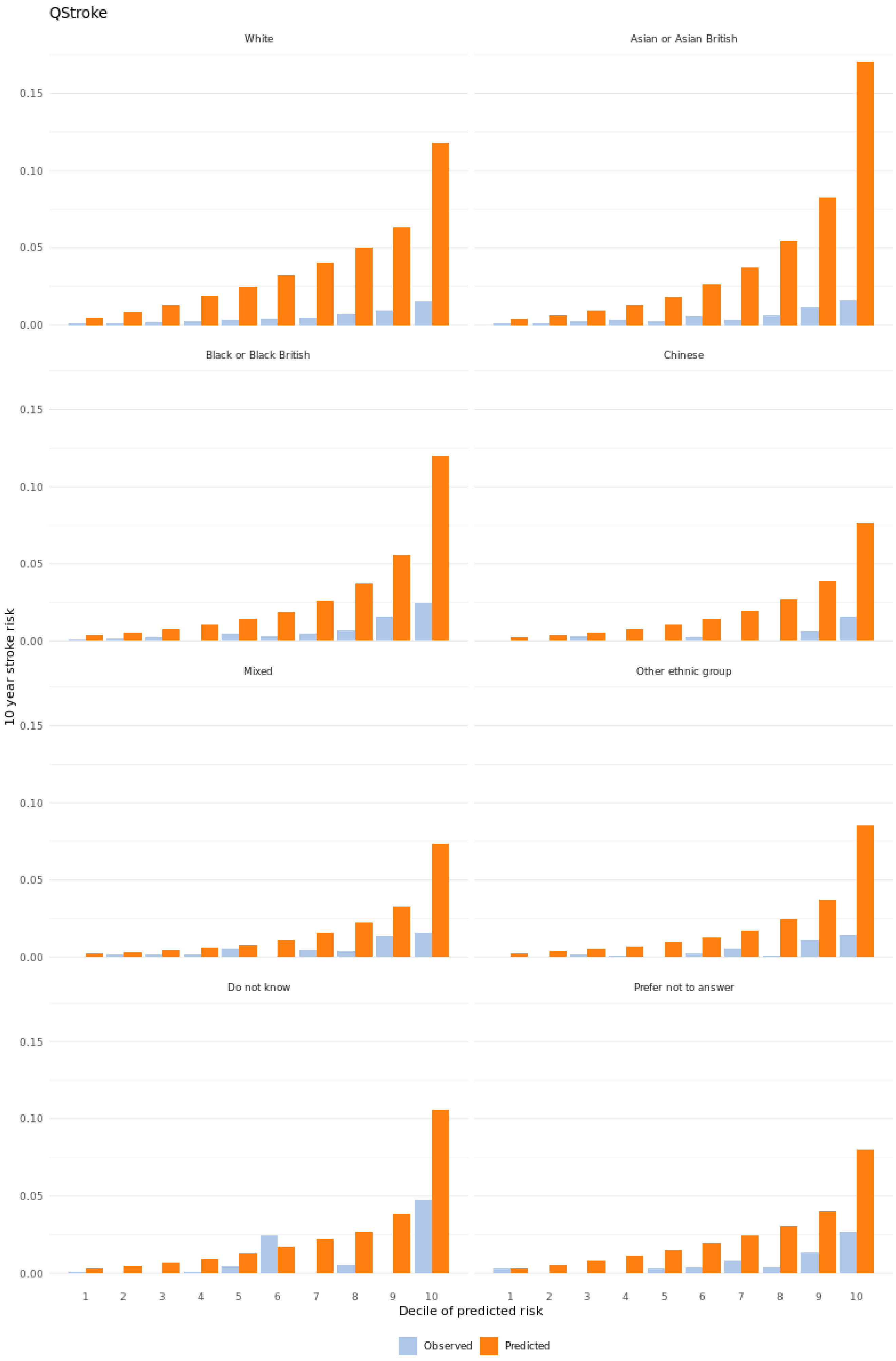

QStroke
White
Asian or Asian British
Black or Black British
Chinese
Mixed
Other ethnic group
Do not know
Prefer not to answer
0.15
0.10
0.05
0.00
10 year stroke risk
1 2 3 4 5 6 7 8 9 10
Decile of predicted risk
Observed
Predicted

### 8.4 Calibration plot based on age at baseline assessment

QRISK3

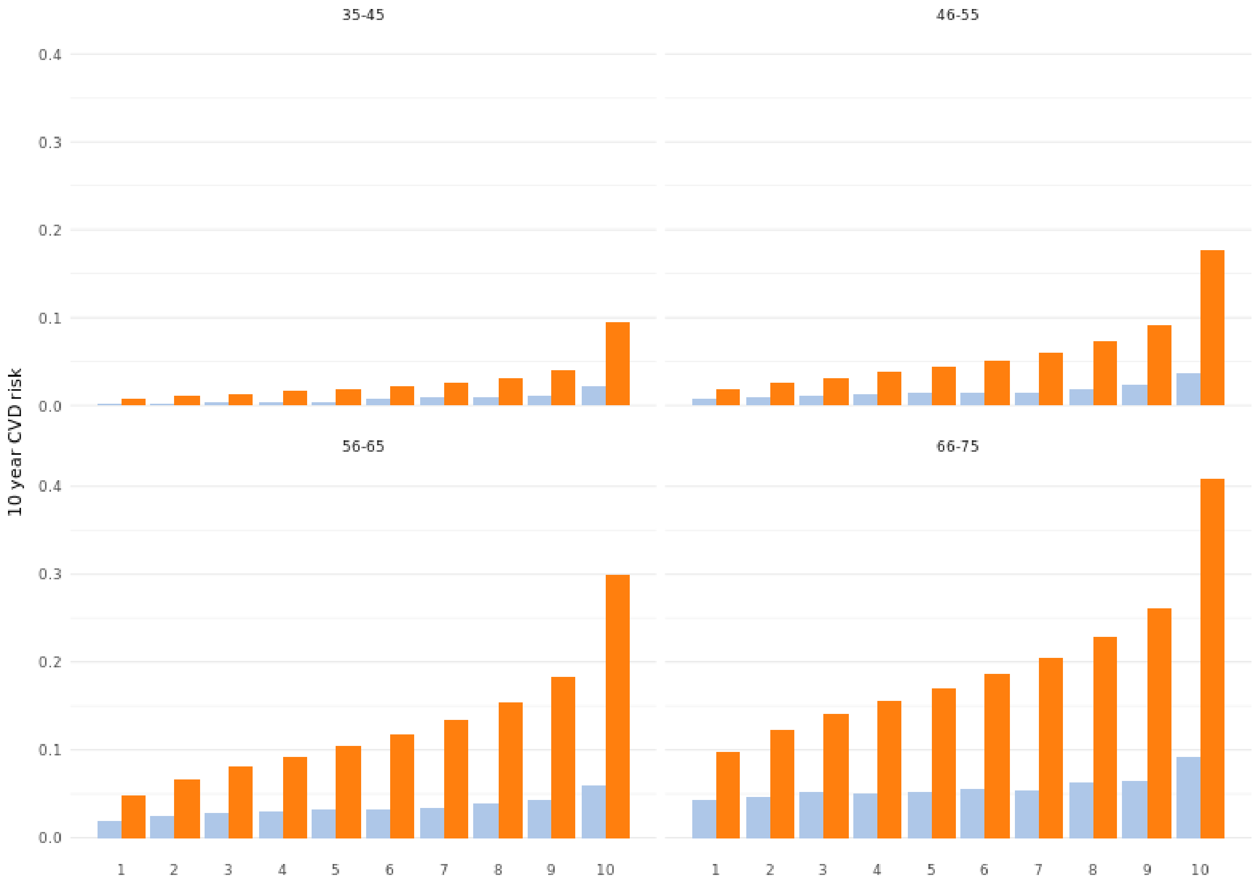

QDiabetes

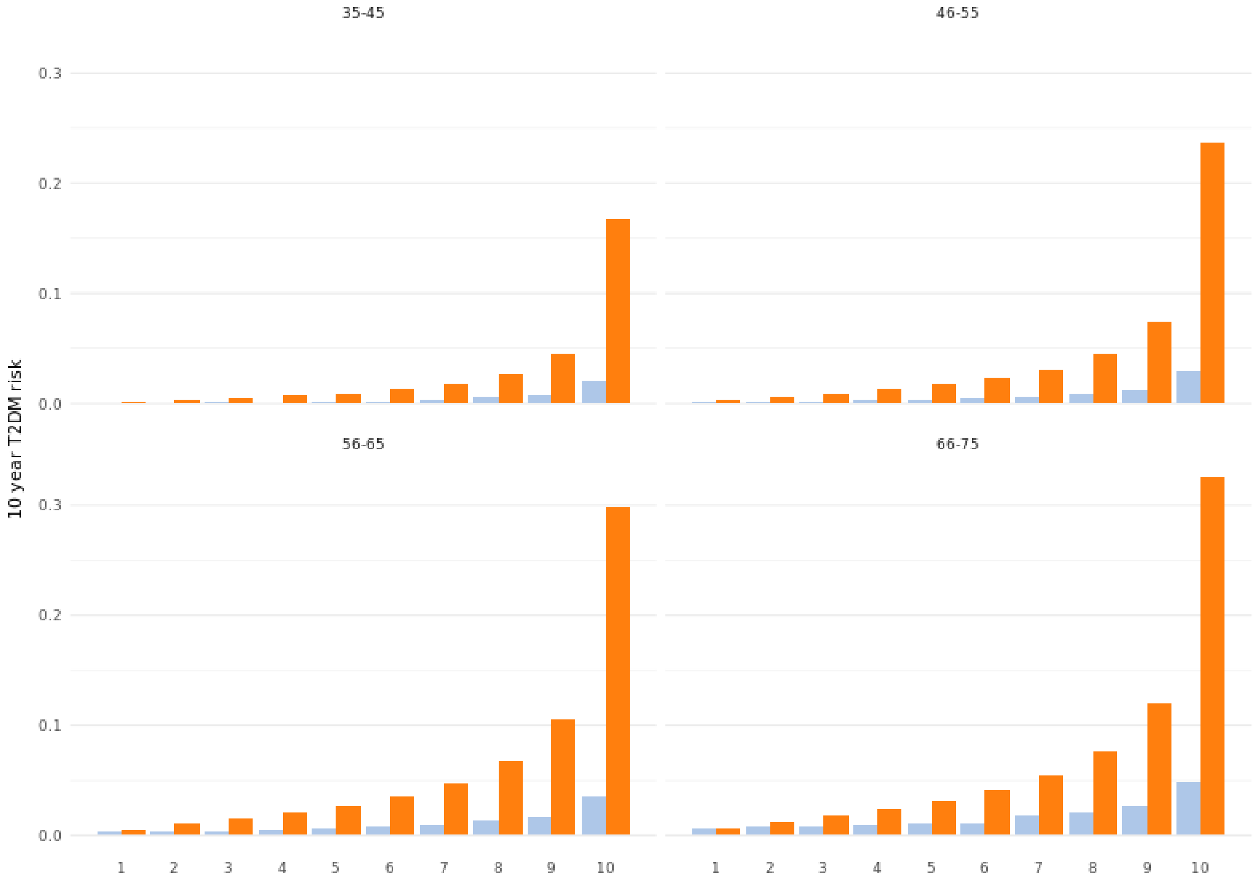

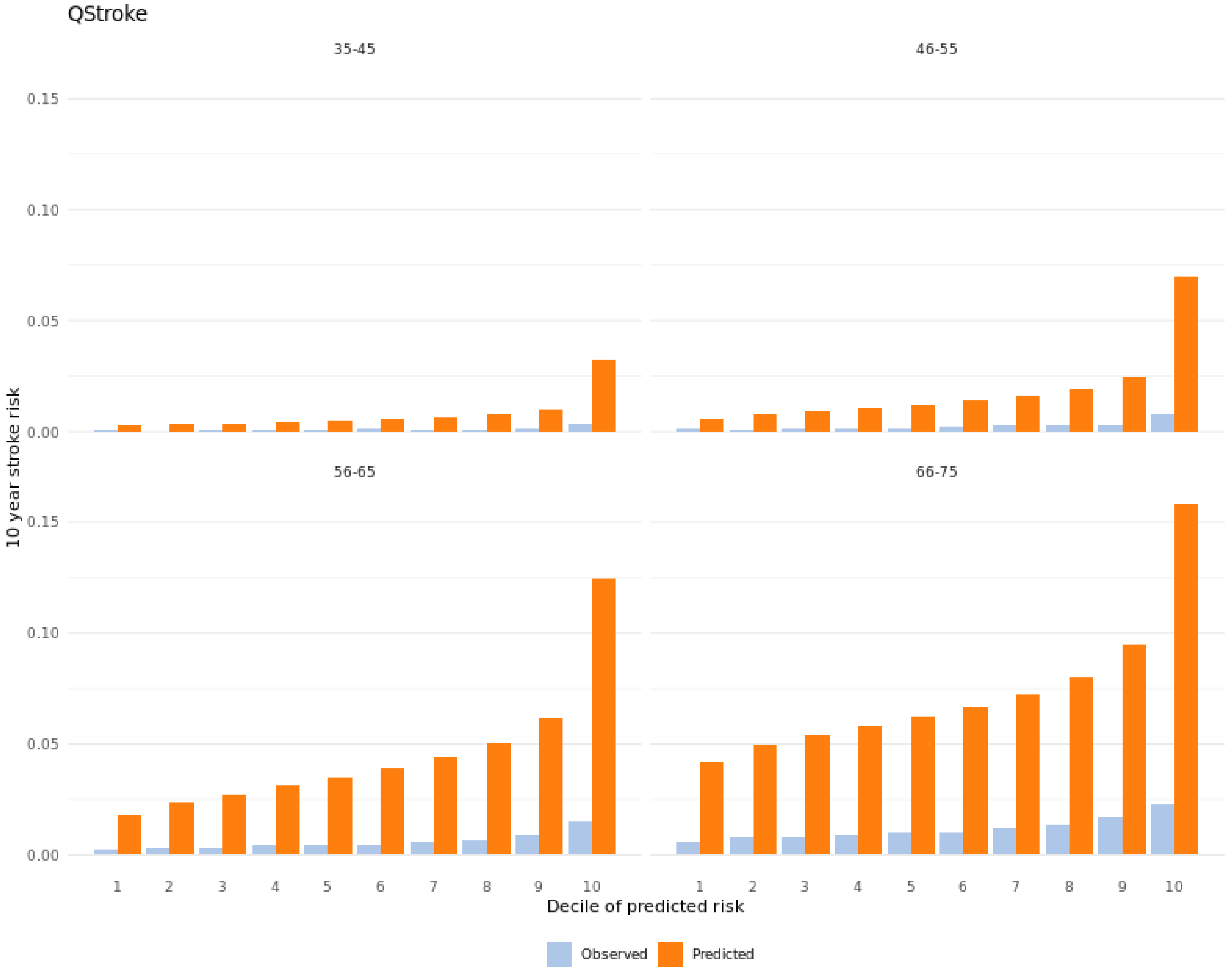
QStroke
35-45
46-55
56-65
66-75
0.15
0.10
0.05
0.00
10 year stroke risk
1
2
3
4
5
6
7
8
9
10
Decile of predicted risk
Observed
Predicted

#### 8.5 Calibration plot based on immigration status

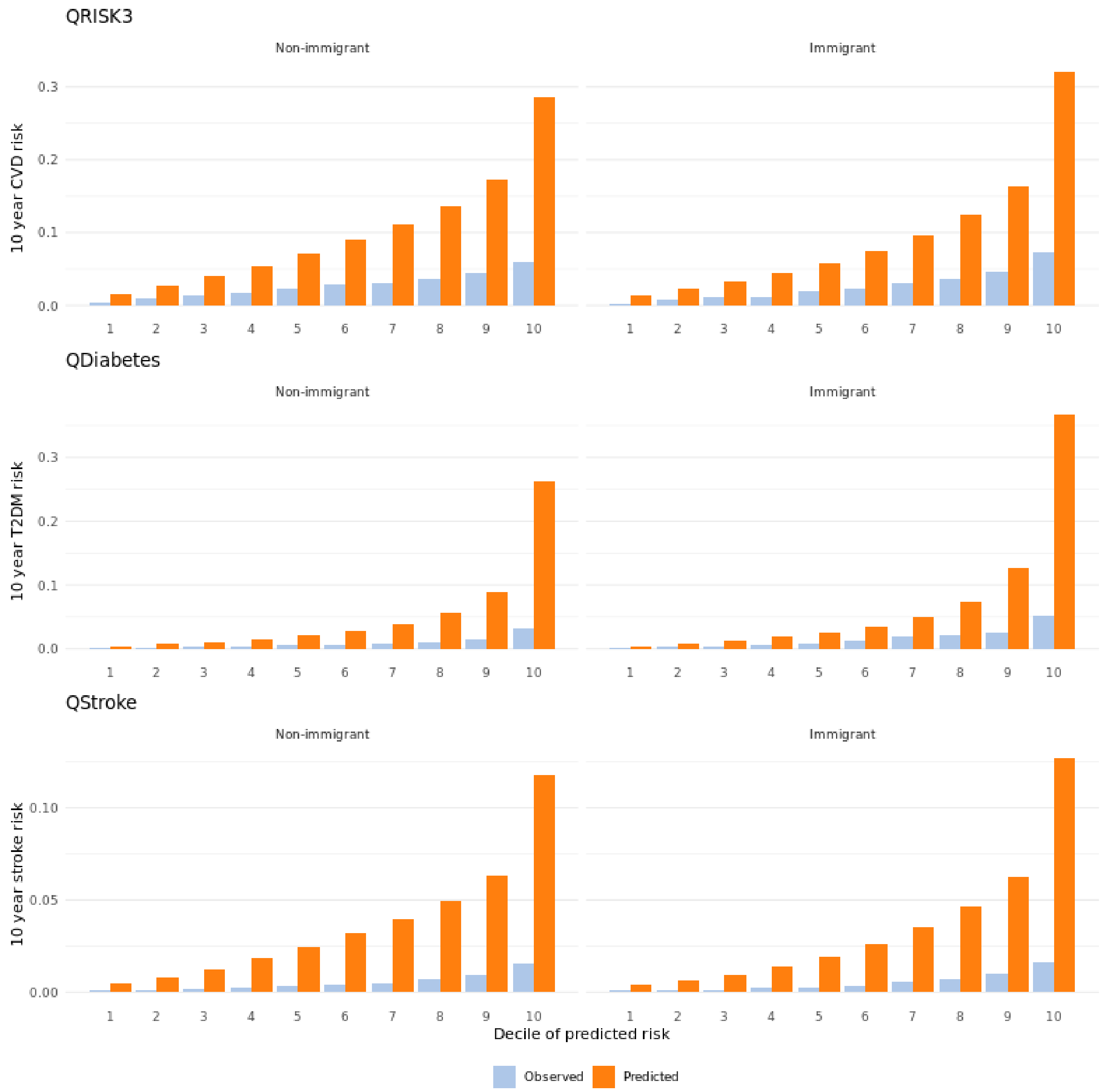

#### 8.6 Calibration plot based on average household income before taxation

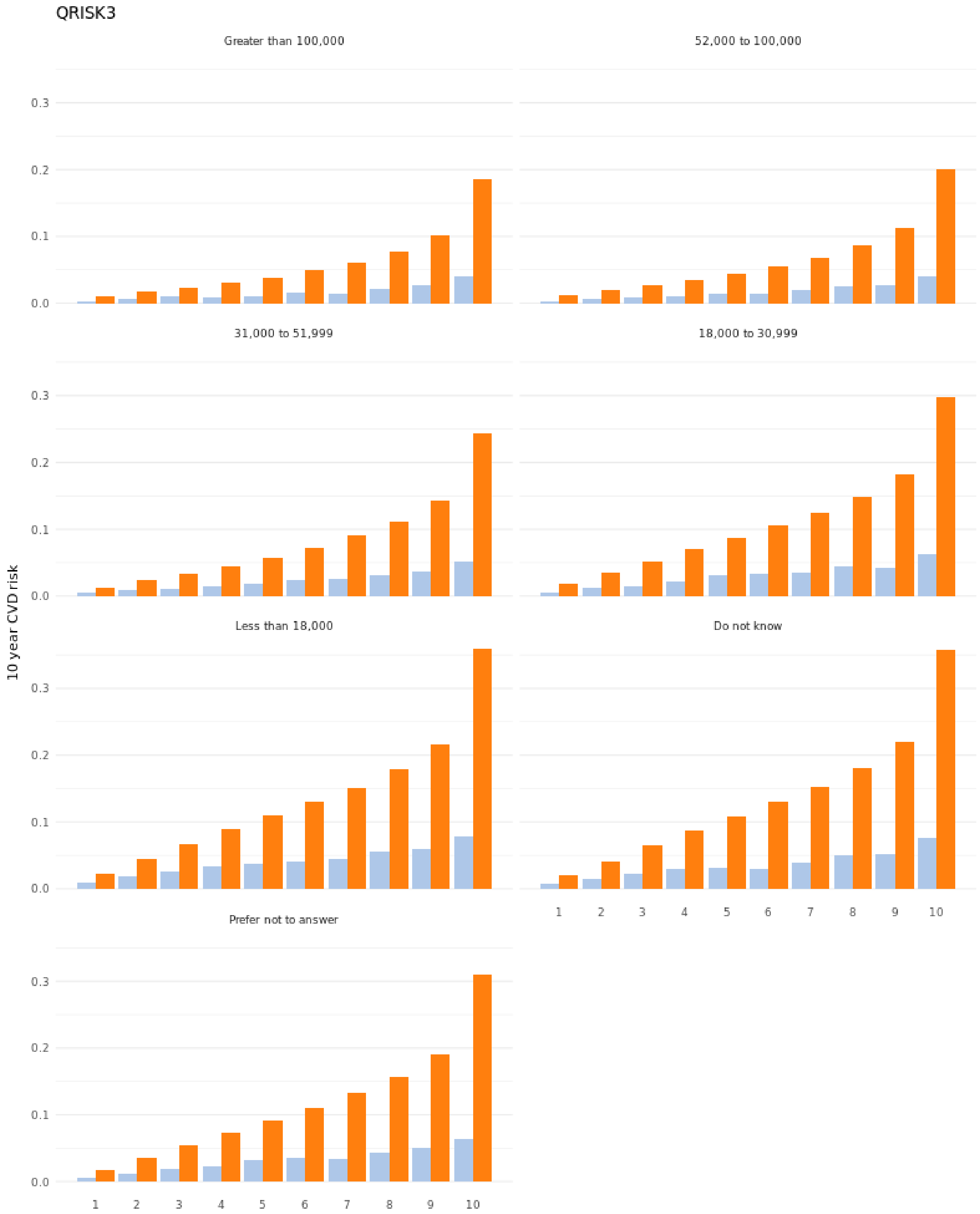

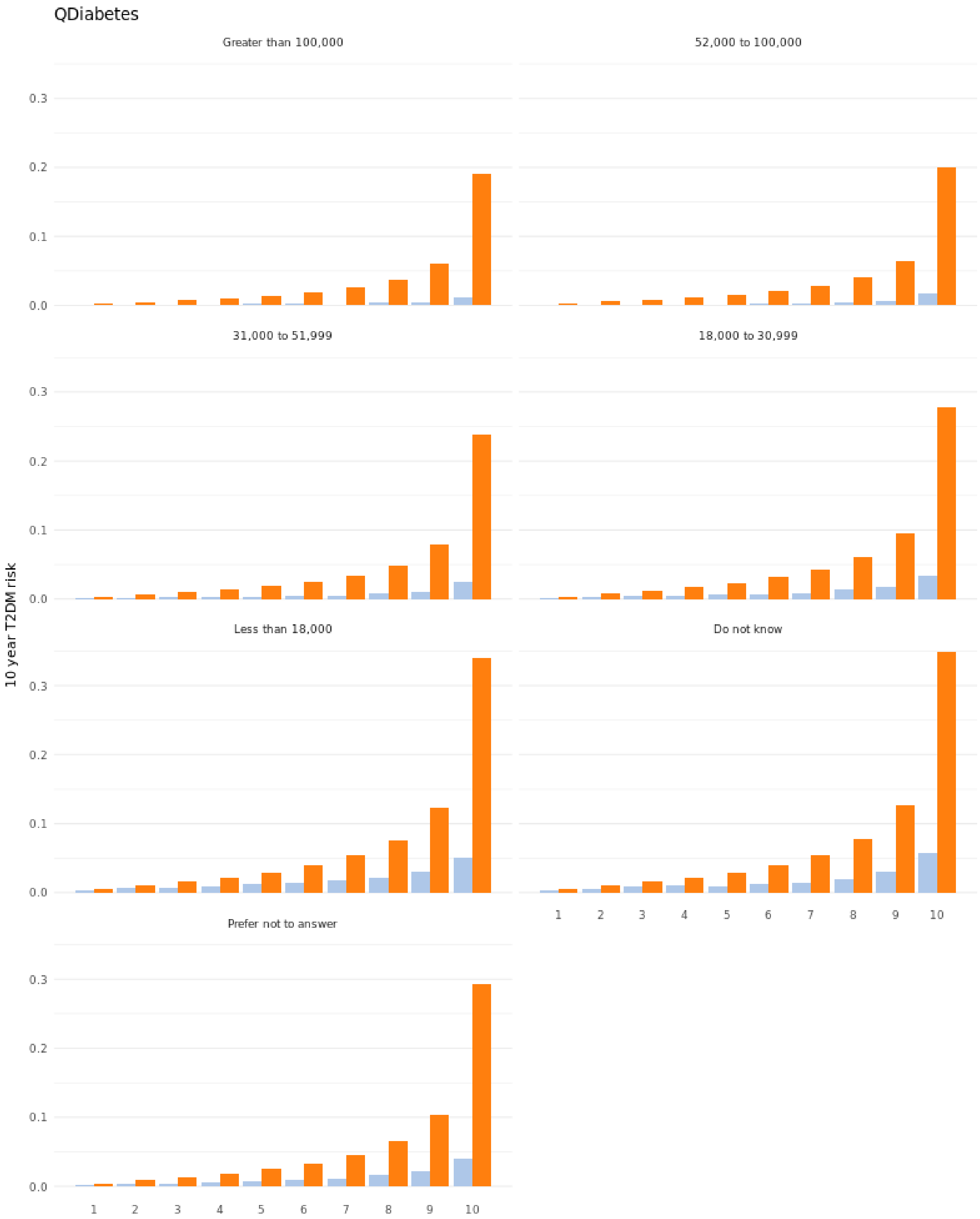
QDiabetes
Greater than 100,000
52,000 to 100,000
31,000 to 51,999
18,000 to 30,999
Less than 18,000
Do not know
Prefer not to answer
10 year T2DM risk
0.0
0.1
0.2
0.3
1
2
3
4
5
6
7
8
9
10

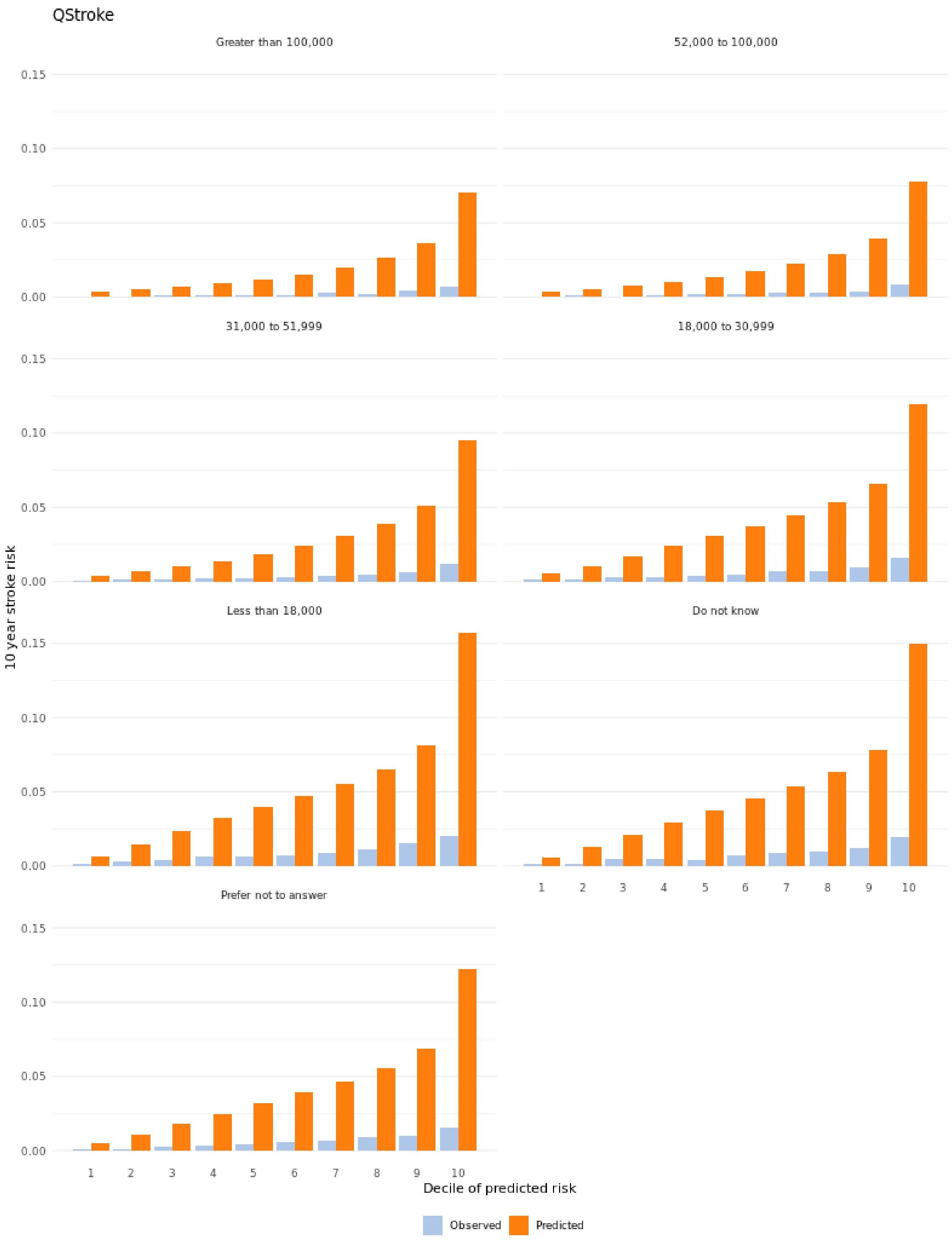
QStroke
Greater than 100,000
52,000 to 100,000
31,000 to 51,999
18,000 to 30,999
Less than 18,000
Do not know
Prefer not to answer
10 year stroke risk
0.00
0.05
0.10
0.15
1
2
3
4
5
6
7
8
9
10
Decile of predicted risk
Observed
Predicted

#### 8.7 Calibration plot based on education level

QRISK3

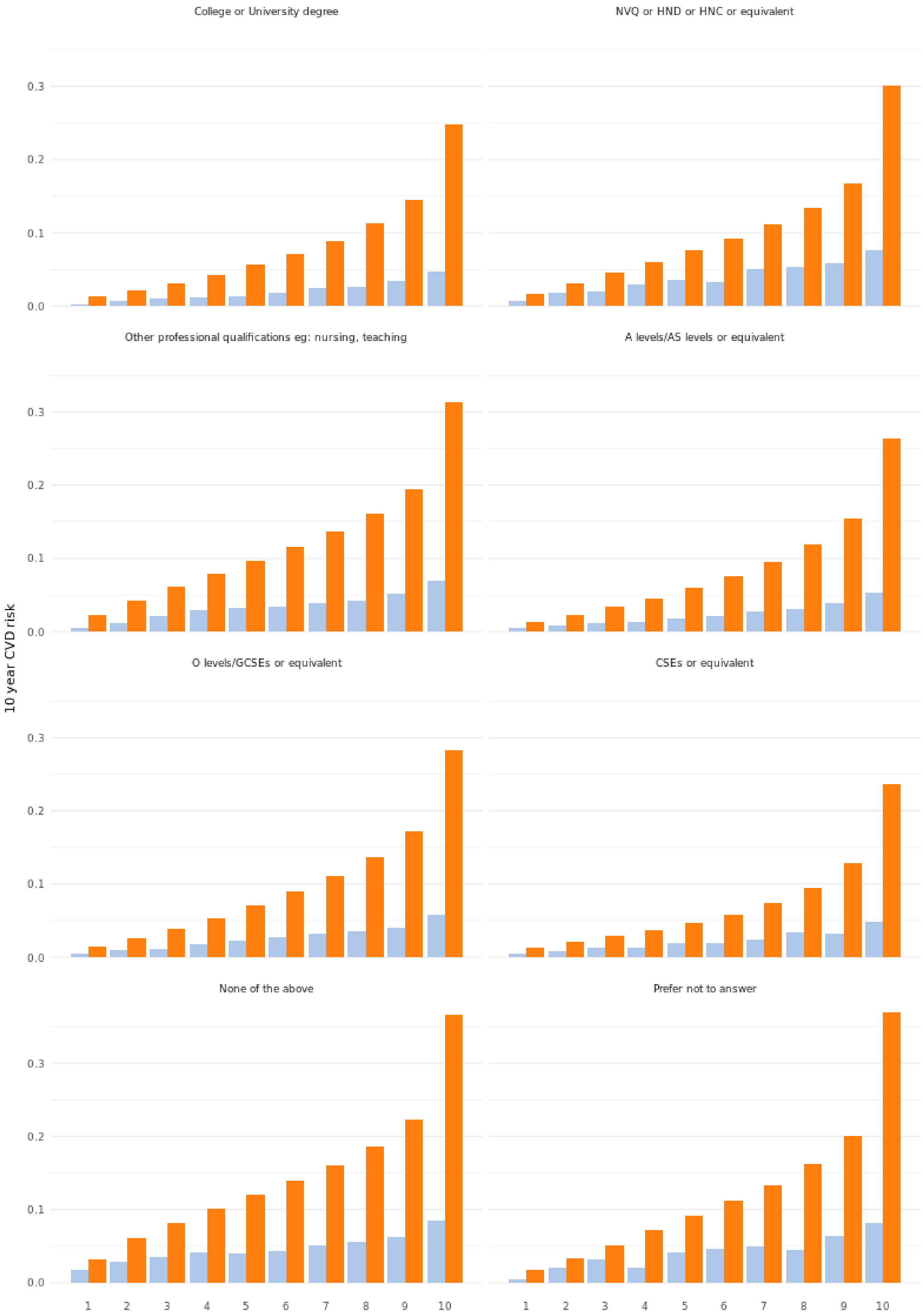

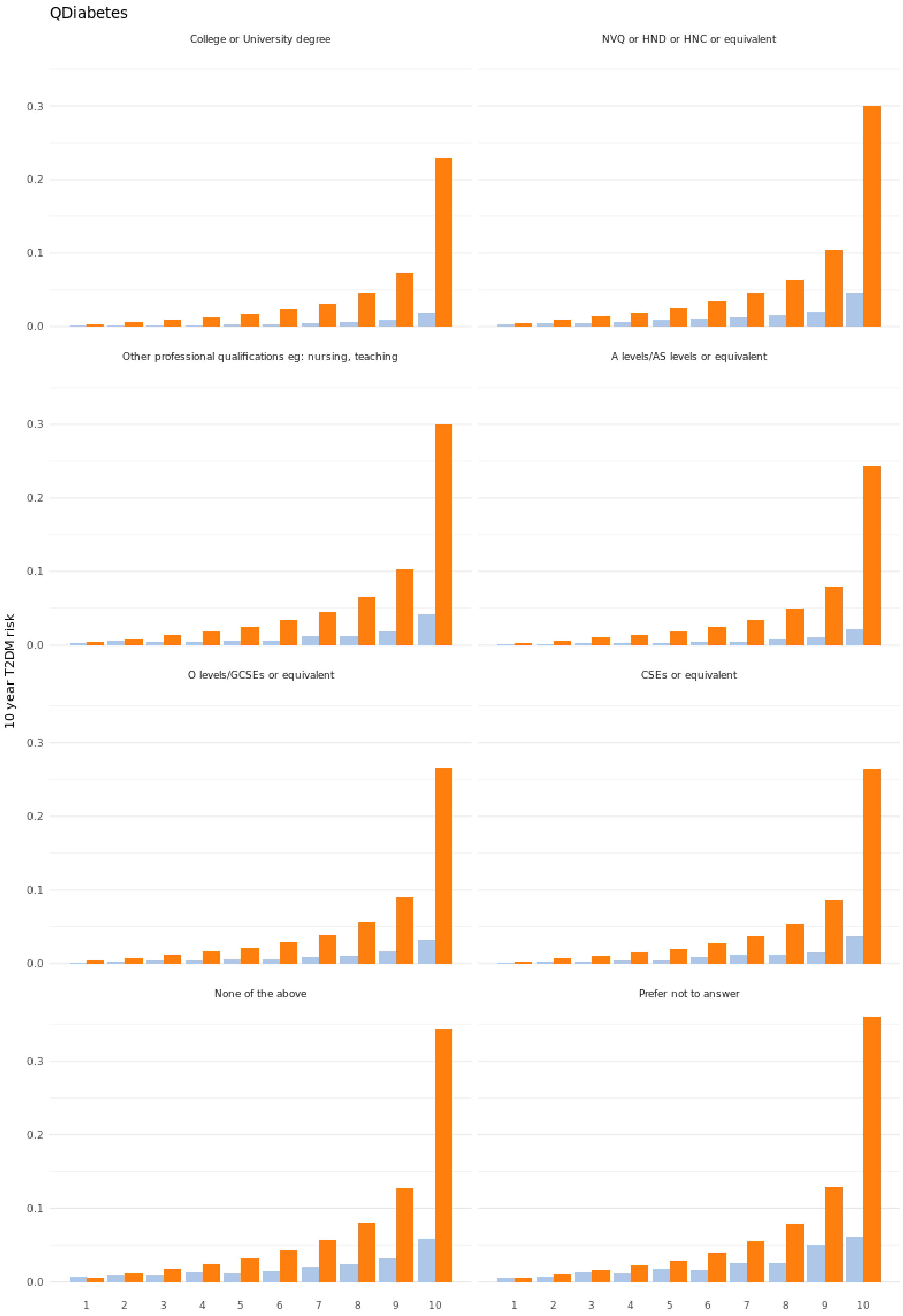
QDiabetes
College or University degree
NVQ or HND or HNC or equivalent
Other professional qualifications eg: nursing, teaching
A levels/AS levels or equivalent
O levels/GCSEs or equivalent
CSEs or equivalent
None of the above
Prefer not to answer
10 year T2DM risk
0.0
0.1
0.2
0.3
1
2
3
4
5
6
7
8
9
10

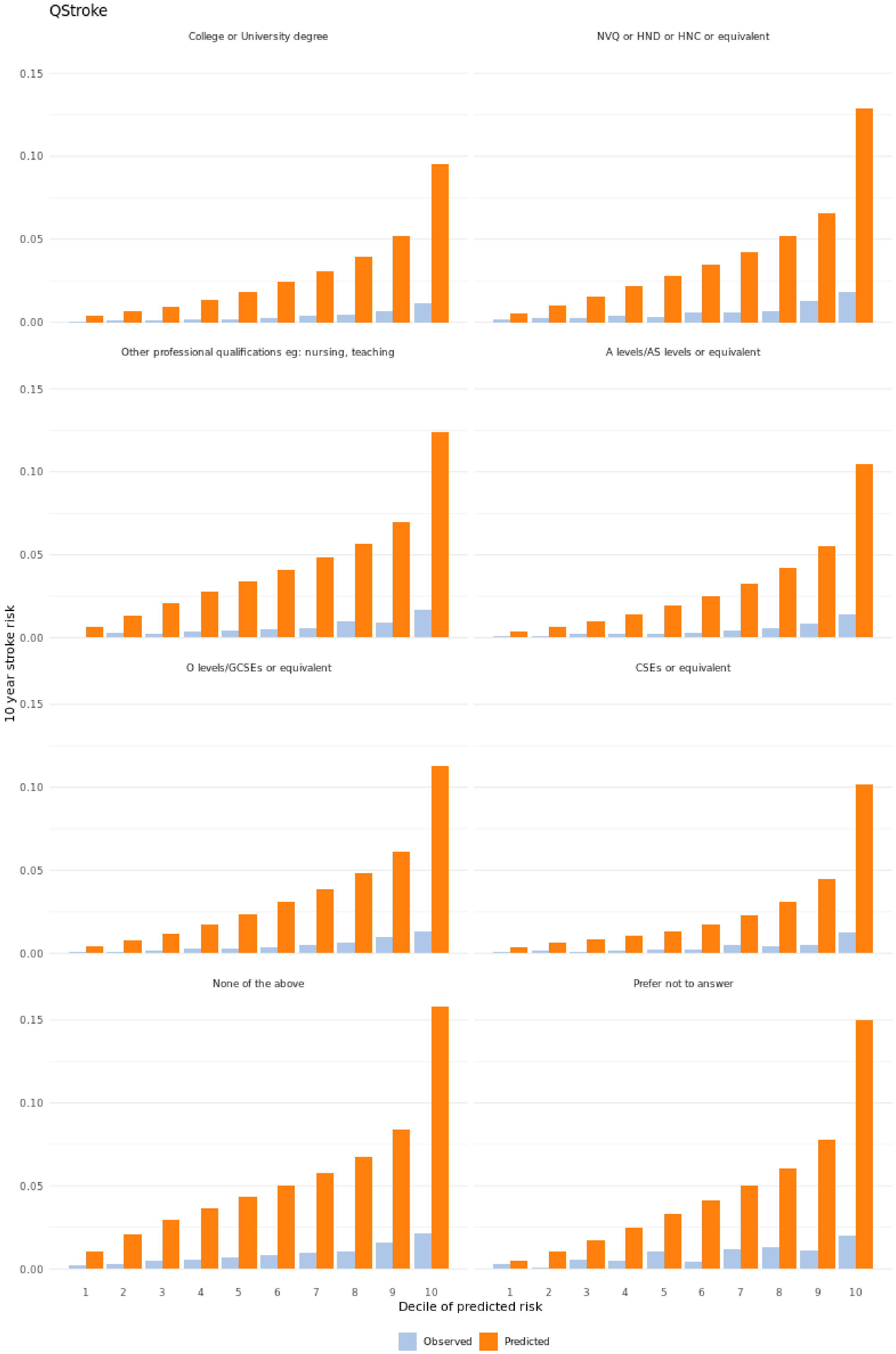
QStroke
College or University degree
NVQ or HND or HNC or equivalent
Other professional qualifications eg: nursing, teaching
A levels/AS levels or equivalent
O levels/GCSEs or equivalent
CSEs or equivalent
None of the above
Prefer not to answer
10 year stroke risk
0.00
0.05
0.10
0.15
1 2 3 4 5 6 7 8 9 10
Decile of predicted risk
Observed
Predicted

#### 8.8 Calibration plot based on Townsend deprivation score

QRISK3

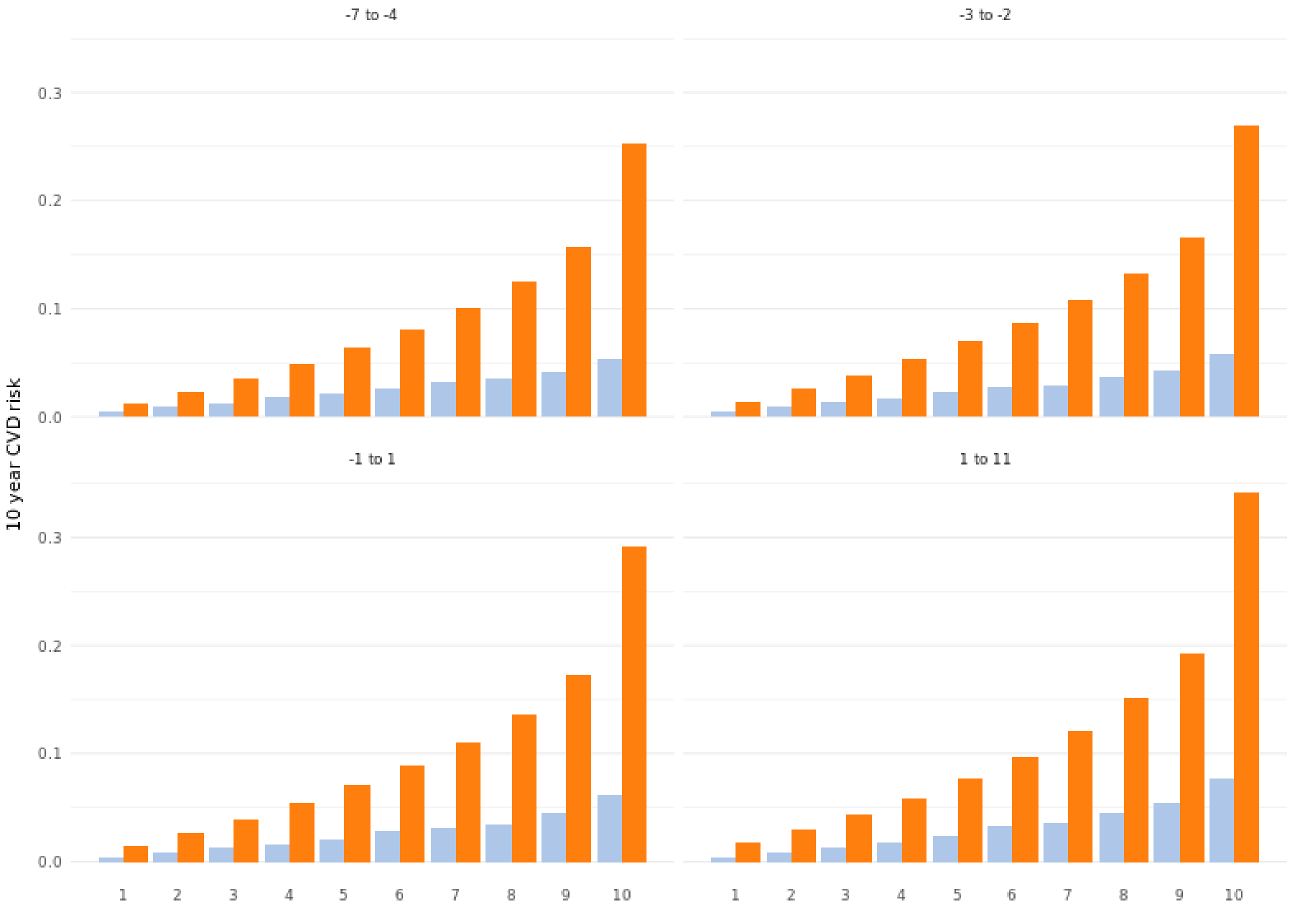

QDiabetes

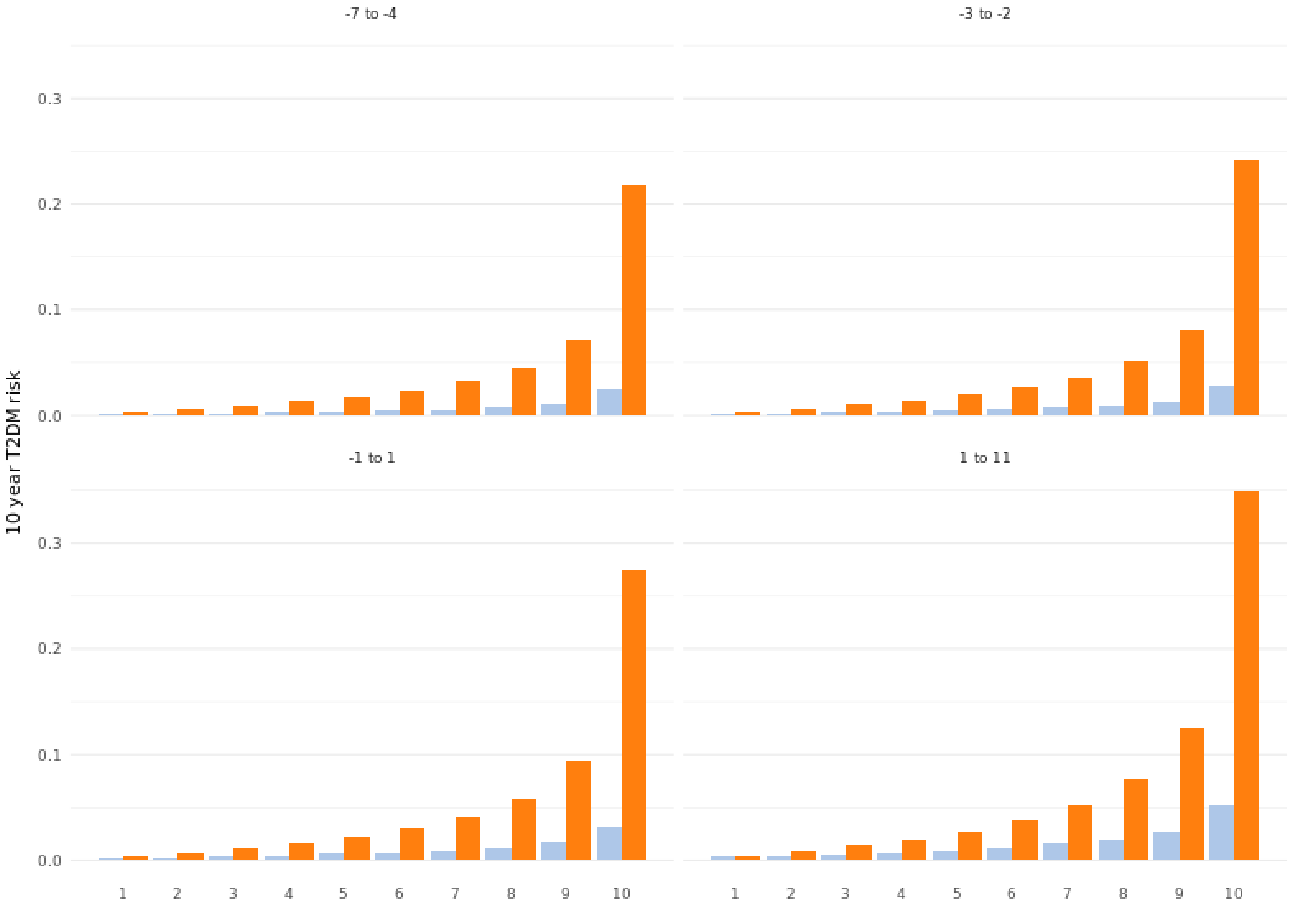

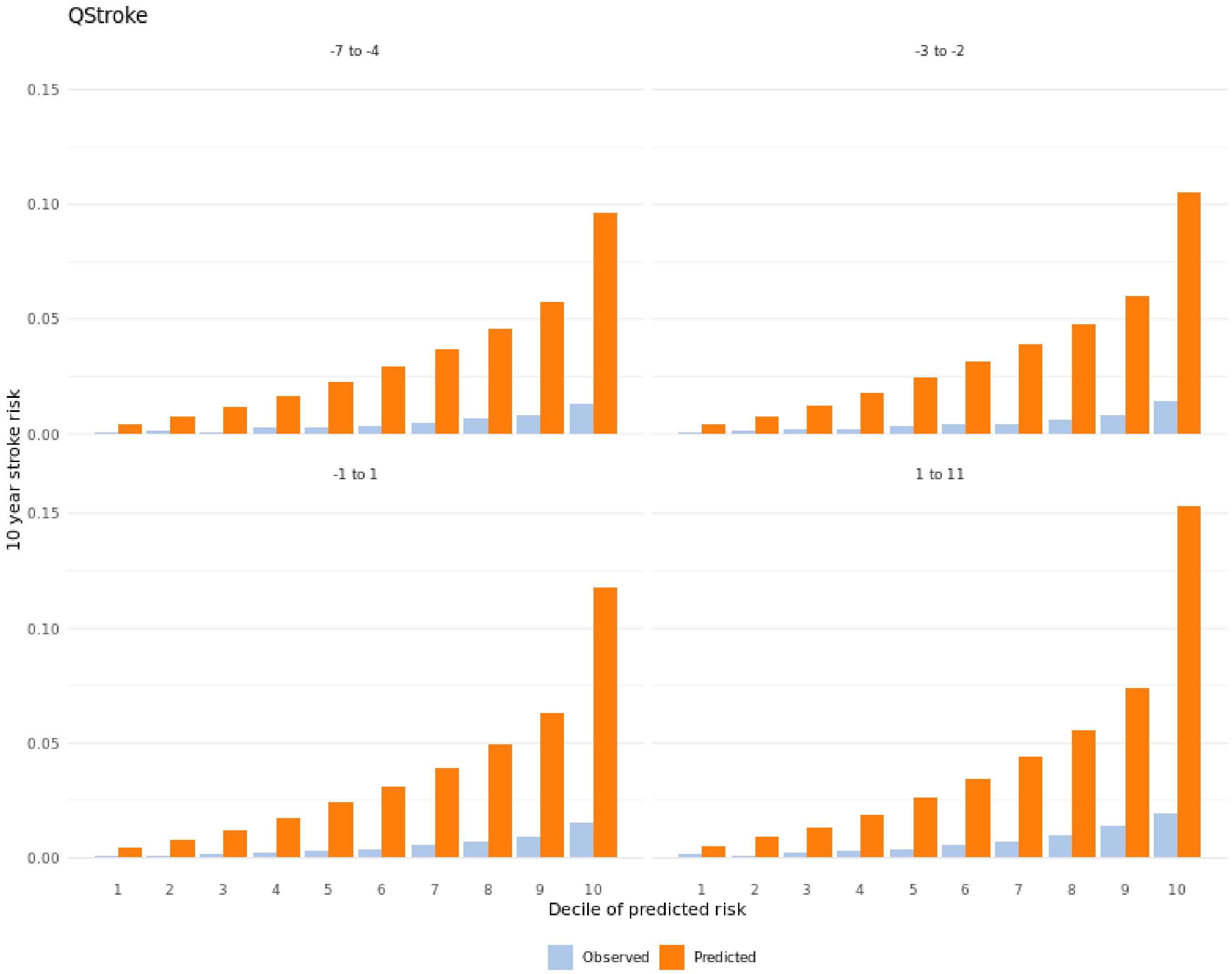
QStroke
-7 to -4
-3 to -2
-1 to 1
1 to 11
0.15
0.10
0.05
0.00
10 year stroke risk
1
2
3
4
5
6
7
8
9
10
Decile of predicted risk
Observed
Predicted

#### 8.9 Calibration plot based on index of multiple deprivation

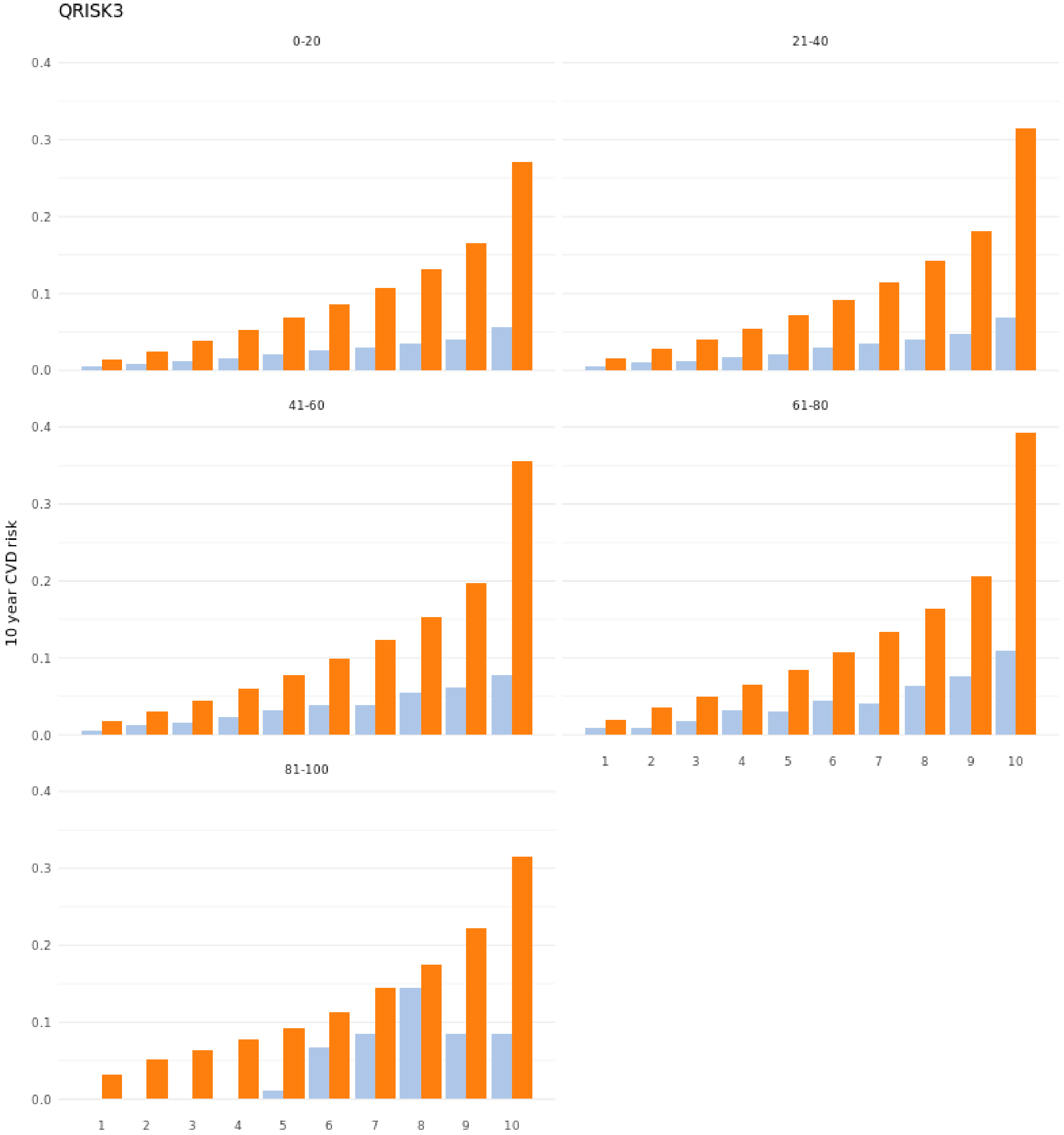

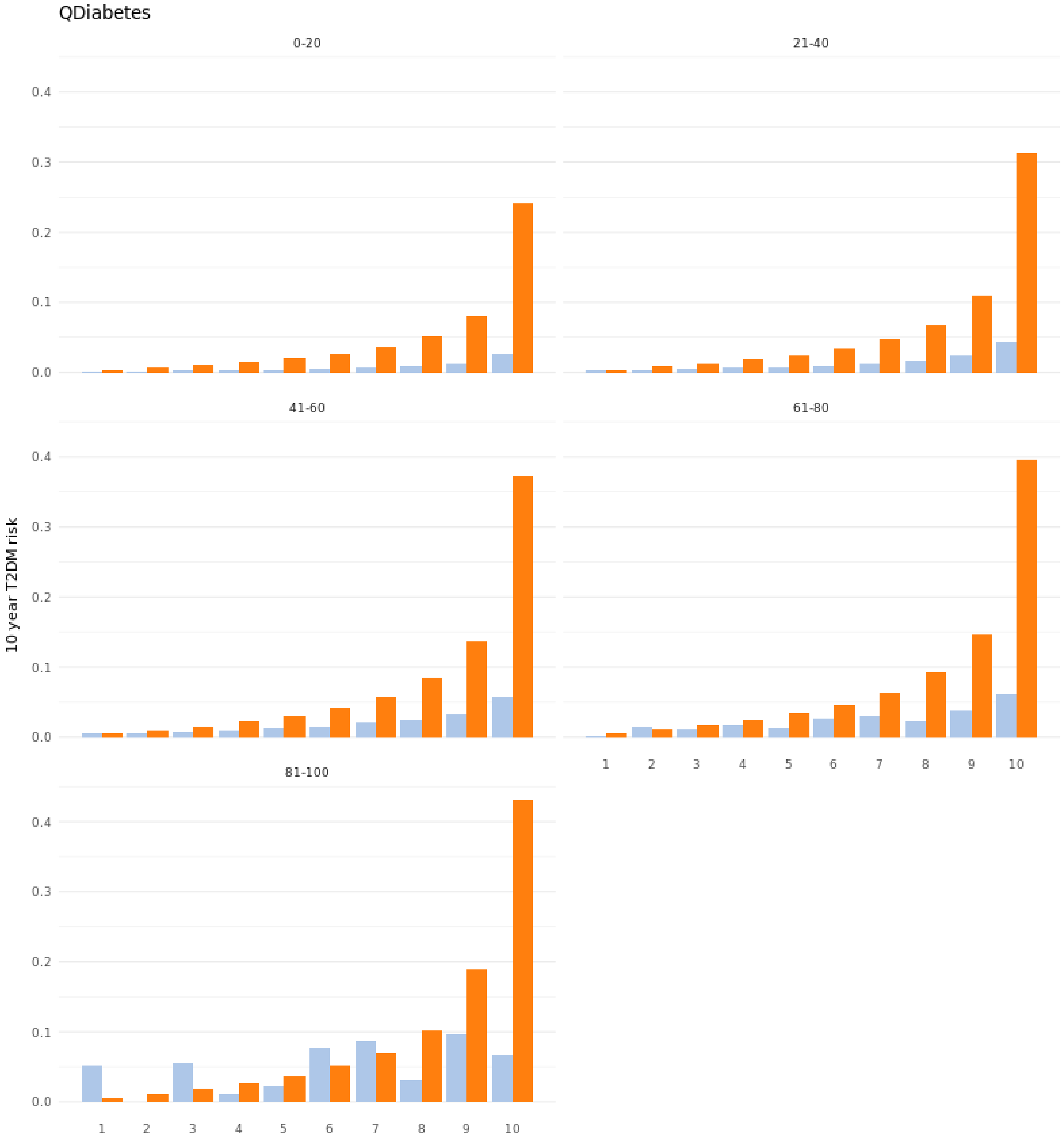
QDiabetes
0-20
21-40
41-60
61-80
81-100
10 year T2DM risk
0.0
0.1
0.2
0.3
0.4
1
2
3
4
5
6
7
8
9
10

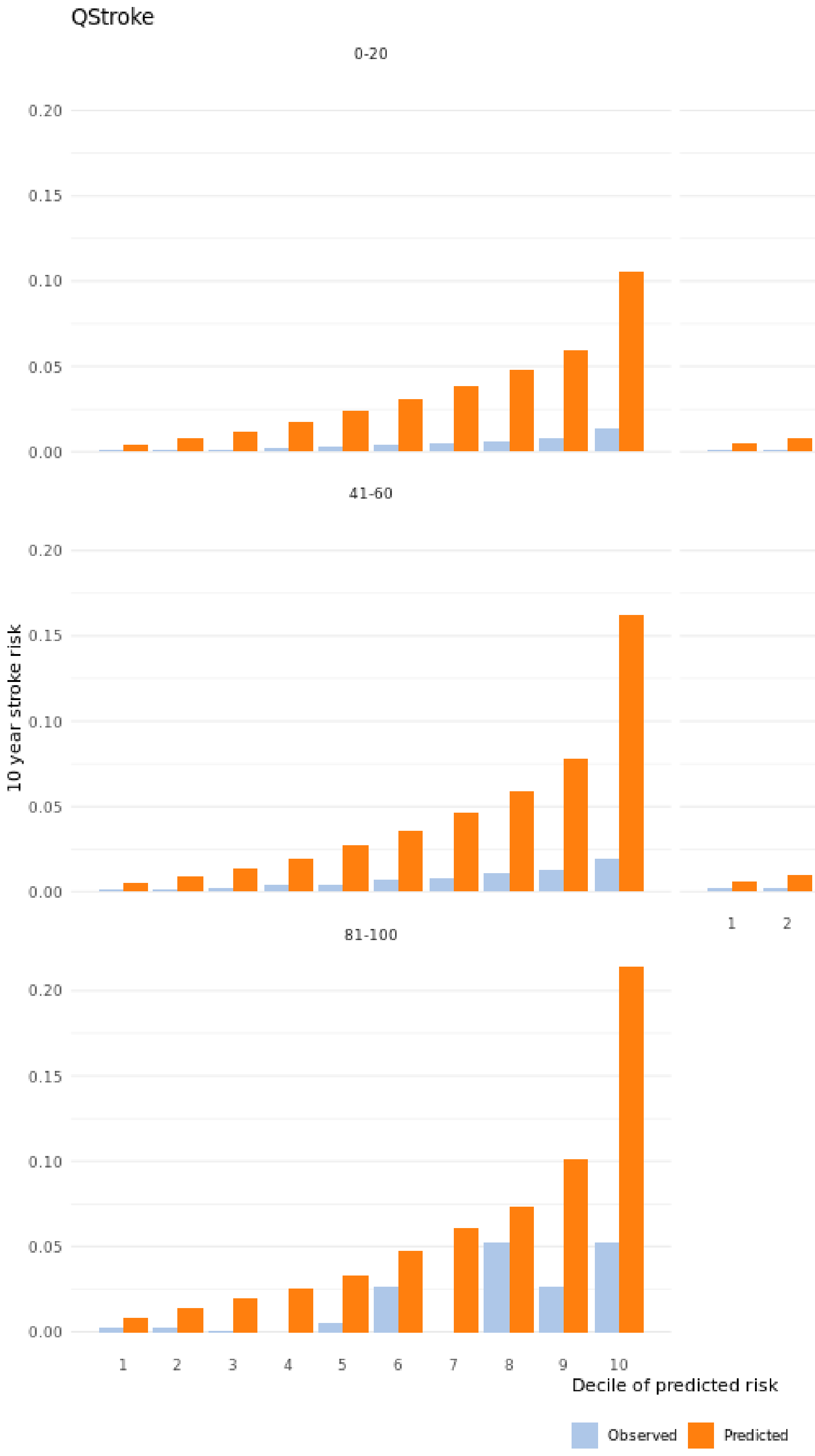

### 9 Calibration, discrimination, and net benefit tables

The tables of sample sizes, survival Brier scores, confusion matrices, and rate parities show the performance of the models based on different fairness metrics.

Survival Brier score is a time-dependent mean squared difference of the predicted and observed values and quantifies the model calibration. The survival Brier scores were not calculated for QStroke as the model's IPCW weighted estimates didn't match the Aalen-Johanssen estimates with any combination of covariates and without.

Confusion matrices show the model discrimination. The thresholds used to dichotomize individual risks were chosen from QPrediction guidelines for clinical use. The rate parities for each metric in the confusion matrix shows the prediction parities. As QStroke has not been recommended by any national guidelines, we omitted the model from the discrimination analysis.

Net benefits (NB) show the model performance as the expected number of true-positive treatments minus a weighted penalty for unnecessary treatment expressed per patient. This is contrasted with the scenarios where all individuals are treated (TrA) and where no-one is treated (TrN).s

All the values are calculated from five MICE imputed datasets and pooled using Rubin's rules. IPCW weights are used to calculate the survival Brier scores and values in the confusion matrices as well as rate parities.

#### 9.1 Table of sample sizes, survival Brier scores, confusion matrices, and rate parities for demographic subgroups for QRISK3

| | QRISK3 | | | | | | | | | |
|---|---|---|---|---|---|---|---|---|---|---|
| | Sample size | Survival brier score | TPR | FPR | TNR | FNR | TPR parity | FPR parity | TNR parity | FNR parity |
| **Overall** | 405855 | 0.78 | 0.63 | 0.38 | 0.62 | 0.37 | 1.00 | 1.00 | 1.00 | 1.00 |
| **Sex** | | | | | | | | | | |
| Male | 234808.0 | 0.77 | 0.78 | 0.46 | 0.54 | 0.22 | Ref | Ref | Ref | Ref |
| Female | 171047.0 | 0.80 | 0.53 | 0.27 | 0.73 | 0.47 | 0.68 | 0.58 | 1.35 | 2.13 |
| **Age group** | | | | | | | | | | |
| [35,45] | 60888.0 | 0.93 | 0.08 | 0.02 | 0.98 | 0.92 | Ref | Ref | Ref | Ref |
| (45,55] | 131177.0 | 0.86 | 0.25 | 0.11 | 0.89 | 0.75 | 3.04 | 6.23 | 0.91 | 0.82 |
| (55,65] | 166011.0 | 0.72 | 0.68 | 0.58 | 0.42 | 0.32 | 8.33 | 32.81 | 0.42 | 0.35 |
| (65,75] | 47779.0 | 0.59 | 0.96 | 0.95 | 0.05 | 0.04 | 11.80 | 53.56 | 0.05 | 0.04 |
| **Ethnicity** | | | | | | | | | | |
| White | 383023.8 | 0.78 | 0.64 | 0.39 | 0.61 | 0.36 | Ref | Ref | Ref | Ref |
| Mixed | 2533.6 | 0.85 | 0.38 | 0.17 | 0.83 | 0.62 | 0.60 | 0.44 | 1.35 | 1.71 |
| Other ethnic group | 3675.8 | 0.84 | 0.44 | 0.19 | 0.81 | 0.56 | 0.69 | 0.49 | 1.33 | 1.54 |
| Prefer not to answer | 1300.2 | 0.79 | 0.47 | 0.29 | 0.71 | 0.53 | 0.73 | 0.74 | 1.17 | 1.47 |
| Chinese | 1385.0 | 0.89 | 0.41 | 0.10 | 0.90 | 0.59 | 0.64 | 0.26 | 1.47 | 1.64 |
| Black or Black British | 6649.2 | 0.86 | 0.46 | 0.15 | 0.85 | 0.54 | 0.72 | 0.38 | 1.39 | 1.49 |
| Asian or Asian British | 7119.4 | 0.75 | 0.65 | 0.36 | 0.64 | 0.35 | 1.02 | 0.92 | 1.05 | 0.97 |
| Do not know | 168.0 | 0.79 | 0.33 | 0.22 | 0.78 | 0.67 | 0.52 | 0.58 | 1.27 | 1.85 |
| **Immigration status** | | | | | | | | | | |
| No | 369896.0 | 0.78 | 0.63 | 0.39 | 0.61 | 0.37 | Ref | Ref | Ref | Ref |
| Yes | 35959.0 | 0.80 | 0.62 | 0.32 | 0.68 | 0.38 | 0.97 | 0.83 | 1.11 | 1.05 |
| **Average household income** | | | | | | | | | | |
| Greater than 100,000 | 20479.6 | 0.86 | 0.35 | 0.15 | 0.85 | 0.65 | Ref | Ref | Ref | Ref |
| 52,000 to 100,000 | 76868.6 | 0.85 | 0.38 | 0.19 | 0.81 | 0.62 | 1.09 | 1.25 | 0.96 | 0.95 |
| 31,000 to 51,999 | 95015.6 | 0.81 | 0.52 | 0.29 | 0.71 | 0.48 | 1.51 | 1.95 | 0.83 | 0.73 |
| 18,000 to 30,999 | 86649.6 | 0.76 | 0.69 | 0.46 | 0.54 | 0.31 | 1.98 | 3.12 | 0.63 | 0.48 |
| Less than 18,000 | 70776.8 | 0.71 | 0.77 | 0.58 | 0.42 | 0.23 | 2.24 | 3.92 | 0.49 | 0.35 |
| Prefer not to answer | 39660.6 | 0.75 | 0.70 | 0.49 | 0.51 | 0.30 | 2.03 | 3.31 | 0.60 | 0.46 |
| Do not know | 16404.2 | 0.72 | 0.77 | 0.58 | 0.42 | 0.23 | 2.24 | 3.90 | 0.49 | 0.35 |
| **Highest education level** | | | | | | | | | | |
| College or University degree | 139399.4 | 0.82 | 0.54 | 0.29 | 0.71 | 0.46 | Ref | Ref | Ref | Ref |
| NVQ or HND or HNC or equivalent | 25162.8 | 0.76 | 0.63 | 0.39 | 0.61 | 0.37 | 1.16 | 1.35 | 0.86 | 0.81 |
| Other professional qualifications eg: nursing, teaching | 20154.0 | 0.74 | 0.73 | 0.52 | 0.48 | 0.27 | 1.34 | 1.80 | 0.67 | 0.60 |
| A levels/AS levels or equivalent | 47187.4 | 0.81 | 0.56 | 0.32 | 0.68 | 0.44 | 1.04 | 1.10 | 0.96 | 0.95 |
| O levels/GCSEs or equivalent | 87101.8 | 0.78 | 0.62 | 0.39 | 0.61 | 0.38 | 1.15 | 1.33 | 0.86 | 0.82 |
| CSEs or equivalent | 23220.4 | 0.83 | 0.41 | 0.22 | 0.78 | 0.59 | 0.75 | 0.77 | 1.10 | 1.29 |
| None of the above | 59587.2 | 0.69 | 0.78 | 0.65 | 0.35 | 0.22 | 1.44 | 2.23 | 0.50 | 0.48 |
| Prefer not to answer | 4042.0 | 0.73 | 0.71 | 0.50 | 0.50 | 0.29 | 1.30 | 1.71 | 0.71 | 0.64 |
| **Townsend deprivation score (quantiles)** | | | | | | | | | | |
| [-7,-4] | 78670 | 0.80 | 0.58 | 0.34 | 0.66 | 0.42 | Ref | Ref | Ref | Ref |
| (-4,-2] | 135751 | 0.79 | 0.61 | 0.38 | 0.62 | 0.39 | 1.05 | 1.09 | 0.95 | 0.93 |
| (-2,1] | 105950 | 0.78 | 0.64 | 0.38 | 0.62 | 0.36 | 1.11 | 1.12 | 0.94 | 0.85 |
| (1,11] | 85484 | 0.76 | 0.70 | 0.42 | 0.58 | 0.30 | 1.22 | 1.22 | 0.89 | 0.70 |
| **Index of multiple deprivation** | | | | | | | | | | |
| [0,20] | 285207.8 | 0.79 | 0.61 | 0.37 | 0.63 | 0.39 | Ref | Ref | Ref | Ref |
| (20,40] | 88085.6 | 0.77 | 0.66 | 0.40 | 0.60 | 0.34 | 1.08 | 1.07 | 0.96 | 0.88 |
| (40,60] | 27027 | 0.75 | 0.68 | 0.43 | 0.57 | 0.32 | 1.10 | 1.16 | 0.91 | 0.84 |
| (60,80] | 5410.2 | 0.73 | 0.75 | 0.46 | 0.54 | 0.25 | 1.22 | 1.25 | 0.85 | 0.65 |
| (80,100] | 124.4 | 0.74 | 0.97 | 0.46 | 0.54 | 0.03 | 1.57 | 1.25 | 0.85 | 0.09 |

#### 9.2 Table of sample sizes, survival brier scores, confusion matrices, and rate parities for demographic subgroups for QDiabetes

| | QDiabetes | | | | | | | | | |
|---|---|---|---|---|---|---|---|---|---|---|
| | Sample size | Survival brier score | TPR | FPR | TNR | FNR | TPR parity | FPR parity | TNR parity | FNR parity |
| **Overall** | 458700 | 0.90 | 0.59 | 0.24 | 0.76 | 0.41 | 1.00 | 1.00 | 1.00 | 1.00 |
| **Sex** | | | | | | | | | | |
| Male | 254758.0 | 0.90 | 0.65 | 0.26 | 0.74 | 0.35 | Ref | Ref | Ref | Ref |
| Female | 203942.0 | 0.91 | 0.54 | 0.22 | 0.78 | 0.46 | 0.84 | 0.86 | 1.05 | 1.30 |
| **Age group** | | | | | | | | | | |
| [35,45] | 60960.0 | 0.95 | 0.48 | 0.11 | 0.89 | 0.52 | Ref | Ref | Ref | Ref |
| (45,55] | 136384.0 | 0.92 | 0.58 | 0.19 | 0.81 | 0.42 | 1.21 | 1.72 | 0.91 | 0.81 |
| (55,65] | 196067.0 | 0.89 | 0.61 | 0.29 | 0.71 | 0.39 | 1.29 | 2.56 | 0.80 | 0.74 |
| (65,75] | 65289.0 | 0.87 | 0.60 | 0.33 | 0.67 | 0.40 | 1.27 | 2.93 | 0.75 | 0.76 |
| **Ethnicity** | | | | | | | | | | |
| White | 434852.4 | 0.91 | 0.58 | 0.23 | 0.77 | 0.42 | Ref | Ref | Ref | Ref |
| Mixed | 2671.6 | 0.85 | 0.65 | 0.39 | 0.61 | 0.35 | 1.12 | 1.67 | 0.80 | 0.84 |
| Other ethnic group | 3849.6 | 0.83 | 0.78 | 0.44 | 0.56 | 0.22 | 1.34 | 1.89 | 0.73 | 0.53 |
| Prefer not to answer | 1455.2 | 0.82 | 0.81 | 0.47 | 0.53 | 0.19 | 1.40 | 2.01 | 0.69 | 0.45 |
| Chinese | 1406.0 | 0.89 | 0.54 | 0.30 | 0.70 | 0.46 | 0.93 | 1.30 | 0.91 | 1.10 |
| Black or Black British | 6748.6 | 0.79 | 0.77 | 0.52 | 0.48 | 0.23 | 1.33 | 2.24 | 0.62 | 0.54 |
| Asian or Asian British | 7532.6 | 0.85 | 0.61 | 0.36 | 0.64 | 0.39 | 1.06 | 1.53 | 0.84 | 0.92 |
| Do not know | 184.0 | 0.83 | 0.70 | 0.46 | 0.54 | 0.30 | 1.21 | 1.97 | 0.71 | 0.72 |
| **Immigration status** | | | | | | | | | | |
| No | 419853.0 | 0.90 | 0.58 | 0.24 | 0.76 | 0.42 | Ref | Ref | Ref | Ref |
| Yes | 38847.0 | 0.88 | 0.65 | 0.31 | 0.69 | 0.35 | 1.11 | 1.29 | 0.91 | 0.85 |
| **Average household income** | | | | | | | | | | |
| Greater than 100,000 | 21976.0 | 0.93 | 0.51 | 0.16 | 0.84 | 0.49 | Ref | Ref | Ref | Ref |
| 52,000 to 100,000 | 81948.0 | 0.93 | 0.55 | 0.17 | 0.83 | 0.45 | 1.08 | 1.08 | 0.98 | 0.92 |
| 31,000 to 51,999 | 103959.8 | 0.91 | 0.53 | 0.21 | 0.79 | 0.47 | 1.05 | 1.32 | 0.94 | 0.95 |
| 18,000 to 30,999 | 99476.4 | 0.90 | 0.58 | 0.26 | 0.74 | 0.42 | 1.14 | 1.65 | 0.88 | 0.85 |
| Less than 18,000 | 86526.4 | 0.88 | 0.62 | 0.32 | 0.68 | 0.38 | 1.23 | 2.05 | 0.80 | 0.76 |
| Prefer not to answer | 45778.0 | 0.89 | 0.61 | 0.28 | 0.72 | 0.39 | 1.20 | 1.75 | 0.86 | 0.79 |
| Do not know | 19035.4 | 0.87 | 0.65 | 0.33 | 0.67 | 0.35 | 1.28 | 2.07 | 0.80 | 0.72 |
| **Highest education level** | | | | | | | | | | |
| College or University degree | 151750.8 | 0.92 | 0.56 | 0.19 | 0.81 | 0.44 | Ref | Ref | Ref | Ref |
| NVQ or HND or HNC or equivalent | 29505.2 | 0.89 | 0.59 | 0.27 | 0.73 | 0.41 | 1.07 | 1.42 | 0.90 | 0.91 |
| Other professional qualifications eg: nursing, teaching | 23437.4 | 0.89 | 0.62 | 0.28 | 0.72 | 0.38 | 1.11 | 1.45 | 0.89 | 0.86 |
| A levels/AS levels or equivalent | 51475.4 | 0.91 | 0.53 | 0.21 | 0.79 | 0.47 | 0.95 | 1.10 | 0.98 | 1.06 |
| O levels/GCSEs or equivalent | 97356.8 | 0.90 | 0.57 | 0.24 | 0.76 | 0.43 | 1.03 | 1.25 | 0.94 | 0.96 |
| CSEs or equivalent | 25010.2 | 0.91 | 0.58 | 0.23 | 0.77 | 0.42 | 1.04 | 1.19 | 0.95 | 0.95 |
| None of the above | 75393.0 | 0.87 | 0.63 | 0.34 | 0.66 | 0.37 | 1.14 | 1.78 | 0.81 | 0.82 |
| Prefer not to answer | 4771.2 | 0.87 | 0.63 | 0.33 | 0.67 | 0.37 | 1.14 | 1.71 | 0.83 | 0.82 |
| **Townsend deprivation score (quantiles)** | | | | | | | | | | |
| [-7,-4] | 88168.0 | 0.92 | 0.53 | 0.19 | 0.81 | 0.47 | Ref | Ref | Ref | Ref |
| (-4,-2] | 153310.0 | 0.91 | 0.55 | 0.22 | 0.78 | 0.45 | 1.04 | 1.16 | 0.96 | 0.96 |
| (-2,1] | 119538.0 | 0.90 | 0.59 | 0.25 | 0.75 | 0.41 | 1.11 | 1.33 | 0.92 | 0.88 |
| (1,11] | 97684.0 | 0.88 | 0.65 | 0.32 | 0.68 | 0.35 | 1.23 | 1.69 | 0.84 | 0.74 |
| **Index of multiple deprivation** | | | | | | | | | | |
| [0,20] | 320227.4 | 0.91 | 0.56 | 0.22 | 0.78 | 0.44 | Ref | Ref | Ref | Ref |
| (20,40] | 100400.4 | 0.89 | 0.64 | 0.29 | 0.71 | 0.36 | 1.14 | 1.33 | 0.91 | 0.82 |
| (40,60] | 31428.0 | 0.87 | 0.64 | 0.34 | 0.66 | 0.36 | 1.16 | 1.59 | 0.84 | 0.80 |
| (60,80] | 6473.2 | 0.86 | 0.62 | 0.37 | 0.63 | 0.38 | 1.11 | 1.71 | 0.80 | 0.86 |
| (80,100] | 171.0 | 0.84 | 0.67 | 0.40 | 0.60 | 0.33 | 1.21 | 1.85 | 0.76 | 0.74 |

#### 9.3 Table of net benefits for QRISK3 and QDiabetes

| | QRISK3 | | | | QDiabetes | | | |
|---|---|---|---|---|---|---|---|---|
| | NB | TrN | TrA | NI avoided | NB | TrN | TrA | NI avoided |
| **Overall** | -0.008 | 0 | 0.33 | -3.01 | -0.003 | 0 | 0.17 | -1.525 |
| **Sex** | | | | | | | | |
| Female | 0.009 | 0 | 0.21 | -1.78 | 0.000 | 0 | 0.15 | -1.314 |
| Male | -0.021 | 0 | 0.41 | -3.90 | -0.005 | 0 | 0.18 | -1.694 |
| **Age group** | | | | | | | | |
| [35,45] | -0.001 | 0 | -0.09 | 0.81 | -0.002 | 0 | 0.02 | -0.184 |
| (45,55] | -0.004 | 0 | 0.02 | -0.19 | -0.003 | 0 | 0.11 | -1.023 |
| (55,65] | -0.017 | 0 | 0.54 | -5.05 | -0.004 | 0 | 0.22 | -2.000 |
| (65,75] | 0.005 | 0 | 0.95 | -8.48 | 0.000 | 0 | 0.27 | -2.393 |
| **Ethnicity** | | | | | | | | |
| White | -0.009 | 0 | 0.34 | -3.10 | -0.003 | 0 | 0.16 | -1.426 |
| Mixed | -0.001 | 0 | 0.09 | -0.84 | -0.010 | 0 | 0.33 | -3.029 |
| Other ethnic group | -0.002 | 0 | 0.11 | -1.01 | 0.004 | 0 | 0.39 | -3.510 |
| Prefer not to answer | 0.003 | 0 | 0.22 | -1.98 | -0.007 | 0 | 0.42 | -3.834 |
| Chinese | -0.004 | 0 | 0.01 | -0.10 | -0.006 | 0 | 0.23 | -2.143 |
| Black or Black British | 0.000 | 0 | 0.07 | -0.61 | 0.008 | 0 | 0.48 | -4.273 |
| Asian or Asian British | 0.013 | 0 | 0.31 | -2.68 | 0.016 | 0 | 0.30 | -2.572 |
| Do not know | 0.001 | 0 | 0.15 | -1.32 | 0.005 | 0 | 0.41 | -3.651 |
| **Immigration status** | | | | | | | | |
| No | -0.008 | 0 | 0.33 | -3.07 | -0.003 | 0 | 0.16 | -1.467 |
| Yes | -0.003 | 0 | 0.26 | -2.38 | 0.003 | 0 | 0.24 | -2.148 |
| **Average household income** | | | | | | | | |
| Greater than 100,000 | -0.006 | 0 | 0.06 | -0.60 | -0.001 | 0 | 0.20 | -1.850 |
| 52,000 to 100,000 | -0.008 | 0 | 0.10 | -0.99 | -0.006 | 0 | 0.08 | -0.788 |
| 31,000 to 51,999 | -0.009 | 0 | 0.22 | -2.08 | -0.005 | 0 | 0.13 | -1.180 |
| 18,000 to 30,999 | -0.010 | 0 | 0.42 | -3.86 | -0.003 | 0 | 0.18 | -1.694 |
| Less than 18,000 | -0.003 | 0 | 0.55 | -4.99 | 0.003 | 0 | 0.26 | -2.307 |
| Prefer not to answer | -0.011 | 0 | 0.45 | -4.14 | -0.006 | 0 | 0.07 | -0.656 |
| Do not know | -0.011 | 0 | 0.55 | -5.02 | 0.002 | 0 | 0.26 | -2.352 |
| **Highest education level** | | | | | | | | |
| College or University degree | -0.010 | 0 | 0.22 | -2.09 | -0.006 | 0 | 0.11 | -1.024 |
| NVQ or HND or HNC or equivalent | 0.003 | 0 | 0.34 | -3.04 | 0.000 | 0 | 0.20 | -1.831 |
| Other professional qualifications eg: nursing, teach | -0.010 | 0 | 0.48 | -4.44 | -0.003 | 0 | 0.21 | -1.905 |
| A levels/AS levels or equivalent | -0.010 | 0 | 0.25 | -2.37 | -0.006 | 0 | 0.13 | -1.214 |
| O levels/GCSEs or equivalent | -0.011 | 0 | 0.33 | -3.08 | -0.004 | 0 | 0.16 | -1.512 |
| CSEs or equivalent | -0.007 | 0 | 0.14 | -1.37 | -0.002 | 0 | 0.15 | -1.390 |
| None of the above | -0.001 | 0 | 0.62 | -5.59 | 0.004 | 0 | 0.28 | -2.513 |
| Prefer not to answer | 0.000 | 0 | 0.46 | -4.11 | 0.008 | 0 | 0.27 | -2.364 |
| **Townsend deprivation score (quantiles)** | | | | | | | | |
| [-7,-4] | -0.009 | 0 | 0.28 | -2.63 | -0.004 | 0 | 0.10 | -0.977 |
| (-4,-2] | -0.010 | 0 | 0.32 | -2.96 | -0.004 | 0 | 0.14 | -1.283 |
| (-2,1] | -0.009 | 0 | 0.33 | -3.04 | -0.003 | 0 | 0.18 | -1.613 |
| (1,11] | -0.003 | 0 | 0.37 | -3.38 | 0.002 | 0 | 0.26 | -2.289 |
| **Index of multiple deprivation** | | | | | | | | |
| [0,20] | -0.010 | 0 | 0.31 | -2.90 | -0.004 | 0 | 0.14 | -1.261 |
| (20,40] | -0.006 | 0 | 0.34 | -3.16 | 0.000 | 0 | 0.22 | -1.970 |
| (40,60] | 0.000 | 0 | 0.38 | -3.44 | 0.005 | 0 | 0.28 | -2.521 |
| (60,80] | 0.009 | 0 | 0.43 | -3.76 | 0.007 | 0 | 0.31 | -2.770 |
| (80,100] | -0.001 | 0 | 0.43 | -3.89 | 0.018 | 0 | 0.35 | -3.016 |

### 10 TRIPOD+AI checklist

TRIPOD+AI

Version: 11-January-2024

| Section/Topic | Item | Development / evaluation[1] | Checklist item | Reported on page |
|---|---|---|---|---|
| **TITLE** | | | | |
| *Title* | 1 | D;E | Identify the study as developing or evaluating the performance of a multivariable prediction model, the target population, and the outcome to be predicted | 2 |
| **ABSTRACT** | | | | |
| *Abstract* | 2 | D;E | See TRIPOD+AI for Abstracts checklist | 2 |
| **INTRODUCTION** | | | | |
| *Background* | 3a | D;E | Explain the healthcare context (including whether diagnostic or prognostic) and rationale for developing or evaluating the prediction model, including references to existing models | 3 |
| | 3b | D;E | Describe the target population and the intended purpose of the prediction model in the context of the care pathway, including its intended users (e.g., healthcare professionals, patients, public) | 3 |
| | 3c | D;E | Describe any known health inequalities between sociodemographic groups | 3 |
| *Objectives* | 4 | D;E | Specify the study objectives, including whether the study describes the development or validation of a prediction model (or both) | 3 |
| **METHODS** | | | | |
| *Data* | 5a | D;E | Describe the sources of data separately for the development and evaluation datasets (e.g., randomised trial, cohort, routine care or registry data), the rationale for using these data, and representativeness of the data | 3 |
| | 5b | D;E | Specify the dates of the collected participant data, including start and end of participant accrual; and, if applicable, end of follow-up | 3 |
| *Participants* | 6a | D;E | Specify key elements of the study setting (e.g., primary care, secondary care, general population) including the number and location of centres | 3 |
| | 6b | D;E | Describe the eligibility criteria for study participants | 3 |
| | 6c | D;E | Give details of any treatments received, and how they were handled during model development or evaluation, if relevant | Not relevant |
| *Data preparation* | 7 | D;E | Describe any data pre-processing and quality checking, including whether this was similar across relevant sociodemographic groups | 6 |
| *Outcome* | 8a | D;E | Clearly define the outcome that is being predicted and the time horizon, including how and when assessed, the rationale for choosing this outcome, and whether the method of outcome assessment is consistent across sociodemographic groups | 5 |
| | 8b | D;E | If outcome assessment requires subjective interpretation, describe the qualifications and demographic characteristics of the outcome assessors | Not relevant |
| | 8c | D;E | Report any actions to blind assessment of the outcome to be predicted | Not relevant |
| *Predictors* | 9a | D | Describe the choice of initial predictors (e.g., literature, previous models, all available predictors) and any pre-selection of predictors before model building | Page 5, Table 1 |
| | 9b | D;E | Clearly define all predictors, including how and when they were measured (and any actions to blind assessment of predictors for the outcome and other predictors) | Page 5, Table 1 |
| | 9c | D;E | If predictor measurement requires subjective interpretation, describe the qualifications and demographic characteristics of the predictor assessors | Not relevant |
| *Sample size* | 10 | D;E | Explain how the study size was arrived at (separately for development and evaluation), and justify that the study size was sufficient to answer the research question. Include details of any sample size calculation | Page 4, Figure 1 |
| *Missing data* | 11 | D;E | Describe how missing data were handled. Provide reasons for omitting any data | x |
| *Analytical methods* | 12a | D | Describe how the data were used (e.g., for development and evaluation of model performance) in the analysis, including whether the data were partitioned, considering any sample size requirements | x |
| | 12b | D | Depending on the type of model, describe how predictors were handled in the analyses (functional form, rescaling, transformation, or any standardisation). | 4 |
| | 12c | D | Specify the type of model, rationale[2], all model-building steps, including any hyperparameter tuning, and method for internal validation | Not relevant |
| | 12d | D;E | Describe if and how any heterogeneity in estimates of model parameter values and model performance was handled and quantified across clusters (e.g., hospitals, countries). See TRIPOD-Cluster for additional considerations[3] | Not relevant |
| | 12e | D;E | Specify all measures and plots used (and their rationale) to evaluate model performance (e.g., discrimination, calibration, clinical utility) and, if relevant, to compare multiple models | 6 |
| | 12f | E | Describe any model updating (e.g., recalibration) arising from the model evaluation, either overall or for particular sociodemographic groups or settings | Not relevant |
| | 12g | E | For model evaluation, describe how the model predictions were calculated (e.g., formula, code, object, application programming interface) | 7 |
| *Class imbalance* | 13 | D;E | If class imbalance methods were used, state why and how this was done, and any subsequent methods to recalibrate the model or the model predictions | Not relevant |
| *Fairness* | 14 | D;E | Describe any approaches that were used to address model fairness and their rationale | 6 |
| *Model output* | 15 | D | Specify the output of the prediction model (e.g., probabilities, classification). Provide details and rationale for any classification and how the thresholds were identified | 7 |

[1] D=items relevant only to the development of a prediction model; E=items relating solely to the evaluation of a prediction model; D;E=items applicable to both the development and evaluation of a prediction model

[2] Separately for all model building approaches.

[3] TRIPOD-Cluster is a checklist of reporting recommendations for studies developing or validating models that explicitly account for clustering or explore heterogeneity in model performance (eg, at different hospitals or centres). Debray et al, BMJ 2023; 380: e071018 [DOI: 10.1136/bmj-2022-071018]

| | | | | |
|---|---|---|---|---|
| *Training versus evaluation* | 16 | D;E | Identify any differences between the development and evaluation data in healthcare setting, eligibility criteria, outcome, and predictors | Not relevant |
| *Ethical approval* | 17 | D;E | Name the institutional research board or ethics committee that approved the study and describe the participant-informed consent or the ethics committee waiver of informed consent | 4 |
| **OPEN SCIENCE** | | | | |
| *Funding* | 18a | D;E | Give the source of funding and the role of the funders for the present study | 14 |
| *Conflicts of interest* | 18b | D;E | Declare any conflicts of interest and financial disclosures for all authors | 14 |
| *Protocol* | 18c | D;E | Indicate where the study protocol can be accessed or state that a protocol was not prepared | 7 |
| *Registration* | 18d | D;E | Provide registration information for the study, including register name and registration number, or state that the study was not registered | Not relevant |
| *Data sharing* | 18e | D;E | Provide details of the availability of the study data | 4 |
| *Code sharing* | 18f | D;E | Provide details of the availability of the analytical code[4] | 7 |
| **PATIENT & PUBLIC INVOLVEMENT** | | | | |
| *Patient & Public Involvement* | 19 | D;E | Provide details of any patient and public involvement during the design, conduct, reporting, interpretation, or dissemination of the study or state no involvement. | Not relevant |
| **RESULTS** | | | | |
| *Participants* | 20a | D;E | Describe the flow of participants through the study, including the number of participants with and without the outcome and, if applicable, a summary of the follow-up time. A diagram may be helpful. | Page 4, Figure 1 |
| | 20b | D;E | Report the characteristics overall and, where applicable, for each data source or setting, including the key dates, key predictors (including demographics), treatments received, sample size, number of outcome events, follow-up time, and amount of missing data. A table may be helpful. Report any differences across key demographic groups. | Page 8, Table 2 Page S10 |
| | 20c | E | For model evaluation, show a comparison with the development data of the distribution of important predictors (demographics, predictors, and outcome). | Page 8, Table 2 Page S10 |
| *Model development* | 21 | D;E | Specify the number of participants and outcome events in each analysis (e.g., for model development, hyperparameter tuning, model evaluation) | Page S12 |
| *Model specification* | 22 | D | Provide details of the full prediction model (e.g., formula, code, object, application programming interface) to allow predictions in new individuals and to enable third-party evaluation and implementation, including any restrictions to access or re-use (e.g., freely available, proprietary)[5] | 4-7 |
| *Model performance* | 23a | D;E | Report model performance estimates with confidence intervals, including for any key subgroups (e.g., sociodemographic). Consider plots to aid presentation. | Page 9, Figure 2 Page S13 |
| | 23b | D;E | If examined, report results of any heterogeneity in model performance across clusters. See TRIPOD Cluster for additional details[3]. | Page 7, Figure 2 Page S13 |
| *Model updating* | 24 | E | Report the results from any model updating, including the updated model and subsequent performance | Not relevant |
| **DISCUSSION** | | | | |
| *Interpretation* | 25 | D;E | Give an overall interpretation of the main results, including issues of fairness in the context of the objectives and previous studies | 9 |
| *Limitations* | 26 | D;E | Discuss any limitations of the study (such as a non-representative sample, sample size, overfitting, missing data) and their effects on any biases, statistical uncertainty, and generalizability | 12 |
| *Usability of the model in the context of current care* | 27a | D | Describe how poor quality or unavailable input data (e.g., predictor values) should be assessed and handled when implementing the prediction model | 12 |
| | 27b | D | Specify whether users will be required to interact in the handling of the input data or use of the model, and what level of expertise is required of users | Not relevant |
| | 27c | D;E | Discuss any next steps for future research, with a specific view to applicability and generalizability of the model | 13 |

From: Collins GS, Moons KGM, Dhiman P, et al. *BMJ* 2024;385:e078378. doi:10.1136/bmj-2023-078378

[4] This relates to the analysis code, for example, any data cleaning, feature engineering, model building, evaluation.
[5] This relates to the code to implement the model to get estimates of risk for a new individual.
